# Using one–to–many urine proteome comparisons to provide clues for fever of unknown origin

**DOI:** 10.1101/2021.09.23.21264016

**Authors:** Chenyang Zhao, Lilong Wei, Jing Wei, Youhe Gao

## Abstract

**Objective:** To provide diagnostic evidence and clues for patients with fever of unknown origin (FUO) through urine proteomics analysis.

**Methods:** Urine samples of FUO were one–to–many analysed by using liquid chromatography tandem mass spectrometry(LC–MS/MS) to identify differential proteins and related biological pathways. One–to–many analysis means a comparative analysis of one sample to many controls.

**Results:** We observed biological pathways related to fever, such as LXR/RXR activation, FXR/RXR activation and acute phase response signaling, etc., which indicates that urine can obviously distinguish disease from health status. In addition, we found that the results of each sample were different, which highlight the necessity of one–to–many analysis.

**Conclusions:** The combined method of urine proteomics and one–to–many analysis can provide clues for FUO, and might also be applied to the exploration of any unknown disease.

## 1. Introduction

Fever is a common phenomenon in clinical manifestations. Since the concept of fever of unknown origin (FUO) was first proposed in 1961 ^[1]^, it has been an urgent problem to be solved in the field of medical diagnosis over the past 60 years. FUO refers to a type of disease that has a fever for more than 3 consecutive weeks with a body temperature of more than 38.3° several times and cannot be confirmed after multiple examinations for at least a week. According to relevant literature reports, there are more than 200 aetiologies that can cause fever, such as bacterial or viral infections, neoplastic diseases and so on ^[2]^.Despite the great development of medical diagnostic technology, there are still some doubts about the exact diagnosis of FUO patients. It is difficult to apply medicine to patients without clear aetiology. Using drugs blindly can delay the condition, and it is also a waste of medical resources. Combining the above problems, it is a challenge for us to provide new diagnostic methods for FUO patients, provide clues for diagnosis, and help the generation and verification of diagnosis.

Urine is a way for the body to excrete metabolic wastes. Without the control of homeostatic mechanisms, urine can accumulate small changes from the whole body, which is earlier and more sensitive than blood^[3]^. A large number of studies have shown that urine is a good source of disease biomarkers. For example, in a tumor–bearing rat model constructed by subcutaneous injection of Walker–256 cells, it was found that urine protein changes with the growth of tumor by monitoring the progression of cancer and urine protein^[4]^. At the same time, in the Walker–256 tail–vein injection rat model, it was found that when tumors grew in different organs, the urine proteome was different, which indicates the differential diagnostic ability of urine. For another example, in a rat model of drug–induced chronic pancreatitis, it was found that significant changes in urinary proteins reported to be associated with pancreatitis had occurred before pathological visualization ^[6]^, which reflected the early and highly sensitive nature of urine. All of these examples provided the basis for the early diagnosis of urine.

In this study, we pioneered a new method of comparison named one–to–many analysis, which means a comparative analysis of a patient and a group of healthy people. We believe that it is difficult to group patients into one group for analysis and comparison in the study of unknown disease. We have no definite evidence to group patients according to their characteristics, because of the unknowability of the disease. In actual clinical applications, doctors often judge a person for a disease based on his or her different performance which differs from the general public. In other words, we compare a person with all normal people, and beyond the normal range, there is a possibility of disease. Although there are differences among normal people, these differences fluctuate within a normal range. Through homogenization treatment within the group, a parameter closer to that of normal people can be obtained. When multiple proteins of patients with unknown diseases are found to differ from this parameter, we can analyse differential proteins by using existing bioinformatics methods to find abnormal biological processes or even damaged organs, to infer unknown diseases. Therefore, we carried out unlabelled quantitative proteomics analysis on the urine of each FUO sample to find biological pathways related to disease through differential proteins, and to provided clues and basis for the clinical diagnosis of FUO.

## 2. Methodology (Design/Approach)

### 2.1 Urine collection and ethics statement

This experiment was based on the reuse of discarded samples from Laboratory Medicine, and the process did not involve any patient identity information. We did not influence any treatment of the patient, nor did we recommend any clinical or auxiliary examination to the patient. The knowledge generated in this study is in the research stage, only for the acquisition of FUO clues.

A total of 13 urine samples were collected from 13 patients with fever on admission at China–Japan Friendship Hospital in Beijing, and the patients’ age ranged from 32 to 88 years. Eight healthy urine samples were collected from healthy donors of health care workers at Beijing Youan Hospital. Approximately 10–15 ml of midstream specimens of the morning urine was collected and stored at –80°C.

Two urine samples were abnormal in the test and clinically verified with urinary protein phenomena, which did not meet the sampling standards of this experiment, and as a result, they were excluded. All patients had fever at admission. On the day of urine sample collection, the patient’s temperature is recorded in Table 1. All participants signed informed consent forms, and the study received ethical approval from China–Japan Friendship Hospital (No. 2019–42–K30).

**Table 1.** Clinical characteristics record of samples

| Table 1. Clinical characteristics record of samples |  |  |
| --- | --- | --- |
| Number | sex | temperature/°C |
| F1 | female | 38.8 |
| F2 | female | 38.4 |
| F3 | male | 37.3 |
| F4 | male | 37.8 |
| F5 | male | 36.0 |
| F6 | male | 36.6 |
| F7 | male | 36.5 |
| F8 | male | 36.8 |
| F9 | female | 36.6 |
| F10 | male | 36.8 |
| F11 | female | 36.9 |

Ps: The temperature recorded when collecting urine samples.

### 2.2 Urine sample preparation

Four millilitres of urine sample reheated at 37°C was centrifuged at 4°C and 12,000g for 20min to remove cell and cell debris and retain the supernatants. Next, 20 mM dithiothreitol (DTT) was added and heated at 99.2°C for 10 min to cool to room temperature. Then 50 mM iodoacetamide (IAA) was added and mixed at room temperature for 40 min in the dark. After the reaction finished, the supernatants were placed into new Eppendorf tubes and mixed with four volumes of ethanol at –20°C overnight to precipitate protein. Mixed liquid was centrifuged at 4°C and 10,000g for 30min, and the precipitate was retained and dried. Proper lysis buffer (8 mol/L urea, 2 mol/L thiourea, 50 mmol/L Tris, and 25 mmol/L DTT) was added to redissolve the protein precipitate and was mixed at 4°C for 2h. The supernatants were retained after centrifugation at 4°C and 12,000g for 30min, and each sample’s protein concentration was measured by the Bradford method.

The FASP method^[7]^ was used to digest urine protein with trypsin (Trypsin Gold, Mass Spec Grade, Promega, Fitchburg, Wisconsin, USA). One hundred micrograms of urine protein was added in the membrane of a 10KD ultrafiltration tube (Pall, Port Washington, NY) and washed twice with UA (8 M urea, 0.1 mol/L Tris–HCl, pH 8.5) and NH4HCO3 (25 mmol/L) at 14,000 g for 40 min at 18 °C. Then, the urine protein was resuspended in 25mmol/L NH4HCO3 and digested by trypsin (enzyme to protein ratio of 1:50) at 37 °C for 14 h–16 h. We obtained the peptides after two centrifugations at 4°C and 12,000g for 30min. These peptides were desalted using Oasis HLB cartridges (Waters, Milford, MA) and then dried by Speed Vac (Thermo Fisher Scientific, Bremen, Germany). Then, the peptides were stored at –80°C.

### 2.3 Peptide fractionation

We applied a method to improve the ability to identify low–abundance peptides. The digested peptides were redissolved in 0.1% formic acid and quantified to 0.5 μg/μL which was measured by the BCA method (Thermo Scientific). A pooled sample (73.5 μg, 3.5 μg of each sample) from 21 samples was loaded onto an equilibrated, high–pH, reversed–phase fractionation spin column (84868, Thermo Scientific) to generate the spectral library for subsequent analysis. Two fractions were collected by pooled sample and water. The remaining eight different fractions were collected by a step gradient of increasing acetonitrile concentrations (5.0, 7.5, 10.0, 12.5, 15.0, 17.5, 20.0 and 50.0% acetonitrile) in a volatile high–pH elution solution. Then the ten fractions were dried by Speed Vac (Thermo Scientific, Bremen, Germany) and resuspended in 20 µl of 0.1% formic acid for LC– MS/MS analysis.

### 2.4 LC–MS/MS analysis

The iRT reagent (Biognosys, Switzerland) was used to calibrate the retention time of the extracted peptide peaks and it was added at a ratio of 1:10 v/v to all of the peptide samples. For analysis, 1 μg of each peptide sample was loaded onto a trap column (75 µm × 2 cm, 3 µm, C18, 100 Å) and separated on a reverse–phase C18 column (50 µm × 15 cm, 2 µm, C18, 100 Å) using the EASY–nLC 1200 HPLC system (Thermo Fisher Scientific, Waltham, MA). The elution gradient was 4%–35% buffer B (0.1% formic acid in 80% acetonitrile, flow rate, 0.4 μL/min) for 90 min. Eluted peptides were analysed by using an Orbitrap Fusion Lumos Tribrid Mass Spectrometer (Thermo Fisher Scientific, Waltham, MA, USA).

To generate the spectral library, the ten fractions from the reversed–phase fractionation spin column were analysed by data– dependent (DDA) MS/MS acquisition mode with a resolution of 120,000 in full scan mode and 30,000 in MS/MS mode. The full scan was performed in the Orbitrap from 350–1,500 m/z, and the high–energy collisional dissociation (HCD) energy was set to 30%. The mass spectrometric parameters included the following: the automatic gain control (AGC) target was set to 4e5; the maximum injection time was 45 ms.

Twenty–one single peptide samples were analysed by data–independent (DIA) MS/MS acquisition mode, and the variable isolation window of the DIA method with 36 windows was applied to DIA acquisition. The mass parameters contained the dollowing: the fall scan was obtained from 350 to 1,500 m/z with a resolution of 60,000, and the DIA scan was obtained from 200 to 2,000 m/z with a resolution of 30,000; the HCD energy was set to 32%; the AGC target was set to 1e6 and the maximum injection time was 100 ms.

### 2.5 Data analysis

Ten raw files of DDA data collected from the mass spectrometer were searched by Proteome Discoverer (version: 2.1, Thermo Scientific, USA) against the Swiss–Prot Home Sapiens Human database, which also includes the iRT sequence. The retrieval arguments contained: tryptic digestion; two missed trypsin cleavage sites were allowed; the carbamidomethyl of cysteine was set as a fixed modification; the oxidation of methionine was set as a variable modification; the fragment ion mass tolerance was 0.02 Da; and the parent ion mass tolerance was set to 10 ppm. The false discovery rate (FDR) of proteins and peptides was less than 1%. A total of 2957 protein groups were identified from the library and the result file was used in Spectronaut™ Pulsar X (Biognosys, Switzerland) software for the subsequent analysis. Nine QC and 21 raw files of DIA data were analysed with the default settings by using the Spectronaut, and the results were filtered depending on having Q–value of less than 0.01 (corresponding to an FDR of 1%). Proteins recognized by more than two unique peptides were retained. The peptide intensity was calculated by summing the peak areas of the respective fragment ions from MS2 and the protein intensity was calculated by summing the intensities of each peptide.

### 2.6 Statistical analysis

The missing values of protein abundance were filled with the k–nearest neighbour (K–NN) method. Differential proteins were identified by the comparison of fever samples and healthy samples with two–sided paired t–tests. The differential protein selection parameters were as follows: fold change≥2 or ≤0.5, p values<0.05 (p values corrected by the BH method). Differential protein canonical pathways were generated by Ingenuity Pathway Analysis (IPA) software (Mountain View, CA, USA), and the functions of differential proteins were searched in the reported literature based on the PubMed database (https://pubmed.ncbi.nlm.nih.gov).

## 3. Results and discussion

### 3.1 One–to–many urine proteome analysis of single patients

#### 3.1.1 Urine proteome analysis of F1 patient

A total of 108 differential proteins were identified by comparing the F1 sample with the healthy group samples, including 85 upregulated proteins and 23 downregulated proteins. The differential protein screening parameters were FC≥2 or ≤0.5, and P<0.05, and the specific information on the differential proteins is listed in Table S1.

Ingenuity Pathway Analysis was used to analyse the differential proteins, and the classical pathway results obtained are listed in Table 2. P<0.05 was considered to be convincing. The signal pathways enriched by differential proteins were mainly manifested in LXR/RXR activation, acute phase response signaling, antigen presentation pathway, FXR/RXR activation, phagosome maturation, folate polyglutamylation, super pathway of serine and glycine biosynthesis I, sucrose degradation V, protein ubiquitination pathway and other signaling pathways. According to the relevant literature reports, the acute phase response signaling pathway ^[8]^ is related to fever; LXR/RXR (liver X receptor/retinoid X receptor) and FXR/RXR (farnesoid–X– receptor/retinoid–X–receptor) usually exist in the biological and pathological pathways involved in glucose and lipid homeostasis and inflammatory response ^[9]^; antigen presentation and phagosome maturation are closely related to the immune process. At the same time, patient F1 was in an acute reaction state of fever (body temperature 38.8°C) when the samples were collected. These results indicated that the changes reflected in urine protein are consistent with clinical manifestations.

**Table 2.**
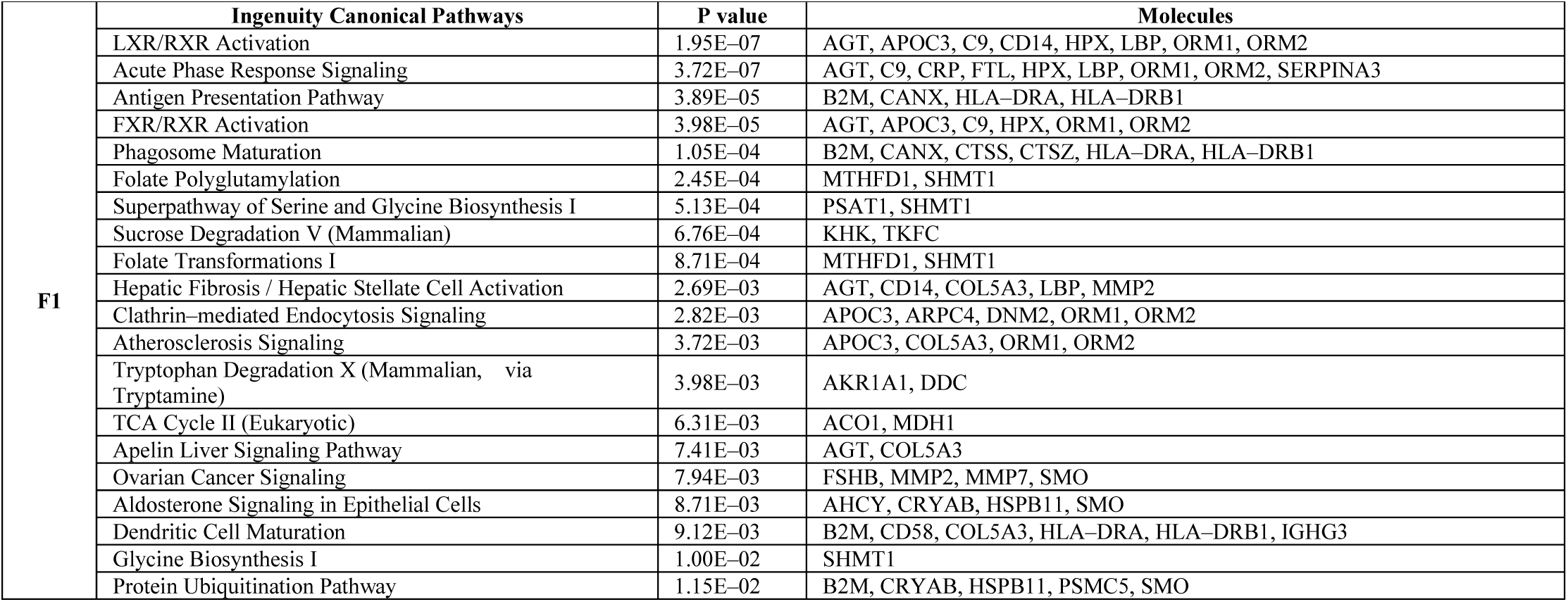
F1 IPA analysis results

#### 3.1.2 Urine proteome analysis of F2 patient

A total of 174 differential proteins were identified by comparing the F2 sample with the healthy group samples, including 131 upregulated proteins and 43 downregulated proteins. The differential protein screening parameters were FC≥2 or ≤0.5, and P<0.05, and the specific information on the differential proteins is listed in Table S1.

Ingenuity Pathway Analysis was used to analyse the differential proteins, and the classical pathway results obtained are listed in Table 3. P<0.05 was considered to be convincing. The signal pathways enriched by differential proteins were mainly manifested in phagosome maturation, antigen presentation pathway, LXR/RXR activation, PD–L1 cancer immunotherapy pathway, Th1 and Th2 activation pathway, FXR/RXR activation, acute phase response signaling, virus entry via endocytic pathways, natural killer cell signaling, B cell development and other pathways through endocytosis. In addition to the previously mentioned signaling pathways related to inflammation, patient F2 showed more signals related to viruses and immunity, such as the signaling pathways of NK cells and B cells and the process of endocytosis of viruses into cells, which indicates that the fever is possibly caused by virus invasion. Then we searched the virus sequence, and failed to find a clear and convincing pathogenic virus. Disease–causing viruses need more in–depth examination.

**Table 3.**
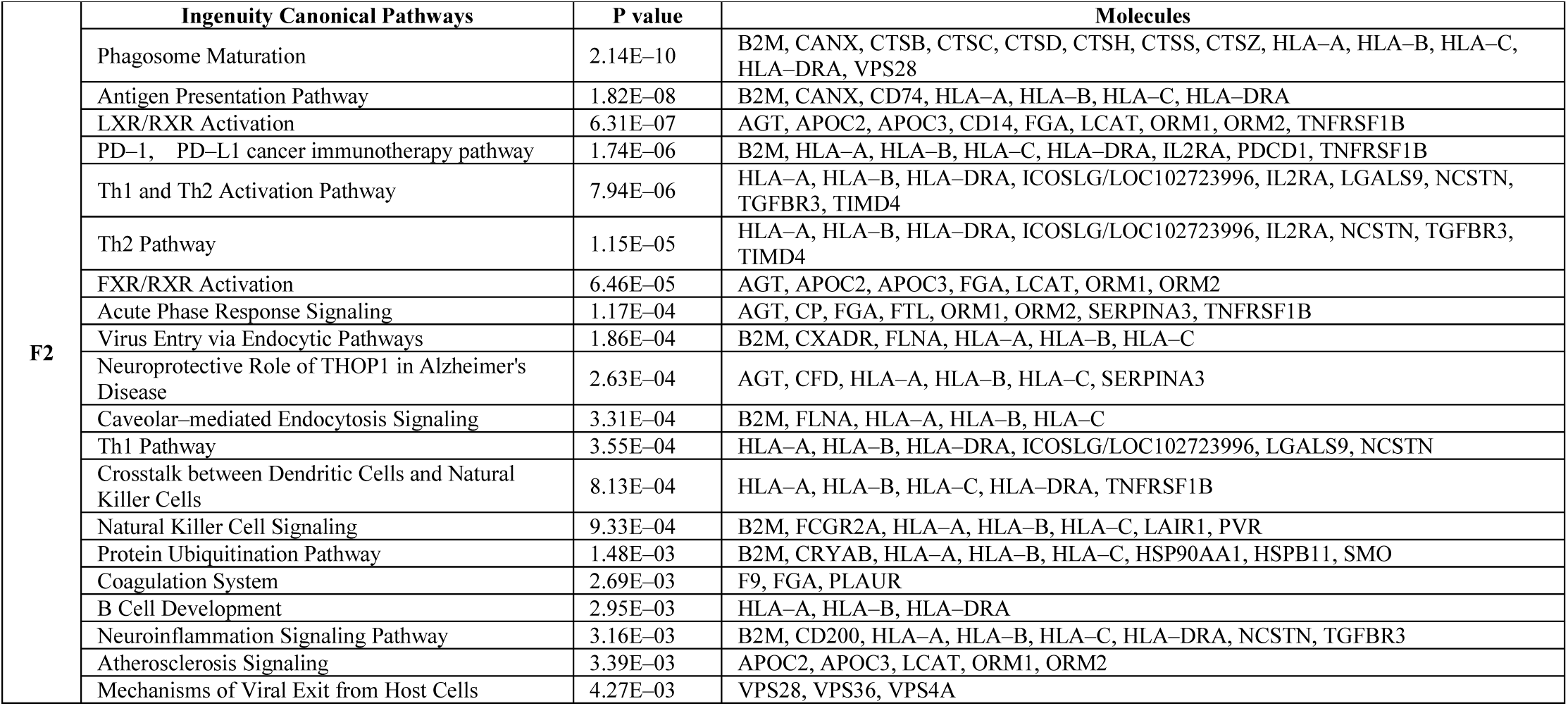
F2 IPA analysis results

#### 3.1.3 Urine proteome analysis of F3 patient

A total of 65 differential proteins were identified by comparing the F3 sample with the healthy group samples, including 50 upregulated proteins and 15 downregulated proteins. The differential protein screening parameters were FC≥2 or ≤0.5, and P<0.05, and the specific information on the differential proteins is listed in Table S1.

Ingenuity Pathway Analysis was used to analyse the differential proteins, and the classical pathway results obtained are listed in Table 4. P<0.05 was considered to be convincing. The signal pathways enriched by differential proteins were mainly manifested in acute phase response, LXR/RXR activation, FXR/RXR activation, complement system, phagosome maturation, airway pathology in chronic obstructive, bladder cancer signaling, inhibition of matrix metalloproteases, atherosclerosis signaling, uracil degradation, thymine degradation, thyroid hormone biosynthesis, ovarian cancer signaling, tumor microenvironment pathway and other signaling pathways. Similarly, the first few pathways that are most significantly related are shown to be related to inflammation.

**Table 4.**
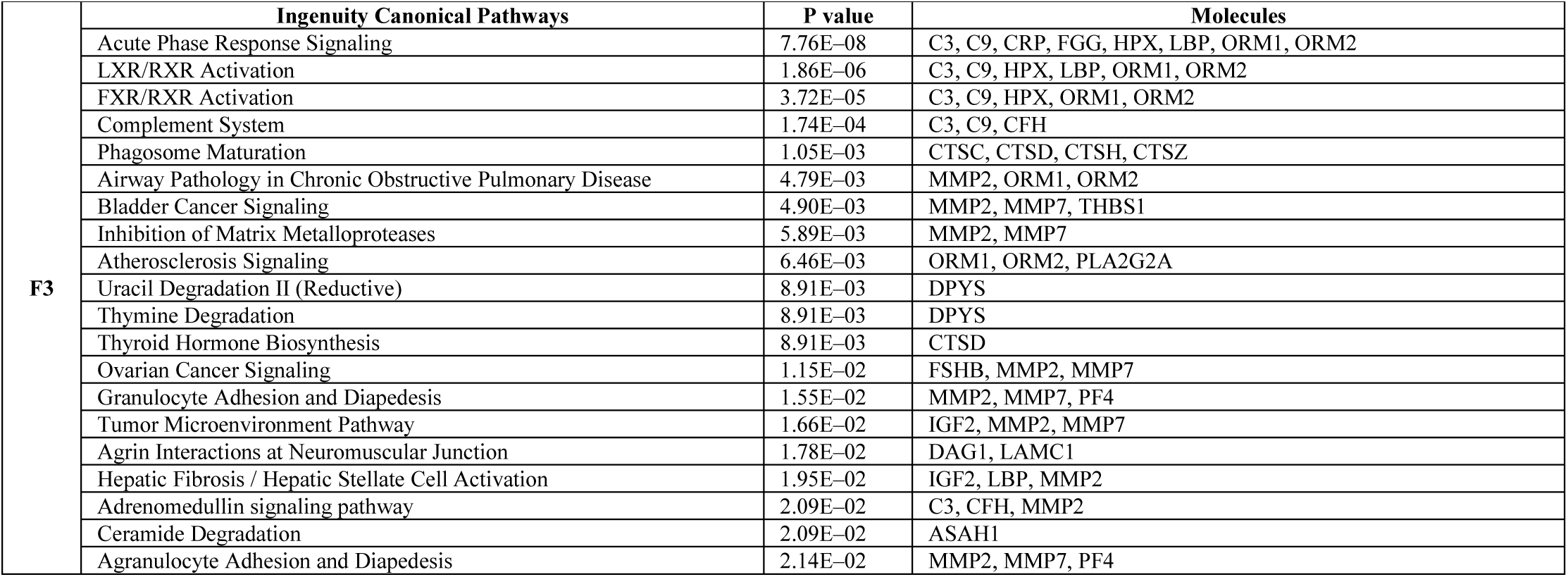
F3 IPA analysis results

#### 3.1.4 Urine proteome analysis of F4 patient

A total of 440 differential proteins were identified by comparing the F4 sample with the healthy group samples, including 279 upregulated proteins and 161 downregulated proteins. The differential protein screening parameters were FC≥2 or ≤0.5, and P<0.05, and the specific information on the differential proteins is listed in Table S1.

Ingenuity Pathway Analysis was used to analyse the differential proteins, and the classical pathway results obtained are listed in Table 5. P<0.05 was considered to be convincing. The signal pathways enriched by differential proteins were mainly manifested in clathrin–mediated endocytosis signaling, epithelial adhesion junction signaling, remodeling adhesion junction signaling, BAG2 signaling pathway, synaptogenesis signaling pathway, reelin signaling in neurons and glycolysis pathway. It also has the same acute inflammation–related pathways as the F1, F2, and F3 samples.

**Table 5.**
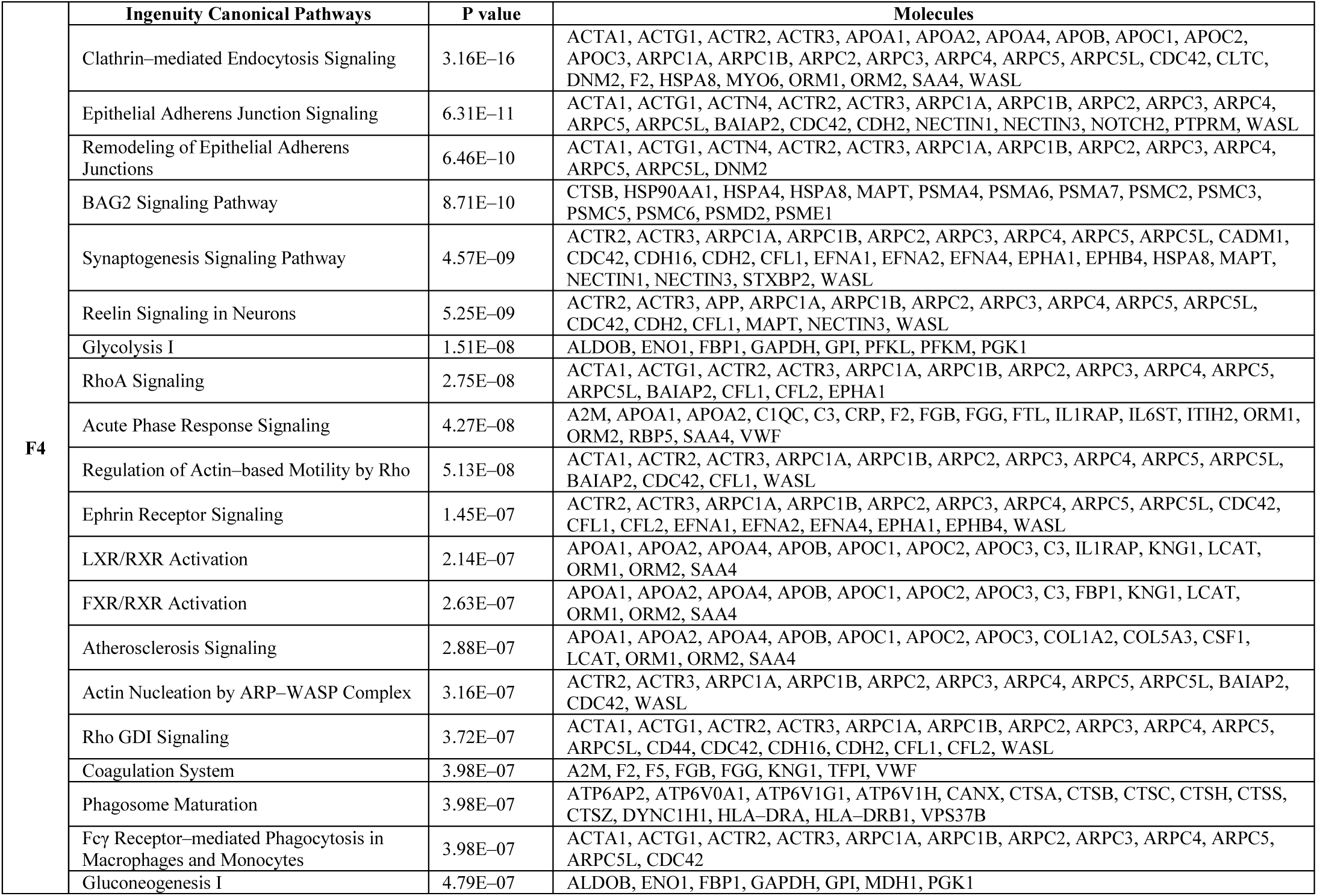
F4 IPA analysis results

#### 3.1.5 Urine proteome analysis of F5 patient

A total of 184 differential proteins were identified by comparing the F5 sample with the healthy group samples, including 143 upregulated proteins and 41 downregulated proteins. The differential protein screening parameters were FC≥2 or ≤0.5, and P<0.05, and the specific information on the differential proteins is listed in Table S1.

Ingenuity Pathway Analysis was used to analyse the differential proteins, and the classical pathway results obtained are listed in Table 6. P<0.05 was considered to be convincing. The signaling pathways enriched by differential proteins were mainly manifested in phagosome maturation, chondroitin sulfate degradation, Ephrin receptor signaling, iron homeostasis signaling pathway, role of MAPK signaling in promoting the pathogenesis of influenza, sphingosine–1–phosphate signaling, axonal guidance signaling, RhoA signal, and so on.

**Table 6.**
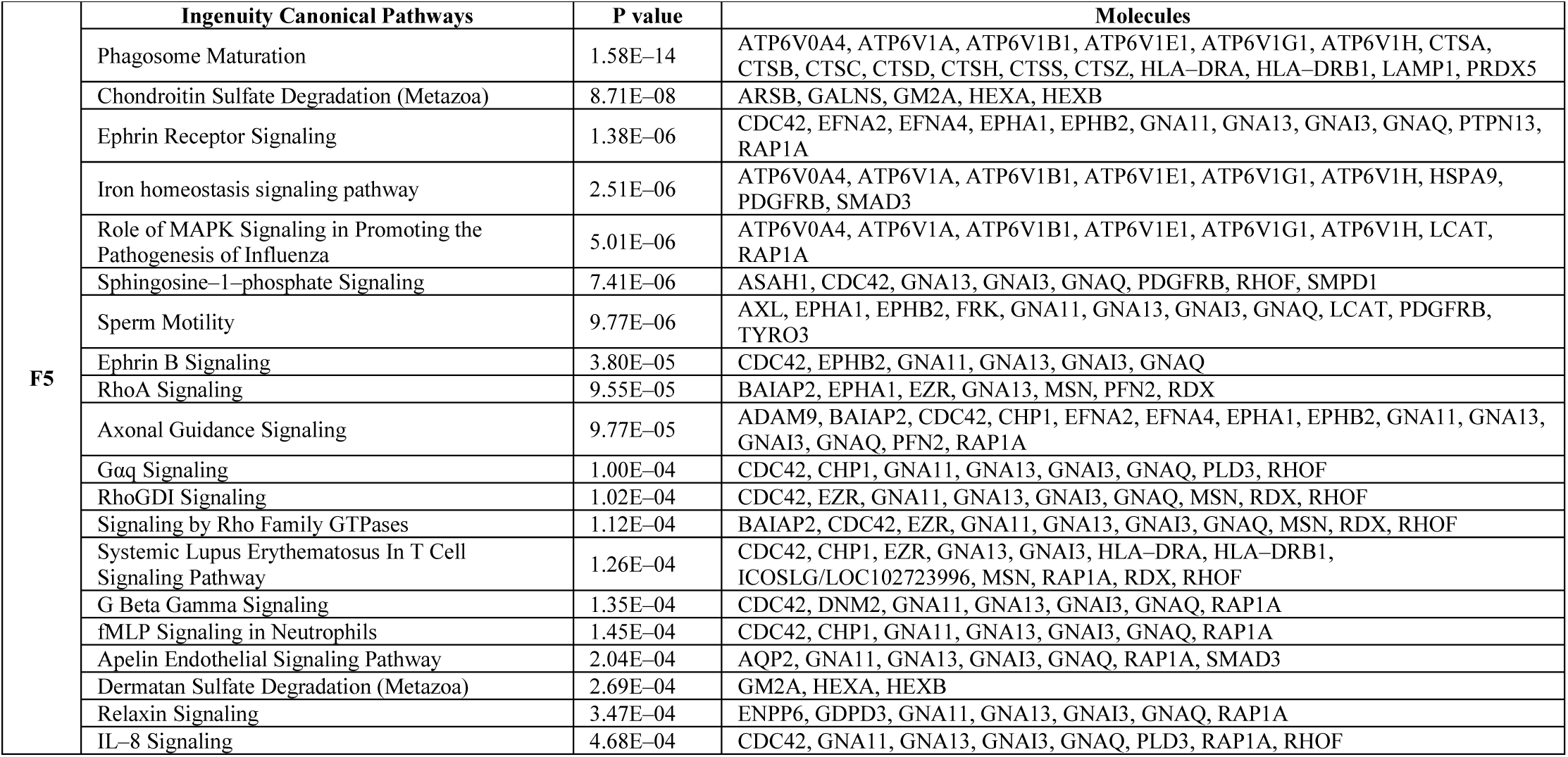
F5 IPA analysis results

#### 3.1.6 Urine proteome analysis of F6 patient

A total of 456 differential proteins were identified by comparing the F6 sample with the healthy group samples, including 298 upregulated proteins and 158 downregulated proteins. The differential protein screening parameters were FC≥2 or ≤0.5, and P<0.05, and the specific information on the differential proteins is listed in Table S1.

Ingenuity Pathway Analysis was used to analyse the differential proteins, and the classical pathway results obtained are listed in Table 7. P<0.05 was considered to be convincing. The signal pathways enriched by differential proteins were mainly manifested in LXR/RXR activation, FXR/RXR activation, phagosome maturation, acute phase response signaling, coagulation system, antigen presentation pathway, intrinsic prothrombin activation pathway, hepatic fibrosis/hepatic stellate cell activation, crosstalk between dendritic cells and natural killer cells, neuroprotective role of THOP1 in Alzheimer’s disease, extrinsic prothrombin activation pathway, Th2 pathway, Th1 and Th2 activation pathway, mechanism of viral exit from host cells, atherosclerosis signaling, dendritic cell maturation, B cell development, and so on.

**Table 7.**
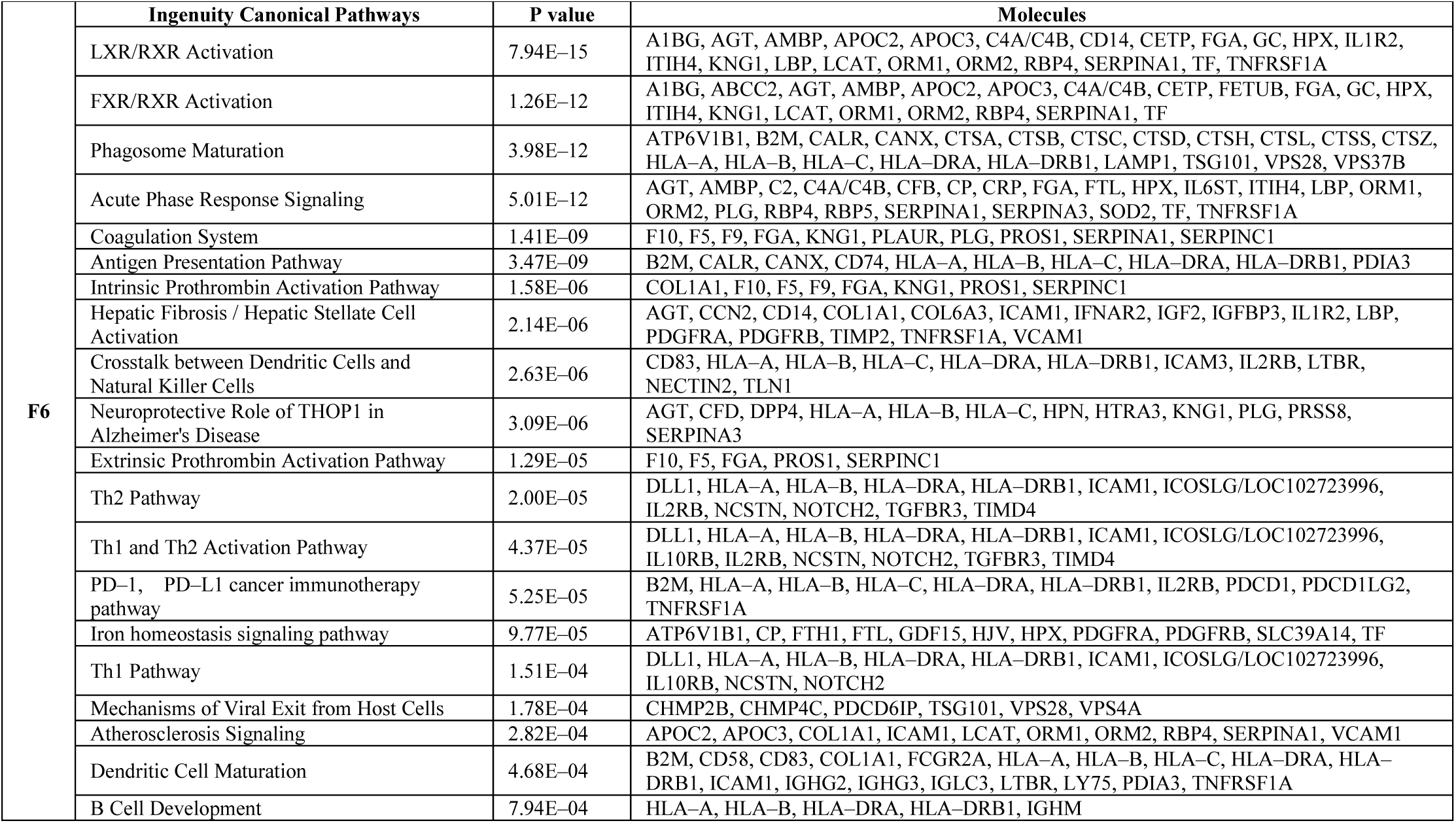
F6 IPA analysis results

#### 3.1.7 Urine proteome analysis of F7 patient

A total of 170 differential proteins were identified by comparing the F7 sample with the healthy group samples, including 153 upregulated proteins and 17 downregulated proteins. The differential protein screening parameters were FC≥2 or ≤0.5, and P<0.05, and the specific information on the differential proteins is listed in Table S1.

Ingenuity Pathway Analysis was used to analyse the differential proteins, and the classical pathway results obtained are listed in Table 8. P<0.05 was considered to be convincing. The signal pathways enriched by differential proteins were mainly manifested in phagosome maturation, LXR/RXR activation, FXR/RXR activation, lactose degradation, phagosome formation, atherosclerosis signaling, clathrin–mediated endocytic signaling, IL–12 signaling and production in macrophages, sphingosine 1–phosphate signal, vitamin C transport pathway, remodeling of epithelial adhesion junctions, production of nitric oxide and reactive oxygen species in macrophages, caveolar–mediated endocytosis signaling, L–carnitine biosynthesis, and so on.

**Table 8.**
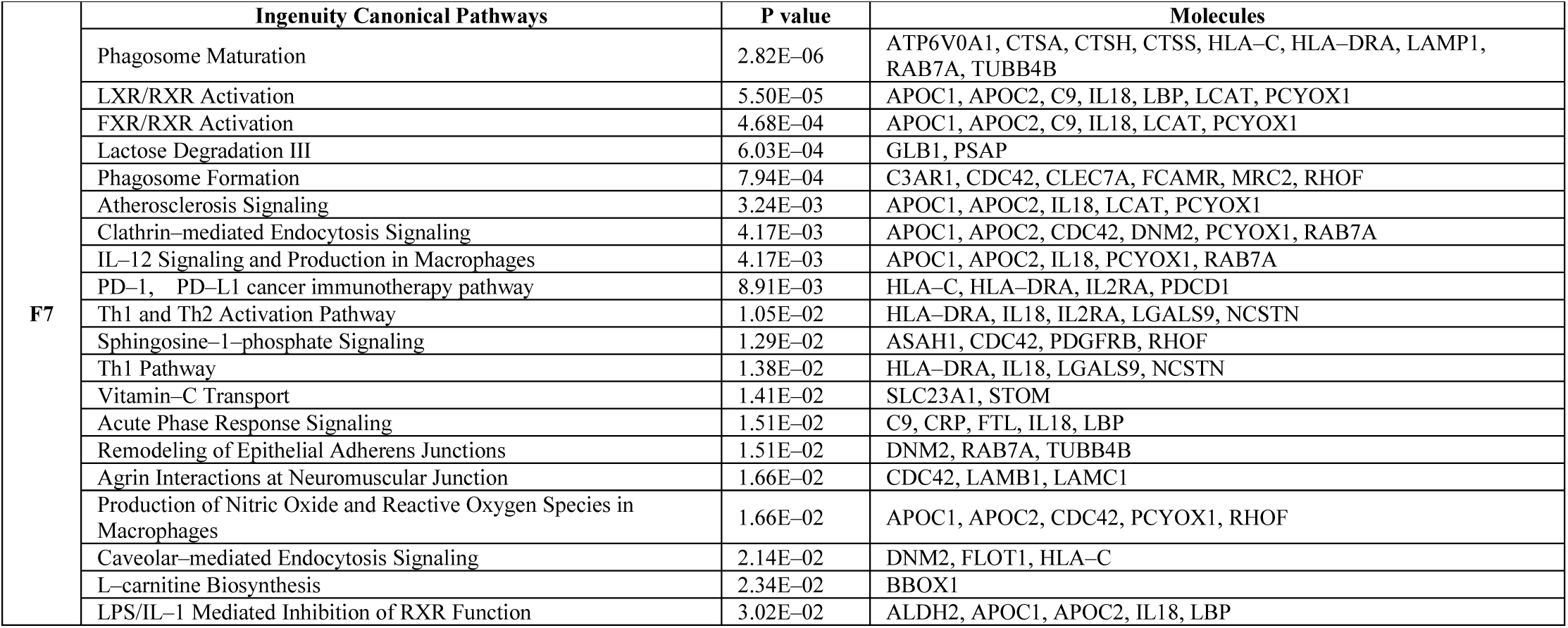
F7 IPA analysis results

#### 3.1.8 Urine proteome analysis of F8 patient

A total of 563 differential proteins were identified by comparing the F8 sample with the healthy group samples, including 460 upregulated proteins and 103 downregulated proteins. The differential protein screening parameters were FC≥2 or ≤0.5, and P<0.05, and the specific information on the differential proteins is listed in Table S1.

Ingenuity Pathway Analysis was used to analyse the differential proteins, and the classical pathway results obtained are listed in Table 9. P<0.05 was considered to be convincing. The signal pathways enriched by differential proteins were mainly manifested in hepatic fibrosis/hepatic stellate cell activation, LXR/RXR activation, phagosome maturation, intrinsic prothrombin activation pathway, atherosclerosis signaling, acute phase response signaling, iron homeostasis Signal pathway, natural killer cell signaling, apelin liver signaling pathway, antigen presentation pathway, FXR/RXR activation, Th1 and Th2 activation pathway, Caveolar–mediated endocytosis signaling, Th2 pathway, complement system, and so on.

**Table 9.**
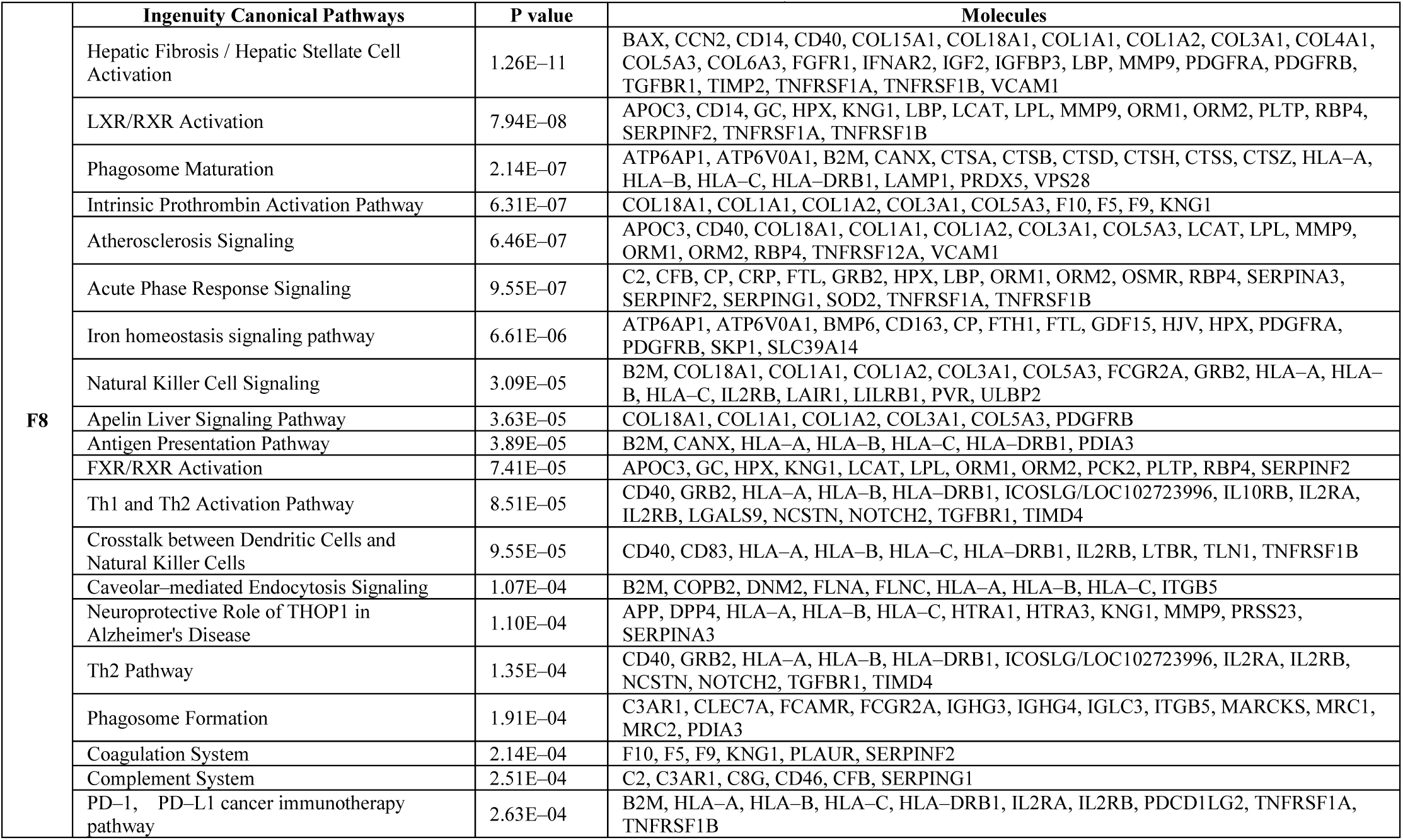
F8 IPA analysis results

#### 3.1.9 Urine proteome analysis of F9 patient

A total of 36 differential proteins were identified by comparing the F9 sample with the healthy group samples, including 32 upregulated proteins and 4 downregulated proteins. The differential protein screening parameters were FC≥2 or ≤0.5, and P< 0.05, and the specific information on the differential proteins is listed in Table S1.

Ingenuity Pathway Analysis was used to analyse the differential proteins, and the classical pathway results obtained are listed in Table 10. P<0.05 was considered to be convincing. The signal pathways enriched by differential proteins were mainly manifested in primary immunodeficiency signaling, ovarian cancer signaling, B cell development, autoimmune thyroid disease signaling, autoimmune thyroid disease signaling, neuroprotective role of THOP1 in Alzheimer’s disease, dendritic cell maturation, role of NFAT in regulating of the immune response, systemic lupus erythematosus signaling, relaxin signaling, glutaryl–CoA degradation, acute phase response signaling, GNRH Signaling, B cell receptor signaling, role of JAK1, JAK2 and TYK2 in interferon signaling, NRF2–mediated oxidative stress response, and so on.

**Table 10.** F9 IPA analysis results

| F9 | Ingenuity Canonical Pathways | P value | Molecules |
| --- | --- | --- | --- |
|  | Primary Immunodeficiency Signaling | 4.79E-05 | IGHG2, IGHM, IGLL1/IGLL5 |
|  | Ovarian Cancer Signaling | 7.94E-05 | CGA, FSHB, LHB, MMP9 |
|  | B Cell Development | 1.29E-03 | HLA-DRB1, IGHM |
|  | Autoimmune Thyroid Disease Signaling | 4.37E-03 | CGA, HLA-DRB1, IGHG2 |
|  | Communication between Innate and Adaptive Immune Cells | 6.92E-03 | HLA-DRB1, IGHG2, IGHM |
|  | Neuroprotective Role of THOP1 in Alzheimer's Disease | 1.17E-02 | MMP9, SERPINA3 |
|  | Superoxide Radicals Degradation | 1.17E-02 | SOD1 |
|  | Dendritic Cell Maturation | 1.55E-02 | FCGR3A/FCGR3B, HLA-DRB1, IGHG2 |
|  | Role of NFAT in Regulation of the Immune Response | 1.58E-02 | FCGR3A/FCGR3B, HLA-DRB1, IGHM |
|  | Phagosome Formation | 1.78E-02 | FCGR3A/FCGR3B, IGHG2 |
|  | Systemic Lupus Erythematosus Signaling | 1.95E-02 | FCGR3A/FCGR3B, IGHG2, IGHM |
|  | Phagosome Maturation | 2.04E-02 | HLA-DRB1, PRDX2 |
|  | Relaxin Signaling | 2.09E-02 | GDPD3, MMP9 |
|  | Glutaryl-CoA Degradation | 2.51E-02 | CA1 |
|  | Acute Phase Response Signaling | 3.02E-02 | FTL, SERPINA3 |
|  | GNRH Signaling | 3.02E-02 | FSHB, LHB |
|  | B Cell Receptor Signaling | 3.16E-02 | IGHG2, IGHM |
|  | Tryptophan Degradation III (Eukaryotic) | 3.47E-02 | CA1 |
|  | Role of JAK1, JAK2 and TYK2 in Interferon Signaling | 3.80E-02 | CGA |
|  | NRF2-mediated Oxidative Stress Response | 3.89E-02 | FTL, SOD1 |

#### 3.1.10 Urine proteome analysis of F10 patient

A total of 409 differential proteins were identified by comparing the F10 sample with the healthy group samples, including 272 upregulated proteins and 137 downregulated proteins. The differential protein screening parameters were FC≥2 or ≤0.5, and P<0.05, and the specific information on the differential proteins is listed in Table S1.

Ingenuity Pathway Analysis was used to analyse the differential proteins, and the classical pathway results obtained are listed in Table 11. P<0.05 was considered to be convincing. The signaling pathways enriched by differential proteins were mainly manifested in LXR/RXR activation, FXR/RXR activation, acute phase response signaling, phagosome maturation, antigen presentation pathway, extrinsic prothrombin activation pathway, mechanism of viral exit from host cells, ascorbate recycling, Wnt/β–catenin signaling, atherosclerosis signaling, caveolar–mediated endocytosis signaling, natural killer cell signaling, and so on.

**Table 11.**
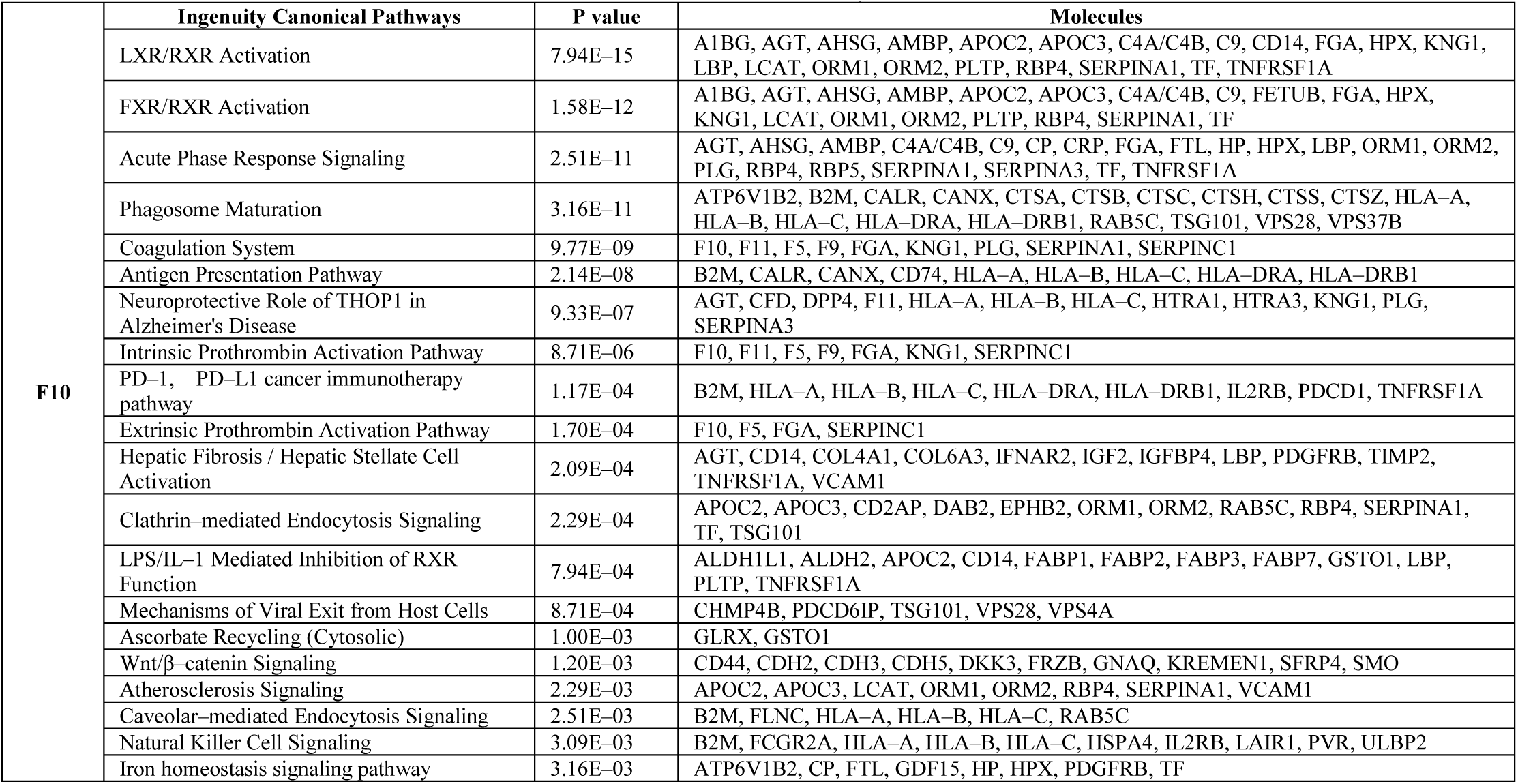
F10 IPA analysis results

#### 3.1.11 Urine proteome analysis of F11 patient

A total of 294 differential proteins were identified by comparing the F11 sample with the healthy group samples, including 217 upregulated proteins and 77 downregulated proteins. The differential protein screening parameters were FC≥2 or ≤0.5, and P<0.05, and the specific information on the differential proteins is listed in Table S1.

Ingenuity Pathway Analysis was used to analyse the differential proteins, and the classical pathway results obtained are listed in Table 12. P<0.05 was considered to be convincing. The signal pathways enriched by differential proteins were mainly manifested in the acute phase response signaling, antigen presentation pathway, neuroprotective role of THOP1 in Alzheimer’s disease, FAT10 signaling pathway, PD–1, PD–L1 cancer immunotherapy pathway, polyamine regulation in colon cancer, BAG2 signaling pathway, protein ubiquitination pathway, phagosome maturation, LXR/RXR activation, B cell development, inhibition of ARE–mediated mRNA degradation pathway, Th2 pathway, neuroinflammation signaling pathway, role of IL–17A in psoriasis, complement system, and so on.

**Table 12.** F11 IPA analysis results

| F11 | Ingenuity Canonical Pathways | P value | Molecules |
| --- | --- | --- | --- |
|  | Acute Phase Response Signaling | 1.45E-10 | AGT, APOH, C1R, C1S, C2, CP, CRP, FTL, IL6ST, ORM1, ORM2, PLG, RBP5, SERPINA3, SERPING1, SOD2, TNFRSF1B |
|  | Antigen Presentation Pathway | 6.92E-10 | B2M, CANX, HLA-A, HLA-B, HLA-C, HLA-DRA, HLA-DRB1, PDIA3, PSMB9 |
|  | Neuroprotective Role of THOP1 in Alzheimer's Disease | 1.32E-07 | AGT, C1R, DPP4, HLA-A, HLA-B, HLA-C, HPN, KLK6, PLG, PRTN3, SERPINA3 |
|  | FAT10 Signaling Pathway | 3.09E-07 | PSMA1, PSMA4, PSMA5, PSMA6, PSMA7, PSMB1, PSMB4, PSMB9 |
|  | PD-1, PD-L1 cancer immunotherapy pathway | 6.03E-07 | B2M, HLA-A, HLA-B, HLA-C, HLA-DRA, HLA-DRB1, IL2RB, PDCD1, PDCD1LG2, TNFRSF1B |
|  | Polyamine Regulation in Colon Cancer | 7.08E-07 | PSMA1, PSMA4, PSMA5, PSMA6, PSMA7, PSMB1, PSMB4, PSMB9 |
|  | BAG2 Signaling Pathway | 7.24E-06 | PSMA1, PSMA4, PSMA5, PSMA6, PSMA7, PSMB1, PSMB4, PSMB9 |
|  | Protein Ubiquitination Pathway | 7.24E-06 | B2M, CRYAB, HLA-A, HLA-B, HLA-C, HSPB11, PSMA1, PSMA4, PSMA5, PSMA6, PSMA7, PSMB1, PSMB4, PSMB9 |
|  | Phagosome Maturation | 1.70E-05 | B2M, CANX, CTSS, CTSZ, HLA-A, HLA-B, HLA-C, HLA-DRA, HLA-DRB1, LAMP1 |
|  | LXR/RXR Activation | 2.24E-05 | A1BG, AGT, APOC1, APOH, CD14, ORM1, ORM2, S100A8, TNFRSF1B |
|  | B Cell Development | 7.59E-05 | HLA-A, HLA-B, HLA-DRA, HLA-DRB1, IGHM |
|  | Crosstalk between Dendritic Cells and Natural Killer Cells | 1.35E-04 | HLA-A, HLA-B, HLA-C, HLA-DRA, HLA-DRB1, IL2RB, TNFRSF1B |
|  | Inhibition of ARE-Mediated mRNA Degradation Pathway | 1.66E-04 | PSMA1, PSMA4, PSMA5, PSMA6, PSMA7, PSMB1, PSMB4, PSMB9, TNFRSF1B |
|  | Th2 Pathway | 2.51E-04 | HLA-A, HLA-B, HLA-DRA, HLA-DRB1, ICOSLG/LOC102723996, IL2RB, NCSTN, TGFBR3 |
|  | Caveolar-mediated Endocytosis Signaling | 3.31E-04 | B2M, COPB2, HLA-A, HLA-B, HLA-C, ITGB1 |
|  | Neuroinflammation Signaling Pathway | 4.37E-04 | B2M, CD200, CRP, CX3CL1, HLA-A, HLA-B, HLA-C, HLA-DRA, HLA-DRB1, NCSTN, SOD2, TGFBR3 |
|  | Role of IL-17A in Psoriasis | 6.17E-04 | S100A7, S100A8, S100A9 |
|  | Huntington's Disease Signaling | 6.76E-04 | CAPN1, CAPNS1, GNAQ, PSMA1, PSMA4, PSMA5, PSMA6, PSMA7, PSMB1, PSMB4, PSMB9 |
|  | FXR/RXR Activation | 9.33E-04 | A1BG, ABCC2, AGT, APOC1, APOH, ORM1, ORM2 |
|  | Complement System | 9.77E-04 | C1R, C1S, C2, SERPING1 |

### 3.2 Group analysis of urinary proteome in patients

Nineteen samples (eleven FUO samples and eight healthy samples) were used for label–free LC–MS/MS quantitation. A total of 2812 proteins with at least 2 unique peptides were identified with FDR<1%. In addition, 129 differential proteins were identified by comparing all FUO samples with all healthy group samples, including 20 upregulated proteins and 109 downregulated proteins. FC≥2 or ≤0.5, and P<0.05, and the specific information on the differential proteins is listed in Table S1.

Ingenuity Pathway Analysis was used to analyse the differential proteins, and the classical pathway results obtained are listed in Table 13. P<0.05 was considered to be convincing. The main signal pathways enriched by differential proteins were the STAT3 pathway, SPINK1 pancreatic cancer pathway, interleukin–15 production, and phagosome maturation, and so on. Among them, the STST3 pathway is closely related to autophagy ^[10]^, and interleukin 15 and phagosome maturation are closely related to the body’s immune system.

**Table 13.** FUO samples IPA analysis results

| F | Ingenuity Canonical Pathways | P value | Molecules |
| --- | --- | --- | --- |
|  | STAT3 Pathway | 1.74E-04 | EGF, FGFR1, FGFR3, GHR, IL2RB, PDGFRB |
|  | SPINK1 Pancreatic Cancer Pathway | 3.47E-04 | CPM, CTSA, CTSB, KLK1 |
|  | IL-15 Production | 8.32E-04 | EPHB3, ERBB4, FGFR1, FGFR3, PDGFRB |
|  | Iron homeostasis signaling pathway | 1.32E-03 | CP, CREB3L3, EGF, FTL, PDGFRB |
|  | Hepatic Fibrosis / Hepatic Stellate Cell Activation | 6.03E-03 | COL15A1, EGF, FGFR1, PDGFRB, TIMP2 |
|  | Synaptogenesis Signaling Pathway | 1.12E-02 | CADM1, CDH19, CREB3L3, EPHB3, EPHB6, NECTIN3 |
|  | Phagosome Maturation | 1.29E-02 | CTSA, CTSB, CTSC, CTSS |
|  | PTEN Signaling | 1.32E-02 | FGFR1, FGFR3, GHR, PDGFRB |
|  | FGF Signaling | 1.45E-02 | CREB3L3, FGFR1, FGFR3 |
|  | Sperm Motility | 1.66E-02 | EPHB3, ERBB4, FGFR1, FGFR3, PDGFRB |
|  | Coagulation System | 1.91E-02 | KNG1, SERPINA5 |
|  | Intrinsic Prothrombin Activation Pathway | 2.57E-02 | KLK1, KNG1 |
|  | Regulation Of The Epithelial Mesenchymal Transition By Growth Factors Pathway | 2.95E-02 | EGF, FGFR1, FGFR3, PDGFRB |
|  | Neuroprotective Role of THOP1 in Alzheimer's Disease | 3.02E-02 | HGFAC, KLK1, KNG1 |
|  | Regulation of the Epithelial-Mesenchymal Transition Pathway | 3.02E-02 | EGF, FGFR1, FGFR3, PDGFRB |
|  | Ephrin Receptor Signaling | 3.31E-02 | CREB3L3, EGF, EPHB3, EPHB6 |
|  | Ceramide Degradation | 4.17E-02 | ASAH1 |
|  | CNTF Signaling | 4.57E-02 | CNTFR, LIFR |
|  | Adipogenesis pathway | 4.68E-02 | DLK1, FGFR1, FGFR3 |
|  | White Adipose Tissue Browning Pathway | 4.90E-02 | CREB3L3, FGFR1, FGFR3 |

We performed orthogonal partial least squares (OPLS–DA) analysis on the total protein. The OPLS–DA score is shown in Figure 1. The difference between the healthy group and the patient group was relatively large, which indicates that urine protein can reflect the disease state. At the same time, the intragroup differences of the patient group were more obvious, which means that the individual differences of the clinical samples were very large. In addition, the comparison of the IPA analysis results obtained by the single–person analysis and the group analysis shows that the group analysis will cover up the independence of individuals, which provides a theoretical basis for the significance of our single–person analysis.

**Figure 1.**
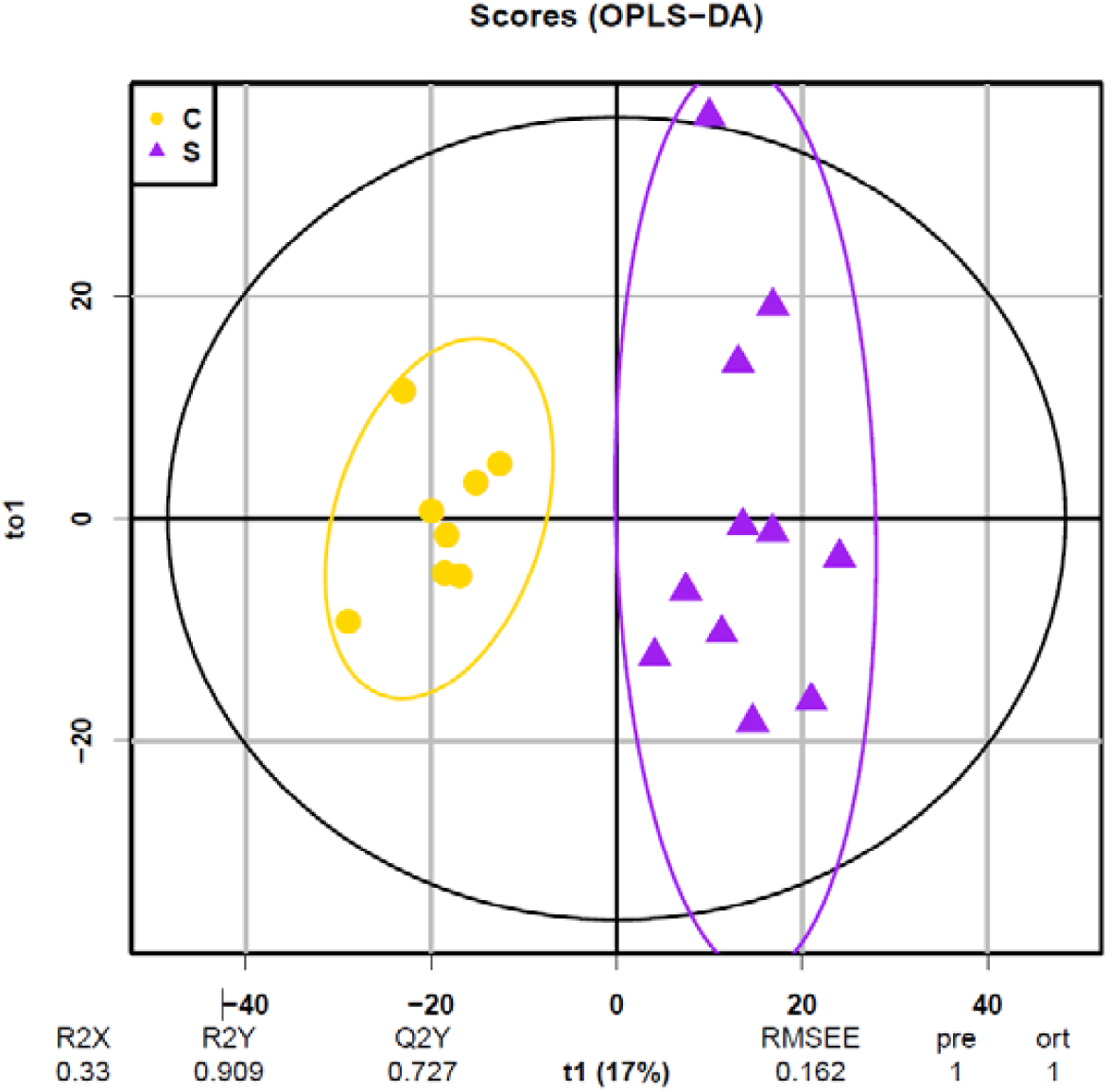
OPLS–DA analysis of identified urinary proteins.

### 3.3 Comparison of one–to–many analysis and group analysis

By comparing the results of the one–to–many analysis and the group analysis, we can observe that the results of the group analysis cannot represent the characteristics of any FUO patient. The group analysis only shows the same characteristics of the samples. In fact, each FUO patient was considered to exist independently. It is unreasonable to group them for analysis based on fever symptoms alone, and the results of 11 FUO samples were obviously different. It is worthwhile to mention that whether it is one–to–many analysis or group analysis, significant differences between FUO samples and healthy samples were found in urine, which provides a theoretical basis for clinical diagnosis of urine and, at the same time, reminds us that if there is a specific group of normal people who meet the control requirements for a specific patient as a control, it is more appropriate than using the same control for everyone, such as matching age, gender, medication history, and so on. More accurate results could be obtained if we could compare and analyse groups that match the indicators.

### 3.4 Parasite and virus search results

Parasite and virus databases downloaded form the UniPort website (https://www.uniprot.org/) were used to search for possible pathogenic factors. However, the microbes and viruses identified lacked sufficient credibility according to the current level of identification. Details are presented in Table S3.

## 4. Conclusions

In this study, we aimed to provide diagnostic evidence and clues for patients with fever of unknown origin (FUO) by using the one–to–many comparison label–free LC–MS/MS method. The differential proteins were screened for the analysis of biological pathways. Our results indicated that (1) the urine proteome can significantly distinguish between patients and healthy samples and (2) one–to–many comparisons can provide personalized clues for FUO patients. In fact, the individual differences in the clinical samples can easily be concealed by group analysis. One–to–many analysis can highlight the individualization of patients and it is also a way to explore clues to unknown disease patients in the future.

## Data Availability

All the data referred to in the manuscript are availability.

## CRediT authorship contribution statement

**Chenyang Zhao:** Conceptualization, Investigation, Writing-original draft, data curation, Visualization. **Lilong Wei:** Conceptualization, Resources, Funding acquisition. **Jing Wei:** Methodology, Investigation, formal analysis. **Youhe Gao:** Conceptualization, Writing-review & editing, Supervision, Funding acquisition.

## Acknowledgements

This work was supported by the National Key Research and Development Program of China (2018YFC0910202 and2 016YFC1306300); the Fundamental Research Funds for the Central Universities (2020KJZX002); the Beijing Natural Science Foundation (7172076); the Beijing Cooperative Construction Project (110651103); the Beijing Normal University (11100704). Key Clinical Specialty Project of Beijing (2020)

