## Supplemental Table 1 for "Using one–to–many urine proteome comparisons to provide clues for fever of unknown origin"

| F1 | | |
|---|---|---|
| **Protein Accessions** | **Fold Change** | **P value** |
| P02750 | 21.74 | 8.95E-09 |
| Q15293 | 17.17 | 3.30E-06 |
| P02763 | 8.01 | 5.50E-05 |
| P01911 | 7.89 | 8.39E-06 |
| P61769 | 6.57 | 7.68E-05 |
| P02741 | 5.98 | 1.14E-06 |
| P01903 | 5.50 | 1.92E-02 |
| Q14764 | 5.30 | 3.18E-05 |
| P02511 | 5.20 | 7.86E-05 |
| P01225 | 4.81 | 3.36E-03 |
| P00915 | 4.34 | 2.73E-02 |
| P01860 | 4.34 | 1.26E-02 |
| P02792 | 4.14 | 4.77E-03 |
| P01019 | 4.13 | 7.68E-05 |
| P27824 | 4.03 | 3.36E-03 |
| Q86YZ3 | 4.03 | 3.60E-02 |
| P18428 | 3.87 | 9.36E-05 |
| O60271 | 3.80 | 1.28E-03 |
| P62195 | 3.79 | 8.38E-05 |
| P50225 | 3.67 | 2.30E-02 |
| P02656 | 3.62 | 1.01E-02 |
| Q08830 | 3.61 | 5.51E-03 |
| Q14117 | 3.55 | 5.50E-05 |
| P09210 | 3.47 | 4.65E-04 |
| P14550 | 3.43 | 4.28E-04 |
| Q969H8 | 3.38 | 2.30E-02 |
| O14966 | 3.32 | 1.35E-03 |
| P09237 | 3.31 | 5.52E-03 |
| P01011 | 3.31 | 1.35E-03 |
| Q9NY33 | 3.27 | 3.65E-02 |
| P17858 | 3.20 | 5.44E-04 |
| P02748 | 3.06 | 3.24E-04 |
| Q9HCE6 | 3.03 | 8.62E-04 |
| P34913 | 2.98 | 4.86E-02 |
| Q7L5L3 | 2.97 | 1.27E-02 |
| Q5JTV8 | 2.96 | 1.06E-02 |
| P25705 | 2.91 | 4.39E-02 |

| | | |
|---|---|---|
| Q9UHI7 | 2.86 | 2.18E-03 |
| P28838 | 2.85 | 5.51E-03 |
| P23526 | 2.78 | 1.25E-03 |
| Q99972 | 2.70 | 1.37E-02 |
| P34896 | 2.69 | 1.17E-03 |
| P19652 | 2.67 | 2.53E-02 |
| P08571 | 2.63 | 2.30E-02 |
| O00754 | 2.63 | 1.35E-03 |
| Q9Y547 | 2.62 | 8.79E-03 |
| P50570 | 2.62 | 2.29E-03 |
| P25774 | 2.61 | 6.70E-03 |
| P50053 | 2.57 | 2.18E-03 |
| Q9BTY2 | 2.57 | 5.15E-03 |
| Q16851 | 2.57 | 2.13E-03 |
| P46926 | 2.57 | 1.26E-02 |
| P59998 | 2.54 | 9.37E-03 |
| Q8TDQ1 | 2.48 | 9.00E-03 |
| P22223 | 2.45 | 2.10E-02 |
| P20711 | 2.43 | 2.18E-02 |
| Q96DG6 | 2.41 | 5.15E-03 |
| Q9HD89 | 2.38 | 6.11E-03 |
| Q8NCC3 | 2.35 | 1.32E-02 |
| Q93099 | 2.34 | 1.90E-02 |
| P08253 | 2.33 | 4.25E-02 |
| Q96H15 | 2.32 | 2.93E-02 |
| P61026 | 2.27 | 5.75E-05 |
| Q6PK18 | 2.27 | 1.62E-02 |
| P07359 | 2.27 | 2.87E-02 |
| P21399 | 2.25 | 1.32E-02 |
| P09327 | 2.24 | 3.92E-02 |
| Q9P1F3 | 2.21 | 4.19E-02 |
| P78371 | 2.19 | 1.59E-02 |
| P20774 | 2.19 | 4.80E-02 |
| P51580 | 2.17 | 1.42E-02 |
| Q3LXA3 | 2.16 | 4.25E-02 |
| P02790 | 2.16 | 6.31E-03 |
| Q9Y617 | 2.14 | 9.98E-03 |
| Q6UX27 | 2.12 | 3.36E-03 |
| P11586 | 2.11 | 5.15E-03 |

| | | |
|---|---|---|
| Q8NHJ6 | 2.09 | 3.27E-02 |
| O75936 | 2.09 | 2.25E-02 |
| P08185 | 2.08 | 7.24E-03 |
| Q9NTK5 | 2.07 | 3.55E-02 |
| P0DMV8 | 2.07 | 3.65E-02 |
| Q9UBR2 | 2.05 | 1.67E-04 |
| P49368 | 2.04 | 1.75E-02 |
| P40925 | 2.00 | 3.78E-03 |
| P04066 | 2.00 | 9.00E-03 |
| Q8N386 | 0.50 | 4.13E-02 |
| P20933 | 0.50 | 3.31E-02 |
| Q6UX71 | 0.49 | 3.42E-02 |
| Q9UQB8 | 0.46 | 5.72E-03 |
| Q92859 | 0.46 | 3.60E-02 |
| P10586 | 0.45 | 9.00E-03 |
| Q99835 | 0.44 | 2.73E-02 |
| Q96JQ0 | 0.43 | 4.84E-02 |
| Q15223 | 0.40 | 2.36E-02 |
| Q9UBG0 | 0.40 | 1.92E-02 |
| P30086 | 0.39 | 4.84E-02 |
| P25940 | 0.34 | 4.19E-02 |
| Q13445 | 0.34 | 5.51E-03 |
| P19256 | 0.34 | 3.65E-02 |
| O75347 | 0.34 | 3.84E-02 |
| P13987 | 0.34 | 3.80E-02 |
| Q9HCU0 | 0.33 | 3.80E-02 |
| Q12864 | 0.32 | 4.84E-02 |
| P11047 | 0.32 | 1.85E-02 |
| Q9GZM5 | 0.30 | 4.58E-02 |
| Q9UNN8 | 0.25 | 2.52E-03 |
| O75339 | 0.23 | 3.92E-02 |
| Q9BWV1 | 0.18 | 2.27E-02 |

| F2 | | |
|---|---|---|
| **Protein Accessions** | **Fold Change** | **P value** |
| Q9Y547 | 15.88 | 1.00E-06 |
| P61769 | 15.13 | 3.84E-06 |
| O00244 | 14.16 | 5.23E-06 |
| Q9H299 | 13.13 | 1.75E-05 |
| Q15293 | 10.99 | 3.63E-03 |
| P00915 | 10.99 | 6.04E-05 |
| Q9NWV4 | 9.17 | 1.19E-05 |
| Q8NHJ6 | 8.79 | 1.98E-06 |
| P02750 | 8.28 | 1.32E-05 |
| O75368 | 8.23 | 3.84E-06 |
| P05534 | 8.18 | 3.84E-06 |
| P52272 | 8.11 | 5.59E-04 |
| Q9P1F3 | 6.57 | 7.90E-06 |
| P04080 | 6.54 | 3.14E-03 |
| P12318 | 6.27 | 1.47E-04 |
| P30508 | 6.15 | 4.28E-05 |
| Q96H15 | 6.09 | 1.32E-05 |
| O75531 | 5.83 | 1.32E-05 |
| P18465 | 5.66 | 1.32E-05 |
| P11684 | 5.48 | 2.44E-02 |
| O60234 | 5.08 | 1.12E-04 |
| Q92530 | 5.08 | 2.17E-02 |
| Q9Y6R1 | 4.99 | 2.11E-04 |
| Q9UHY7 | 4.98 | 2.97E-05 |
| Q9UBR2 | 4.84 | 1.32E-05 |
| Q9NRX4 | 4.77 | 4.72E-03 |
| P20774 | 4.65 | 1.16E-04 |
| P02763 | 4.57 | 5.34E-04 |
| Q86VP6 | 4.52 | 5.13E-04 |
| P02655 | 4.24 | 4.44E-04 |
| P36222 | 4.24 | 1.47E-04 |
| P16189 | 4.16 | 7.70E-04 |
| Q496F6 | 4.11 | 1.12E-04 |
| P25774 | 4.10 | 1.47E-04 |
| P01717 | 4.04 | 5.05E-05 |
| Q8TDQ1 | 4.03 | 1.31E-04 |
| P20333 | 3.99 | 4.83E-03 |

| | | |
|---|---|---|
| P35270 | 3.99 | 6.42E-03 |
| Q14019 | 3.83 | 7.70E-04 |
| Q9UHI7 | 3.83 | 1.12E-04 |
| P02656 | 3.83 | 3.80E-03 |
| Q93091 | 3.82 | 1.93E-03 |
| Q13740 | 3.79 | 6.87E-05 |
| P55786 | 3.71 | 1.09E-03 |
| Q9H3K6 | 3.70 | 4.80E-02 |
| Q13867 | 3.68 | 3.72E-04 |
| O43707 | 3.65 | 1.17E-03 |
| O95497 | 3.62 | 3.18E-03 |
| P04233 | 3.61 | 4.67E-03 |
| P01903 | 3.60 | 3.85E-02 |
| Q9Y279 | 3.51 | 9.18E-03 |
| O00754 | 3.50 | 7.98E-05 |
| P07737 | 3.47 | 1.50E-04 |
| P02792 | 3.46 | 1.00E-02 |
| P27824 | 3.43 | 6.38E-03 |
| P19652 | 3.42 | 3.18E-03 |
| O43491 | 3.41 | 3.80E-03 |
| Q8N357 | 3.38 | 1.01E-03 |
| Q07075 | 3.37 | 4.80E-02 |
| Q13113 | 3.33 | 2.48E-03 |
| Q03405 | 3.32 | 1.78E-03 |
| Q13421 | 3.23 | 2.77E-02 |
| Q9NZT2 | 3.21 | 2.01E-02 |
| Q15084 | 3.21 | 5.86E-03 |
| Q9BXP8 | 3.11 | 4.73E-03 |
| Q969H8 | 3.05 | 2.77E-02 |
| O15204 | 3.05 | 1.32E-02 |
| P06454 | 2.99 | 1.80E-02 |
| P05023 | 2.98 | 7.09E-04 |
| Q7L5L3 | 2.92 | 1.40E-02 |
| P07237 | 2.91 | 3.25E-02 |
| Q5JTV8 | 2.90 | 6.38E-03 |
| Q9UHI8 | 2.89 | 3.93E-02 |
| Q96C19 | 2.88 | 1.90E-02 |
| Q15274 | 2.87 | 2.32E-02 |
| Q6UXN8 | 2.83 | 2.32E-03 |

| | | |
|---|---|---|
| P36980 | 2.80 | 1.39E-02 |
| Q9NUM4 | 2.80 | 1.36E-02 |
| O75022 | 2.79 | 4.83E-03 |
| Q8NCC3 | 2.78 | 2.32E-03 |
| P02511 | 2.77 | 6.26E-03 |
| Q9BTM9 | 2.76 | 4.86E-03 |
| Q15116 | 2.75 | 2.72E-04 |
| P35754 | 2.74 | 3.85E-02 |
| P01019 | 2.73 | 1.01E-03 |
| Q92542 | 2.69 | 3.12E-02 |
| P08729 | 2.66 | 1.68E-03 |
| A0PJK1 | 2.61 | 2.29E-02 |
| Q96IU4 | 2.58 | 2.32E-02 |
| Q9Y6X5 | 2.57 | 6.86E-03 |
| Q15121 | 2.54 | 2.96E-02 |
| P53634 | 2.52 | 3.80E-04 |
| P02671 | 2.51 | 2.06E-02 |
| Q9H665 | 2.50 | 3.07E-03 |
| P01011 | 2.50 | 8.13E-03 |
| O00182 | 2.49 | 2.34E-03 |
| Q03167 | 2.49 | 1.08E-02 |
| P07686 | 2.47 | 3.07E-03 |
| P22223 | 2.47 | 1.38E-02 |
| P08571 | 2.46 | 2.50E-02 |
| P07858 | 2.44 | 5.30E-03 |
| P12111 | 2.42 | 4.33E-02 |
| P30043 | 2.41 | 3.32E-02 |
| Q695T7 | 2.37 | 3.54E-02 |
| P00746 | 2.35 | 3.25E-02 |
| P78310 | 2.35 | 3.87E-02 |
| P07900 | 2.34 | 4.80E-02 |
| P07339 | 2.34 | 1.17E-03 |
| P01700 | 2.32 | 2.32E-02 |
| P62942 | 2.32 | 2.32E-02 |
| P26368 | 2.30 | 2.29E-02 |
| P67809 | 2.28 | 5.00E-02 |
| P0CG47 | 2.27 | 2.77E-02 |
| Q9UHG2 | 2.26 | 1.80E-02 |
| P29401 | 2.25 | 3.06E-02 |

| | | |
|---|---|---|
| Q9Y5Y7 | 2.24 | 4.33E-02 |
| P00740 | 2.22 | 3.86E-02 |
| Q86VB7 | 2.19 | 1.14E-03 |
| P09668 | 2.18 | 9.18E-03 |
| P04216 | 2.18 | 3.44E-04 |
| Q6GTX8 | 2.17 | 1.78E-03 |
| P00450 | 2.17 | 3.63E-02 |
| Q1EHB4 | 2.17 | 3.85E-02 |
| Q5T2W1 | 2.15 | 4.83E-03 |
| O75608 | 2.14 | 3.86E-03 |
| P04066 | 2.13 | 3.44E-03 |
| P01589 | 2.12 | 3.32E-02 |
| Q7Z6A9 | 2.10 | 3.26E-02 |
| P21333 | 2.09 | 1.90E-02 |
| P06865 | 2.05 | 3.99E-03 |
| P0DOX8 | 2.05 | 3.34E-02 |
| Q9Y5E6 | 0.47 | 4.33E-02 |
| Q9NZ53 | 0.46 | 1.90E-02 |
| O60494 | 0.45 | 2.77E-02 |
| Q9UNN8 | 0.45 | 7.89E-03 |
| O75882 | 0.45 | 4.07E-02 |
| Q6UXB4 | 0.43 | 7.40E-03 |
| Q8TB96 | 0.42 | 2.57E-02 |
| Q7KYR7 | 0.42 | 2.96E-02 |
| Q9HCM3 | 0.42 | 3.52E-02 |
| P10586 | 0.41 | 4.72E-03 |
| Q6UX71 | 0.41 | 1.31E-02 |
| Q9UBG0 | 0.40 | 1.85E-02 |
| Q5SZK8 | 0.39 | 3.26E-02 |
| Q9NQS3 | 0.39 | 1.38E-02 |
| Q9HBR0 | 0.38 | 1.72E-02 |
| P13598 | 0.38 | 3.11E-02 |
| Q96JQ0 | 0.37 | 2.50E-02 |
| Q92673 | 0.36 | 4.23E-02 |
| P24855 | 0.36 | 3.44E-02 |
| Q13445 | 0.35 | 3.63E-03 |
| O75144 | 0.35 | 2.29E-02 |
| P51570 | 0.35 | 1.70E-02 |
| Q99835 | 0.35 | 1.10E-02 |

| | | |
|---|---|---|
| P41217 | 0.34 | 4.46E-02 |
| P15151 | 0.34 | 3.74E-02 |
| Q9UN37 | 0.33 | 3.87E-02 |
| Q9UK41 | 0.33 | 3.86E-02 |
| P04180 | 0.33 | 1.90E-02 |
| Q86YT9 | 0.32 | 4.07E-02 |
| Q92859 | 0.32 | 9.18E-03 |
| Q96MU8 | 0.31 | 4.80E-02 |
| P0C7U0 | 0.30 | 4.21E-02 |
| P31025 | 0.30 | 2.35E-02 |
| Q6UXG3 | 0.29 | 3.14E-02 |
| Q86VN1 | 0.29 | 1.88E-02 |
| Q9H1U4 | 0.28 | 4.80E-02 |
| Q9BWV1 | 0.28 | 2.77E-02 |
| Q9HCU0 | 0.27 | 2.15E-02 |
| P11047 | 0.25 | 7.84E-03 |
| Q8NDA2 | 0.23 | 4.80E-02 |
| Q8IUL8 | 0.18 | 2.29E-02 |
| P12273 | 0.12 | 4.07E-02 |
| P55017 | 0.11 | 2.35E-02 |

| F3 | | |
|---|---|---|
| **Protein Accessions** | **Fold Change** | **P value** |
| P04279 | 34.12 | 9.11E-09 |
| Q02383 | 27.48 | 5.32E-08 |
| Q15293 | 7.67 | 1.68E-04 |
| P14555 | 6.70 | 1.68E-04 |
| Q6W4X9 | 6.44 | 1.48E-03 |
| P02763 | 5.67 | 2.80E-04 |
| P02750 | 5.51 | 3.76E-04 |
| O76070 | 4.54 | 1.79E-03 |
| P01721 | 3.95 | 4.56E-03 |
| P02679 | 3.93 | 4.00E-02 |
| P01024 | 3.79 | 1.98E-03 |
| Q969H8 | 3.76 | 1.53E-02 |
| Q13113 | 3.68 | 8.07E-04 |
| P09237 | 3.67 | 4.01E-03 |
| P01225 | 3.60 | 2.67E-02 |
| P25311 | 3.51 | 3.54E-02 |
| Q86VP6 | 3.37 | 2.45E-03 |
| P02776 | 3.37 | 3.14E-03 |
| A0A075B6K4 | 3.25 | 4.78E-02 |
| Q8N114 | 3.02 | 4.78E-02 |
| Q9P1F3 | 3.00 | 6.17E-03 |
| Q9Y6R1 | 2.99 | 1.93E-02 |
| O75368 | 2.88 | 1.20E-02 |
| Q9UBR2 | 2.88 | 7.06E-06 |
| Q9UHI7 | 2.85 | 2.65E-03 |
| P19652 | 2.81 | 2.31E-02 |
| A0PJK1 | 2.80 | 4.01E-02 |
| P09543 | 2.77 | 1.48E-03 |
| Q8TDQ1 | 2.69 | 6.17E-03 |
| Q9BYE9 | 2.65 | 7.77E-03 |
| P02790 | 2.46 | 2.74E-03 |
| P01699 | 2.40 | 8.07E-03 |
| P02748 | 2.38 | 4.01E-03 |
| P50897 | 2.37 | 5.36E-04 |
| P08253 | 2.36 | 4.78E-02 |
| P07339 | 2.32 | 2.74E-03 |
| P08603 | 2.32 | 1.67E-03 |

| | | |
|---|---|---|
| P18428 | 2.30 | 1.14E-02 |
| Q13510 | 2.27 | 4.01E-03 |
| P07996 | 2.24 | 2.45E-03 |
| P01717 | 2.19 | 1.20E-02 |
| P09668 | 2.18 | 1.93E-02 |
| P07686 | 2.14 | 1.93E-02 |
| P02741 | 2.11 | 7.20E-03 |
| Q9HAT2 | 2.10 | 1.93E-02 |
| Q07507 | 2.09 | 1.67E-03 |
| O14786 | 2.06 | 3.36E-02 |
| P53634 | 2.06 | 6.11E-03 |
| P01344 | 2.06 | 4.78E-02 |
| Q92743 | 2.04 | 1.81E-02 |
| Q13445 | 0.49 | 1.93E-02 |
| P43121 | 0.42 | 1.99E-02 |
| P11047 | 0.42 | 4.92E-02 |
| Q92520 | 0.40 | 4.78E-02 |
| Q9NU53 | 0.37 | 4.78E-02 |
| Q9UNN8 | 0.36 | 7.77E-03 |
| P12955 | 0.35 | 4.78E-02 |
| Q92859 | 0.31 | 1.71E-02 |
| Q14118 | 0.29 | 4.13E-02 |
| Q9UJJ9 | 0.26 | 4.78E-02 |
| Q8N386 | 0.25 | 1.01E-02 |
| Q9H0X4 | 0.25 | 4.25E-02 |
| Q14117 | 0.21 | 3.27E-02 |
| P45877 | 0.15 | 3.24E-02 |
| Q8TB96 | 0.13 | 8.03E-03 |

| F4 | | |
| --- | --- | --- |
| **Protein Accessions** | **Fold Change** | **P value** |
| P02792 | 14.92 | 1.18E-05 |
| Q14764 | 11.70 | 8.23E-06 |
| P02679 | 11.42 | 7.61E-05 |
| P02675 | 9.76 | 5.93E-05 |
| P01903 | 8.93 | 9.91E-05 |
| Q07075 | 8.09 | 1.19E-04 |
| O60271 | 7.96 | 8.23E-06 |
| Q9HCE6 | 7.80 | 2.25E-06 |
| P01023 | 7.21 | 5.01E-04 |
| P01911 | 6.93 | 1.91E-02 |
| Q14204 | 6.73 | 4.00E-04 |
| P50570 | 6.72 | 2.69E-06 |
| Q9ULZ3 | 6.71 | 1.98E-03 |
| P00966 | 6.66 | 4.14E-06 |
| P17858 | 6.37 | 2.69E-06 |
| P06727 | 6.30 | 2.31E-04 |
| Q9Y6X5 | 6.28 | 1.53E-04 |
| P02794 | 6.07 | 1.72E-03 |
| Q07837 | 5.94 | 8.23E-06 |
| Q16851 | 5.84 | 8.23E-06 |
| Q9BR76 | 5.77 | 6.57E-04 |
| P34913 | 5.72 | 1.15E-03 |
| Q6UWR7 | 5.63 | 2.63E-04 |
| P00439 | 5.58 | 2.51E-04 |
| O95954 | 5.24 | 4.51E-05 |
| P21399 | 5.17 | 7.01E-05 |
| O75891 | 5.16 | 4.14E-06 |
| Q3LXA3 | 5.16 | 1.17E-04 |
| Q9H2A2 | 5.12 | 6.43E-05 |
| O95154 | 5.12 | 1.93E-04 |
| Q9Y2T3 | 5.06 | 2.38E-04 |
| P11586 | 5.00 | 2.49E-05 |
| P02511 | 4.99 | 8.73E-05 |
| Q9UHL4 | 4.97 | 2.70E-04 |
| P07900 | 4.93 | 8.90E-05 |
| P40925 | 4.89 | 8.23E-06 |
| Q15274 | 4.88 | 2.86E-04 |

| | | |
|---|---|---|
| P28838 | 4.87 | 1.09E-04 |
| Q9Y617 | 4.87 | 1.61E-05 |
| Q15293 | 4.84 | 2.52E-02 |
| P62195 | 4.81 | 8.77E-05 |
| Q9HB90 | 4.77 | 1.10E-03 |
| O14841 | 4.75 | 7.57E-03 |
| P61026 | 4.71 | 9.55E-06 |
| Q99832 | 4.70 | 8.90E-05 |
| Q7L5L3 | 4.63 | 1.87E-04 |
| Q06495 | 4.58 | 2.11E-03 |
| Q9NUM4 | 4.57 | 1.87E-04 |
| P49368 | 4.56 | 8.87E-05 |
| Q96DG6 | 4.54 | 1.61E-05 |
| O75936 | 4.51 | 5.44E-05 |
| P02656 | 4.48 | 5.53E-04 |
| P04114 | 4.43 | 4.84E-04 |
| P08133 | 4.41 | 1.91E-04 |
| Q9UBR1 | 4.38 | 7.32E-04 |
| P02763 | 4.36 | 3.50E-04 |
| Q9NVS9 | 4.35 | 3.82E-04 |
| Q5R3I4 | 4.30 | 1.50E-05 |
| P02647 | 4.29 | 2.98E-02 |
| P02750 | 4.24 | 2.20E-04 |
| Q53T59 | 4.24 | 5.17E-04 |
| P14550 | 4.22 | 4.27E-05 |
| P05062 | 4.18 | 2.95E-05 |
| Q92747 | 4.16 | 1.86E-04 |
| P50053 | 4.16 | 8.64E-03 |
| Q96KP4 | 4.13 | 2.55E-04 |
| P61160 | 4.10 | 6.02E-05 |
| P08237 | 4.09 | 3.37E-04 |
| P02652 | 4.08 | 1.61E-02 |
| Q14117 | 4.05 | 8.23E-06 |
| P32189 | 4.03 | 8.26E-04 |
| P53634 | 4.01 | 8.23E-06 |
| P28332 | 4.01 | 1.09E-04 |
| P49773 | 3.98 | 4.60E-03 |
| P17980 | 3.97 | 7.50E-04 |
| Q9P1F3 | 3.93 | 1.37E-04 |

| | | |
|---|---|---|
| Q6ZQN7 | 3.93 | 4.82E-05 |
| Q9Y5S2 | 3.92 | 1.66E-03 |
| Q93050 | 3.92 | 2.95E-05 |
| P30039 | 3.91 | 3.29E-04 |
| P09210 | 3.91 | 7.01E-05 |
| P62333 | 3.90 | 9.08E-03 |
| Q92542 | 3.87 | 1.04E-03 |
| O43776 | 3.84 | 1.43E-03 |
| Q93099 | 3.84 | 2.63E-04 |
| P00746 | 3.82 | 8.14E-04 |
| Q93091 | 3.80 | 1.47E-03 |
| Q13200 | 3.79 | 3.15E-03 |
| P21695 | 3.78 | 3.28E-04 |
| Q8N2U0 | 3.77 | 1.10E-02 |
| Q9Y2S2 | 3.77 | 1.87E-04 |
| O43488 | 3.71 | 4.82E-05 |
| P51606 | 3.67 | 4.82E-05 |
| P68133 | 3.67 | 1.61E-05 |
| P19823 | 3.66 | 2.09E-02 |
| P02654 | 3.65 | 3.82E-04 |
| O00338 | 3.64 | 3.52E-03 |
| P34896 | 3.62 | 3.63E-05 |
| P09327 | 3.61 | 6.58E-04 |
| P12955 | 3.60 | 1.61E-05 |
| O14966 | 3.56 | 1.91E-04 |
| O15144 | 3.56 | 1.91E-04 |
| Q13277 | 3.56 | 1.48E-05 |
| P78371 | 3.53 | 6.58E-04 |
| P02655 | 3.51 | 8.86E-03 |
| Q96FL8 | 3.51 | 1.68E-03 |
| Q14019 | 3.48 | 1.08E-03 |
| B5ME19 | 3.46 | 5.97E-03 |
| P51580 | 3.46 | 8.73E-05 |
| P35542 | 3.45 | 3.24E-02 |
| Q14847 | 3.45 | 8.96E-05 |
| O75264 | 3.41 | 8.90E-05 |
| Q9Y547 | 3.41 | 8.50E-04 |
| P11766 | 3.37 | 5.81E-04 |
| P60953 | 3.37 | 5.17E-04 |

| | | |
|---|---|---|
| Q15833 | 3.37 | 8.96E-05 |
| P61158 | 3.37 | 1.87E-04 |
| Q86VP6 | 3.36 | 3.57E-04 |
| P07858 | 3.34 | 1.87E-04 |
| Q06278 | 3.33 | 2.04E-03 |
| P24534 | 3.30 | 1.05E-02 |
| Q5T2W1 | 3.29 | 7.90E-05 |
| O00754 | 3.25 | 8.11E-05 |
| P45381 | 3.25 | 3.39E-03 |
| Q9UHI7 | 3.24 | 2.66E-04 |
| P09668 | 3.23 | 1.55E-04 |
| P17900 | 3.22 | 1.88E-02 |
| P19652 | 3.22 | 2.24E-03 |
| P27105 | 3.22 | 1.16E-03 |
| Q9UJ68 | 3.21 | 1.91E-04 |
| Q96BW5 | 3.20 | 1.10E-03 |
| Q96FV2 | 3.20 | 1.87E-04 |
| P34932 | 3.17 | 1.05E-04 |
| P20711 | 3.16 | 7.62E-04 |
| P46926 | 3.16 | 6.75E-04 |
| P48643 | 3.16 | 5.05E-03 |
| P50502 | 3.14 | 1.87E-04 |
| P63261 | 3.12 | 1.35E-04 |
| Q9H2M3 | 3.07 | 9.81E-03 |
| Q9BUT1 | 3.06 | 1.54E-03 |
| P13798 | 3.01 | 5.53E-04 |
| Q9UHY7 | 3.00 | 1.11E-03 |
| P13716 | 2.99 | 4.25E-03 |
| Q93088 | 2.99 | 9.75E-03 |
| P17050 | 2.98 | 1.69E-04 |
| Q9UM54 | 2.98 | 3.46E-02 |
| P25311 | 2.96 | 2.47E-02 |
| O75083 | 2.95 | 8.35E-04 |
| P30711 | 2.95 | 3.15E-02 |
| Q03154 | 2.94 | 2.44E-03 |
| O00244 | 2.93 | 4.35E-02 |
| Q9UBR2 | 2.93 | 8.23E-06 |
| P55072 | 2.92 | 4.64E-03 |
| P17405 | 2.92 | 7.01E-05 |

| | | |
|---|---|---|
| P50990 | 2.92 | 1.99E-03 |
| Q00610 | 2.92 | 7.40E-03 |
| Q9NWV4 | 2.90 | 1.28E-02 |
| O43707 | 2.88 | 2.83E-03 |
| P31153 | 2.88 | 1.37E-03 |
| P23526 | 2.87 | 1.87E-04 |
| Q01518 | 2.87 | 5.84E-03 |
| P59998 | 2.86 | 1.23E-03 |
| P45974 | 2.85 | 4.82E-05 |
| P49189 | 2.85 | 6.34E-04 |
| Q9H4A4 | 2.83 | 4.01E-02 |
| P09525 | 2.82 | 1.99E-03 |
| Q9UBQ7 | 2.80 | 1.35E-03 |
| Q9UJU6 | 2.78 | 9.45E-03 |
| SWISS-PROT:P10096 | 2.78 | 1.93E-04 |
| O15143 | 2.77 | 4.60E-02 |
| Q9BPX5 | 2.77 | 5.07E-03 |
| A0A0C4DH24 | 2.76 | 2.66E-02 |
| P09467 | 2.75 | 1.02E-02 |
| Q14914 | 2.75 | 2.72E-03 |
| Q9NTX5 | 2.73 | 1.35E-03 |
| P07148 | 2.73 | 3.39E-03 |
| Q7L9L4 | 2.73 | 6.64E-03 |
| P25774 | 2.72 | 1.40E-03 |
| Q9NTK5 | 2.71 | 1.95E-03 |
| P10636 | 2.71 | 5.09E-03 |
| P08473 | 2.70 | 1.28E-02 |
| P27487 | 2.70 | 8.14E-04 |
| P10768 | 2.70 | 3.62E-04 |
| P51570 | 2.70 | 1.53E-04 |
| P53396 | 2.70 | 3.62E-03 |
| Q6XQN6 | 2.69 | 3.99E-03 |
| P02747 | 2.68 | 2.48E-03 |
| O43681 | 2.67 | 1.87E-04 |
| Q9Y6R1 | 2.65 | 1.28E-02 |
| O75348 | 2.64 | 2.18E-03 |
| P27824 | 2.64 | 1.86E-02 |
| P52758 | 2.64 | 2.92E-02 |
| P54803 | 2.62 | 4.19E-04 |

| | | |
|---|---|---|
| Q13183 | 2.62 | 7.50E-04 |
| Q9UI12 | 2.62 | 1.90E-03 |
| Q16186 | 2.59 | 1.50E-02 |
| Q8WWA0 | 2.59 | 4.35E-02 |
| P08183 | 2.57 | 1.17E-03 |
| P54793 | 2.56 | 1.84E-02 |
| A0PJK1 | 2.53 | 6.15E-03 |
| P35558 | 2.51 | 4.46E-03 |
| Q06323 | 2.51 | 3.61E-03 |
| P13796 | 2.51 | 3.73E-02 |
| P60900 | 2.50 | 9.42E-03 |
| Q9Y281 | 2.50 | 6.15E-03 |
| O15511 | 2.47 | 1.87E-02 |
| P04406 | 2.46 | 9.39E-04 |
| Q1EHB4 | 2.46 | 1.30E-02 |
| P35998 | 2.45 | 7.43E-03 |
| Q99536 | 2.45 | 9.26E-03 |
| P13639 | 2.44 | 1.80E-02 |
| P00338 | 2.43 | 7.37E-03 |
| P23528 | 2.42 | 1.52E-03 |
| P67809 | 2.41 | 4.87E-02 |
| Q13510 | 2.40 | 4.19E-04 |
| Q8NCW5 | 2.39 | 4.50E-03 |
| P00441 | 2.39 | 2.44E-03 |
| O96009 | 2.39 | 7.77E-03 |
| Q9BTY2 | 2.39 | 2.51E-03 |
| Q9NS93 | 2.38 | 3.23E-02 |
| P00558 | 2.38 | 3.19E-04 |
| P36871 | 2.38 | 1.97E-02 |
| Q08257 | 2.36 | 6.24E-03 |
| O00401 | 2.36 | 8.76E-03 |
| A0A0C4DH38 | 2.35 | 7.77E-03 |
| P30085 | 2.35 | 1.68E-02 |
| A6NIZ1 | 2.34 | 9.06E-03 |
| Q9NVJ2 | 2.33 | 3.74E-02 |
| P61981 | 2.31 | 3.48E-02 |
| O00115 | 2.30 | 2.34E-03 |
| O14818 | 2.29 | 2.45E-03 |
| Q6UWV6 | 2.29 | 5.36E-03 |

| | | |
|---|---|---|
| P32754 | 2.29 | 1.47E-02 |
| Q9H008 | 2.28 | 4.13E-04 |
| P47755 | 2.27 | 8.10E-03 |
| Q9Y646 | 2.27 | 7.77E-03 |
| P61006 | 2.25 | 2.07E-02 |
| P10619 | 2.25 | 3.61E-03 |
| P00352 | 2.24 | 9.93E-03 |
| Q08830 | 2.24 | 3.88E-02 |
| Q86X76 | 2.24 | 5.85E-03 |
| Q04917 | 2.23 | 3.74E-02 |
| Q13228 | 2.23 | 4.67E-03 |
| Q9C0H2 | 2.22 | 2.80E-02 |
| Q7Z3F1 | 2.22 | 4.93E-02 |
| P01024 | 2.21 | 2.23E-02 |
| P60981 | 2.21 | 7.92E-03 |
| Q96C19 | 2.21 | 4.69E-02 |
| P31946 | 2.21 | 5.66E-03 |
| Q9NQR4 | 2.20 | 1.13E-02 |
| P01717 | 2.20 | 3.85E-03 |
| Q13113 | 2.20 | 1.78E-02 |
| Q14194 | 2.19 | 3.10E-03 |
| P00491 | 2.19 | 1.78E-02 |
| P30153 | 2.19 | 7.17E-03 |
| P06744 | 2.18 | 2.04E-03 |
| P48637 | 2.17 | 9.94E-03 |
| Q9P2T1 | 2.17 | 1.90E-03 |
| P08603 | 2.15 | 6.29E-04 |
| P47756 | 2.15 | 3.62E-03 |
| Q495M3 | 2.15 | 5.66E-03 |
| O15145 | 2.15 | 2.36E-02 |
| P08729 | 2.14 | 7.77E-03 |
| Q8N357 | 2.14 | 3.50E-02 |
| O94760 | 2.12 | 6.48E-04 |
| Q15493 | 2.12 | 5.84E-03 |
| P62942 | 2.11 | 2.33E-02 |
| P82980 | 2.11 | 3.94E-03 |
| P51688 | 2.10 | 4.32E-02 |
| P06733 | 2.10 | 9.93E-03 |
| P50395 | 2.10 | 3.62E-03 |

| | | |
|---|---|---|
| P25789 | 2.09 | 1.38E-02 |
| P29401 | 2.08 | 3.76E-02 |
| Q9NR99 | 2.08 | 2.61E-02 |
| Q96LD4 | 2.08 | 2.91E-04 |
| Q8WWB7 | 2.05 | 5.78E-03 |
| P78417 | 2.03 | 6.93E-03 |
| P68104 | 2.02 | 1.75E-02 |
| P11142 | 2.01 | 1.04E-02 |
| O75309 | 0.50 | 3.18E-02 |
| O75594 | 0.49 | 6.39E-04 |
| Q08722 | 0.48 | 2.56E-02 |
| Q9UQB8 | 0.48 | 2.18E-03 |
| P10586 | 0.48 | 4.07E-03 |
| Q9UJJ9 | 0.48 | 4.62E-02 |
| O14498 | 0.48 | 4.24E-02 |
| Q9NPH3 | 0.48 | 4.35E-02 |
| P28827 | 0.47 | 2.53E-02 |
| Q9NZ53 | 0.47 | 9.94E-03 |
| Q6UX71 | 0.47 | 9.84E-03 |
| O60888 | 0.47 | 9.71E-03 |
| P07942 | 0.46 | 1.35E-02 |
| O94910 | 0.46 | 4.35E-02 |
| P55957 | 0.45 | 3.74E-02 |
| Q15223 | 0.45 | 1.22E-02 |
| Q99835 | 0.45 | 1.04E-02 |
| O43921 | 0.45 | 4.36E-02 |
| Q8NBS9 | 0.45 | 4.61E-02 |
| Q92520 | 0.44 | 1.78E-02 |
| P00734 | 0.44 | 4.89E-02 |
| P54760 | 0.44 | 3.47E-02 |
| O75882 | 0.44 | 1.96E-02 |
| P21709 | 0.43 | 2.82E-02 |
| Q06481 | 0.43 | 1.09E-02 |
| Q9GZM5 | 0.43 | 3.76E-02 |
| Q9H741 | 0.43 | 4.14E-02 |
| P55899 | 0.43 | 2.40E-02 |
| Q05707 | 0.42 | 3.07E-02 |
| P20138 | 0.42 | 1.82E-02 |
| Q13443 | 0.42 | 4.10E-02 |

| | | |
|---|---|---|
| O43291 | 0.42 | 4.35E-02 |
| P25940 | 0.42 | 2.53E-02 |
| Q7LBR1 | 0.41 | 4.54E-02 |
| P78552 | 0.41 | 3.24E-02 |
| Q9HBR0 | 0.41 | 1.13E-02 |
| P19022 | 0.41 | 2.96E-02 |
| Q9Y4D7 | 0.40 | 2.45E-02 |
| P13611 | 0.40 | 4.22E-02 |
| Q06418 | 0.40 | 2.30E-02 |
| Q14118 | 0.40 | 2.06E-02 |
| P02741 | 0.40 | 2.52E-02 |
| P13591 | 0.40 | 3.90E-02 |
| P16234 | 0.40 | 3.54E-02 |
| P45877 | 0.40 | 3.20E-02 |
| P10646 | 0.40 | 3.03E-02 |
| Q08334 | 0.40 | 3.76E-02 |
| Q96J84 | 0.40 | 4.62E-02 |
| P26992 | 0.39 | 4.45E-02 |
| Q9NPR2 | 0.39 | 4.80E-03 |
| P22105 | 0.39 | 4.09E-02 |
| O95490 | 0.39 | 9.80E-03 |
| P40189 | 0.39 | 2.47E-02 |
| P04275 | 0.39 | 3.74E-02 |
| P08123 | 0.39 | 4.95E-02 |
| Q9P2B2 | 0.38 | 2.90E-02 |
| Q04721 | 0.38 | 4.98E-02 |
| P13598 | 0.38 | 1.59E-02 |
| Q8TDQ0 | 0.38 | 3.56E-02 |
| Q13445 | 0.38 | 2.03E-03 |
| Q9NY25 | 0.38 | 4.10E-02 |
| O75787 | 0.38 | 2.82E-02 |
| Q12913 | 0.37 | 4.20E-02 |
| Q6P531 | 0.37 | 2.59E-02 |
| P05067 | 0.37 | 4.52E-02 |
| Q99523 | 0.36 | 4.88E-02 |
| P52798 | 0.36 | 2.60E-02 |
| Q6UXB4 | 0.36 | 1.90E-03 |
| P02462 | 0.36 | 4.89E-02 |
| P09619 | 0.36 | 5.84E-03 |

| | | |
|---|---|---|
| P0DMQ5 | 0.36 | 2.68E-02 |
| P20827 | 0.36 | 1.28E-02 |
| P19256 | 0.36 | 1.49E-02 |
| P11047 | 0.36 | 8.10E-03 |
| Q16832 | 0.35 | 2.61E-02 |
| Q9UKU6 | 0.35 | 1.93E-02 |
| Q969Z4 | 0.35 | 4.45E-02 |
| Q8TB96 | 0.35 | 7.70E-03 |
| Q8IZA0 | 0.35 | 2.58E-02 |
| Q9UBG0 | 0.35 | 4.18E-03 |
| Q68D85 | 0.34 | 1.82E-02 |
| Q15116 | 0.34 | 1.78E-02 |
| P16070 | 0.34 | 1.84E-02 |
| Q9UBP0 | 0.34 | 1.78E-02 |
| Q96RW7 | 0.34 | 3.59E-02 |
| Q9H9H4 | 0.33 | 4.35E-02 |
| Q96PD5 | 0.33 | 4.67E-02 |
| Q5ZPR3 | 0.33 | 2.68E-02 |
| O75339 | 0.32 | 2.46E-02 |
| P12259 | 0.32 | 1.87E-02 |
| P23284 | 0.32 | 2.30E-02 |
| Q9NZP8 | 0.32 | 2.39E-02 |
| O75084 | 0.32 | 9.24E-03 |
| Q5JXA9 | 0.32 | 1.80E-02 |
| Q13591 | 0.32 | 4.93E-02 |
| Q92673 | 0.32 | 1.78E-02 |
| Q9P0T7 | 0.31 | 3.98E-02 |
| Q9NPF0 | 0.31 | 2.29E-02 |
| Q08345 | 0.31 | 2.24E-02 |
| Q5SZK8 | 0.31 | 9.93E-03 |
| P41217 | 0.31 | 1.97E-02 |
| Q12860 | 0.31 | 4.43E-02 |
| Q8NDA2 | 0.31 | 3.87E-02 |
| O75144 | 0.31 | 9.75E-03 |
| Q96JQ0 | 0.31 | 8.30E-03 |
| P04180 | 0.31 | 7.77E-03 |
| O43278 | 0.31 | 9.64E-03 |
| Q9HCN6 | 0.31 | 4.35E-02 |
| Q15262 | 0.30 | 1.21E-02 |

| | | |
|---|---|---|
| Q8NC42 | 0.30 | 1.50E-02 |
| O43556 | 0.30 | 1.72E-02 |
| P15151 | 0.30 | 1.50E-02 |
| Q9BY67 | 0.30 | 4.67E-02 |
| P15291 | 0.29 | 1.13E-02 |
| Q7KYR7 | 0.29 | 5.85E-03 |
| P14784 | 0.29 | 2.40E-02 |
| Q9UN74 | 0.29 | 4.10E-02 |
| P08572 | 0.28 | 2.95E-02 |
| Q7Z7G0 | 0.28 | 3.51E-02 |
| Q9UNN8 | 0.28 | 8.26E-04 |
| Q9ULK6 | 0.28 | 2.36E-02 |
| Q9NQS3 | 0.28 | 2.61E-03 |
| Q9UEF7 | 0.28 | 4.35E-02 |
| Q6V0I7 | 0.27 | 3.66E-02 |
| Q9Y653 | 0.27 | 2.67E-02 |
| Q9HCM3 | 0.27 | 6.54E-03 |
| P26927 | 0.27 | 2.61E-02 |
| Q9NR34 | 0.26 | 3.76E-02 |
| P24855 | 0.26 | 9.71E-03 |
| Q92859 | 0.26 | 2.62E-03 |
| Q86YT9 | 0.25 | 1.47E-02 |
| Q9HCU0 | 0.25 | 8.64E-03 |
| Q9UBQ6 | 0.25 | 2.69E-02 |
| P0C7U0 | 0.24 | 1.68E-02 |
| P09603 | 0.24 | 3.76E-02 |
| Q8TBP5 | 0.24 | 2.68E-02 |
| Q9H0X4 | 0.24 | 9.80E-03 |
| SWISS-PROT:P19012 | 0.24 | 3.13E-02 |
| P58499 | 0.24 | 1.65E-02 |
| Q6UXG3 | 0.24 | 1.14E-02 |
| P98095 | 0.23 | 4.93E-02 |
| Q9Y5E6 | 0.23 | 4.18E-03 |
| P01042 | 0.23 | 3.85E-02 |
| Q9Y639 | 0.22 | 2.30E-02 |
| P16035 | 0.22 | 1.50E-02 |
| Q9HBB8 | 0.22 | 2.61E-02 |
| P35916 | 0.21 | 1.68E-02 |
| Q96MU8 | 0.21 | 1.50E-02 |

| | | |
|---|---|---|
| Q9Y2G1 | 0.21 | 1.09E-02 |
| P80370 | 0.19 | 3.12E-02 |
| Q8IUL8 | 0.18 | 1.09E-02 |
| Q86UX2 | 0.18 | 2.68E-02 |
| P07451 | 0.17 | 4.15E-02 |
| Q9H1U4 | 0.17 | 1.09E-02 |
| P12109 | 0.17 | 3.86E-02 |
| P12273 | 0.17 | 2.68E-02 |
| Q8NBJ4 | 0.16 | 2.36E-02 |
| P31997 | 0.15 | 4.02E-02 |
| P55017 | 0.13 | 1.29E-02 |
| P41181 | 0.09 | 3.56E-02 |
| P10721 | 0.09 | 2.68E-02 |

| F5 | | |
|---|---|---|
| **Protein Accessions** | **Fold Change** | **P value** |
| P04279 | 17.62 | 2.84E-05 |
| Q02383 | 14.28 | 4.15E-06 |
| Q93091 | 12.23 | 1.23E-06 |
| P02750 | 9.46 | 1.60E-05 |
| O00754 | 8.94 | 2.99E-07 |
| Q13621 | 8.35 | 1.66E-06 |
| Q15293 | 6.34 | 6.93E-04 |
| Q86VP6 | 5.45 | 5.83E-05 |
| Q9H6S3 | 4.86 | 3.35E-04 |
| P50225 | 4.69 | 1.85E-03 |
| O00115 | 4.47 | 1.60E-05 |
| P22748 | 4.31 | 3.08E-05 |
| Q9BTY2 | 4.24 | 5.83E-05 |
| Q07075 | 4.12 | 1.29E-02 |
| P07339 | 4.09 | 1.19E-05 |
| P07858 | 4.07 | 8.40E-05 |
| P15848 | 4.04 | 1.81E-04 |
| Q13510 | 4.04 | 2.46E-05 |
| Q12929 | 4.00 | 2.98E-04 |
| Q99653 | 3.89 | 1.60E-05 |
| Q96RF0 | 3.80 | 9.51E-05 |
| P01911 | 3.77 | 6.22E-04 |
| Q6W4X9 | 3.75 | 1.25E-02 |
| P26038 | 3.74 | 1.26E-04 |
| Q9NUM4 | 3.70 | 1.30E-03 |
| P07686 | 3.69 | 8.40E-05 |
| Q9HD89 | 3.69 | 1.41E-04 |
| P02741 | 3.69 | 3.98E-05 |
| P02763 | 3.64 | 1.60E-03 |
| Q9UBR2 | 3.61 | 1.50E-06 |
| Q8N5I2 | 3.60 | 5.83E-05 |
| P17900 | 3.59 | 1.61E-02 |
| Q16819 | 3.58 | 2.02E-02 |
| P05937 | 3.56 | 4.72E-03 |
| P01903 | 3.56 | 3.53E-02 |
| P17050 | 3.56 | 8.08E-05 |
| O94832 | 3.51 | 1.26E-04 |

| | | |
|---|---|---|
| Q13113 | 3.46 | 3.10E-04 |
| P04066 | 3.45 | 4.46E-05 |
| Q15274 | 3.35 | 6.87E-03 |
| Q9H223 | 3.32 | 7.06E-04 |
| Q9Y6X5 | 3.31 | 6.22E-04 |
| Q12765 | 3.28 | 5.87E-03 |
| Q9UHR4 | 3.27 | 1.43E-04 |
| P50570 | 3.20 | 1.89E-04 |
| P09668 | 3.19 | 3.10E-04 |
| P25774 | 3.19 | 6.22E-04 |
| Q9BYG5 | 3.16 | 1.51E-04 |
| Q7L5L3 | 3.15 | 5.17E-03 |
| O75629 | 3.15 | 6.70E-04 |
| P53634 | 3.15 | 5.83E-05 |
| Q9HBH0 | 3.12 | 2.95E-04 |
| P10619 | 3.04 | 4.00E-04 |
| Q969L2 | 3.01 | 3.35E-03 |
| P20472 | 3.00 | 1.92E-02 |
| Q9UQB8 | 2.99 | 2.81E-06 |
| P06865 | 2.95 | 8.08E-05 |
| Q8TE68 | 2.94 | 5.08E-04 |
| P09936 | 2.90 | 7.00E-03 |
| Q9UGM3 | 2.90 | 9.10E-03 |
| P15311 | 2.89 | 9.88E-04 |
| P50148 | 2.88 | 5.08E-04 |
| P17405 | 2.84 | 1.16E-04 |
| O96009 | 2.83 | 3.95E-03 |
| P54803 | 2.81 | 4.63E-04 |
| Q9Y646 | 2.80 | 1.37E-03 |
| O75348 | 2.79 | 6.93E-04 |
| P00492 | 2.79 | 3.34E-04 |
| A6NIZ1 | 2.76 | 5.20E-03 |
| P16278 | 2.76 | 1.53E-03 |
| Q8NCC3 | 2.75 | 1.74E-03 |
| P00746 | 2.75 | 1.08E-02 |
| O00338 | 2.74 | 1.14E-03 |
| P35241 | 2.71 | 9.51E-05 |
| P15313 | 2.71 | 1.43E-03 |
| Q6UWR7 | 2.70 | 2.08E-02 |

| | | |
|---|---|---|
| P29992 | 2.68 | 4.50E-04 |
| P34059 | 2.66 | 3.83E-03 |
| Q9H3G5 | 2.65 | 2.49E-03 |
| P62195 | 2.65 | 2.78E-02 |
| Q9NZN3 | 2.62 | 7.92E-04 |
| P00568 | 2.61 | 3.35E-04 |
| P50897 | 2.58 | 5.83E-05 |
| Q9HBG4 | 2.58 | 3.69E-02 |
| P38606 | 2.57 | 2.95E-04 |
| P11279 | 2.55 | 8.21E-03 |
| P05413 | 2.54 | 3.22E-02 |
| Q5JS37 | 2.54 | 3.96E-03 |
| O00299 | 2.54 | 2.34E-04 |
| Q15833 | 2.52 | 1.70E-03 |
| Q96LD4 | 2.51 | 1.43E-04 |
| O75695 | 2.51 | 1.98E-03 |
| Q9NTK5 | 2.51 | 4.75E-03 |
| Q9HB40 | 2.49 | 7.62E-03 |
| Q9Y6E0 | 2.49 | 5.16E-03 |
| Q8NHP8 | 2.49 | 4.50E-04 |
| P21266 | 2.49 | 2.87E-03 |
| Q9UM54 | 2.45 | 1.74E-02 |
| P17858 | 2.45 | 1.88E-03 |
| Q9UI12 | 2.43 | 1.08E-03 |
| Q8N357 | 2.41 | 1.16E-02 |
| P36222 | 2.41 | 1.03E-02 |
| P84022 | 2.38 | 4.08E-02 |
| P54793 | 2.36 | 3.40E-02 |
| P00918 | 2.36 | 3.53E-02 |
| P60953 | 2.36 | 1.70E-02 |
| O00159 | 2.36 | 1.29E-02 |
| P02511 | 2.35 | 1.70E-02 |
| Q8NFJ5 | 2.35 | 2.58E-02 |
| Q9HAU8 | 2.33 | 1.43E-03 |
| P28161 | 2.33 | 5.47E-03 |
| P51688 | 2.32 | 3.40E-02 |
| P38646 | 2.32 | 4.93E-03 |
| P36871 | 2.30 | 1.94E-02 |
| Q16851 | 2.30 | 3.40E-03 |

| | | |
|---|---|---|
| P98164 | 2.30 | 3.29E-04 |
| Q99519 | 2.30 | 9.51E-05 |
| P62834 | 2.26 | 1.06E-02 |
| Q8WWB7 | 2.26 | 3.44E-03 |
| P62330 | 2.26 | 1.23E-02 |
| P24593 | 2.25 | 1.08E-02 |
| Q9Y376 | 2.24 | 8.69E-03 |
| P15941 | 2.16 | 1.03E-02 |
| P35080 | 2.16 | 1.15E-02 |
| P08183 | 2.15 | 1.01E-02 |
| Q12923 | 2.15 | 2.76E-02 |
| P16152 | 2.13 | 7.62E-03 |
| Q13277 | 2.12 | 4.50E-04 |
| P41181 | 2.11 | 2.78E-02 |
| Q14764 | 2.11 | 1.02E-02 |
| Q9BQI0 | 2.10 | 3.66E-02 |
| P08754 | 2.10 | 1.18E-02 |
| P36543 | 2.08 | 1.43E-03 |
| P07602 | 2.08 | 7.62E-03 |
| Q9UHI7 | 2.08 | 1.45E-02 |
| P17174 | 2.07 | 8.94E-03 |
| P42685 | 2.07 | 2.01E-02 |
| P18669 | 2.07 | 1.84E-02 |
| Q8IV08 | 2.06 | 1.74E-02 |
| P15586 | 2.06 | 2.48E-03 |
| P30044 | 2.06 | 1.07E-02 |
| P40925 | 2.04 | 1.43E-03 |
| Q14344 | 2.02 | 8.02E-03 |
| Q9HBR0 | 0.49 | 3.22E-02 |
| P29323 | 0.49 | 1.40E-02 |
| P04180 | 0.47 | 4.06E-02 |
| P10586 | 0.47 | 6.03E-03 |
| P30530 | 0.46 | 3.83E-03 |
| P43121 | 0.45 | 1.03E-02 |
| Q13445 | 0.45 | 5.37E-03 |
| P09619 | 0.44 | 1.65E-02 |
| Q9BQ51 | 0.43 | 3.44E-03 |
| Q9UNN8 | 0.42 | 4.73E-03 |
| P13598 | 0.41 | 3.28E-02 |

| | | |
|---|---|---|
| P21709 | 0.40 | 3.75E-02 |
| P28827 | 0.40 | 2.38E-02 |
| Q15223 | 0.39 | 1.18E-02 |
| Q06418 | 0.38 | 3.35E-02 |
| P19256 | 0.37 | 2.63E-02 |
| Q96JQ0 | 0.36 | 1.94E-02 |
| Q9NQS3 | 0.36 | 8.02E-03 |
| Q06481 | 0.36 | 1.03E-02 |
| P52798 | 0.35 | 3.19E-02 |
| P45877 | 0.35 | 4.06E-02 |
| Q13443 | 0.35 | 4.33E-02 |
| Q8TB96 | 0.35 | 1.18E-02 |
| P15151 | 0.34 | 3.35E-02 |
| Q99795 | 0.34 | 4.23E-02 |
| Q8N386 | 0.34 | 7.00E-03 |
| Q92859 | 0.32 | 7.38E-03 |
| Q02413 | 0.32 | 4.16E-02 |
| O75144 | 0.31 | 1.65E-02 |
| P48740 | 0.31 | 2.83E-02 |
| Q86T13 | 0.31 | 7.38E-03 |
| P0C7U0 | 0.30 | 3.78E-02 |
| Q9UBG0 | 0.30 | 5.08E-03 |
| Q9H1U4 | 0.30 | 3.63E-02 |
| Q6UXG3 | 0.29 | 2.58E-02 |
| P11047 | 0.27 | 7.38E-03 |
| Q9HCU0 | 0.26 | 1.43E-02 |
| O75339 | 0.24 | 2.58E-02 |
| O43921 | 0.23 | 1.92E-02 |
| Q9BWV1 | 0.21 | 1.37E-02 |
| Q8NDA2 | 0.16 | 3.12E-02 |

| F6 | | |
|---|---|---|
| **Protein Accessions** | **Fold Change** | **P value** |
| P05413 | 78.04 | 9.59E-07 |
| O15540 | 55.00 | 9.15E-07 |
| Q15293 | 49.47 | 2.28E-10 |
| P01911 | 31.69 | 9.43E-10 |
| P61769 | 27.63 | 1.37E-07 |
| P02750 | 26.43 | 4.68E-08 |
| P01903 | 19.95 | 1.32E-06 |
| Q9NWV4 | 18.19 | 3.68E-08 |
| Q9UBR2 | 16.90 | 1.96E-08 |
| P04179 | 16.32 | 1.55E-07 |
| P02763 | 15.75 | 3.41E-08 |
| P05534 | 15.53 | 6.70E-07 |
| Q8N357 | 13.89 | 1.96E-08 |
| P09668 | 13.45 | 1.25E-08 |
| P01700 | 12.73 | 4.25E-07 |
| Q9P1F3 | 12.31 | 1.78E-06 |
| P11684 | 11.77 | 9.23E-05 |
| O75368 | 11.74 | 2.67E-07 |
| Q92484 | 11.72 | 2.57E-06 |
| Q969H8 | 11.60 | 1.85E-06 |
| P01717 | 11.17 | 4.16E-08 |
| P01714 | 11.04 | 9.76E-05 |
| P02792 | 11.02 | 1.05E-06 |
| P0DOX3 | 10.80 | 1.59E-05 |
| A0A075B6K5 | 10.64 | 2.47E-06 |
| P00915 | 10.35 | 1.99E-05 |
| P02655 | 10.00 | 4.11E-05 |
| P00751 | 9.86 | 5.34E-04 |
| Q9UHI8 | 9.80 | 9.23E-05 |
| Q99988 | 9.68 | 2.01E-05 |
| P02776 | 9.59 | 2.11E-07 |
| Q93091 | 9.44 | 7.82E-07 |
| P30508 | 9.32 | 8.86E-07 |
| Q9Y6X5 | 9.01 | 2.11E-07 |
| Q07075 | 8.84 | 3.39E-05 |
| P02511 | 8.78 | 4.25E-07 |
| Q96H15 | 8.74 | 3.18E-07 |
| P00441 | 8.68 | 2.37E-07 |
| O43491 | 8.57 | 1.25E-06 |
| P0DOX8 | 8.55 | 2.11E-07 |
| P00742 | 8.49 | 1.23E-06 |
| Q9BZM5 | 8.23 | 1.38E-06 |
| P02753 | 8.10 | 5.66E-07 |
| O43556 | 8.04 | 1.32E-06 |
| O00244 | 7.99 | 4.36E-05 |
| P01011 | 7.99 | 4.85E-07 |

| | | |
|---|---|---|
| P17405 | 7.92 | 3.68E-08 |
| P17900 | 7.72 | 3.55E-05 |
| P15814 | 7.71 | 3.81E-07 |
| Q92954 | 7.49 | 9.15E-05 |
| Q15274 | 7.47 | 7.07E-06 |
| P36980 | 7.41 | 7.11E-06 |
| P01721 | 7.36 | 8.18E-05 |
| P26641 | 7.36 | 6.60E-06 |
| P02656 | 7.33 | 1.00E-05 |
| A0A075B6I0 | 7.28 | 4.95E-06 |
| P27824 | 7.27 | 7.07E-06 |
| P27797 | 7.27 | 7.51E-05 |
| P06454 | 7.18 | 3.12E-04 |
| P52272 | 7.07 | 2.06E-03 |
| P78310 | 6.92 | 4.05E-06 |
| P19652 | 6.60 | 6.90E-06 |
| Q92530 | 6.56 | 5.20E-06 |
| P20774 | 6.50 | 3.17E-06 |
| Q8IX05 | 6.49 | 5.66E-07 |
| P18465 | 6.48 | 4.11E-05 |
| P07306 | 6.48 | 3.79E-05 |
| P13640 | 6.40 | 2.37E-04 |
| Q6UWR7 | 6.24 | 4.11E-05 |
| Q9UHY7 | 6.13 | 1.36E-06 |
| P04211 | 6.08 | 1.89E-06 |
| P25311 | 5.96 | 1.58E-04 |
| P01703 | 5.93 | 1.22E-05 |
| Q92887 | 5.88 | 2.41E-05 |
| P50225 | 5.87 | 1.22E-03 |
| P25774 | 5.87 | 1.58E-06 |
| P01009 | 5.84 | 4.25E-07 |
| P00995 | 5.81 | 3.48E-04 |
| P0DOY3 | 5.76 | 3.38E-04 |
| P84090 | 5.75 | 7.12E-04 |
| P01699 | 5.73 | 2.47E-06 |
| A0A0C4DH72 | 5.72 | 6.12E-04 |
| O00548 | 5.58 | 1.37E-02 |
| Q13740 | 5.52 | 4.25E-07 |
| P21583 | 5.49 | 1.59E-05 |
| Q8NFU3 | 5.47 | 1.79E-04 |
| Q9H8J5 | 5.45 | 1.17E-05 |
| Q6UWV6 | 5.43 | 3.58E-06 |
| Q9BW04 | 5.43 | 2.21E-04 |
| P0CF74 | 5.34 | 6.53E-03 |
| Q8NCW5 | 5.20 | 2.54E-04 |
| Q9BTY2 | 5.19 | 2.47E-06 |
| O00754 | 5.18 | 1.32E-06 |
| P07359 | 5.13 | 7.59E-06 |

| | | |
|---|---|---|
| P29966 | 5.10 | 1.04E-04 |
| Q13228 | 5.07 | 1.62E-06 |
| Q13113 | 5.01 | 5.66E-06 |
| P07686 | 5.01 | 2.44E-06 |
| P27487 | 4.98 | 4.32E-06 |
| P15144 | 4.90 | 3.87E-04 |
| P02741 | 4.83 | 5.66E-07 |
| O60234 | 4.77 | 5.12E-05 |
| Q96IU4 | 4.76 | 8.36E-05 |
| O95861 | 4.72 | 2.06E-03 |
| Q9Y490 | 4.71 | 9.15E-05 |
| A0A075B6S5 | 4.69 | 9.52E-05 |
| Q9H299 | 4.66 | 5.04E-03 |
| Q8NCC3 | 4.64 | 1.10E-05 |
| Q9Y547 | 4.63 | 1.28E-05 |
| A0A075B6H9 | 4.62 | 9.83E-04 |
| P02794 | 4.56 | 4.48E-03 |
| P01599 | 4.56 | 7.83E-03 |
| P00740 | 4.55 | 9.83E-04 |
| P10109 | 4.54 | 7.36E-05 |
| Q9Y4L1 | 4.48 | 5.52E-04 |
| O00584 | 4.45 | 3.13E-05 |
| P12111 | 4.37 | 2.40E-04 |
| P35754 | 4.32 | 8.16E-04 |
| P53634 | 4.31 | 3.58E-06 |
| P0C0L4 | 4.30 | 3.78E-04 |
| P14207 | 4.26 | 9.30E-04 |
| P04066 | 4.26 | 2.06E-06 |
| P35080 | 4.25 | 1.48E-04 |
| Q02985 | 4.22 | 5.20E-06 |
| P00167 | 4.21 | 2.20E-03 |
| A0A0C4DH68 | 4.18 | 3.89E-05 |
| Q9Y5Y7 | 4.15 | 1.46E-04 |
| Q14019 | 4.15 | 1.21E-04 |
| Q9NUM4 | 4.12 | 2.37E-04 |
| A2NJV5 | 4.08 | 2.49E-04 |
| Q9HB90 | 4.05 | 3.20E-03 |
| P46108 | 4.05 | 2.01E-03 |
| P07737 | 4.04 | 1.85E-05 |
| Q9HC35 | 4.03 | 5.17E-05 |
| P01019 | 4.01 | 3.55E-05 |
| P01624 | 3.99 | 8.55E-04 |
| P12104 | 3.98 | 1.77E-03 |
| Q6ZVN8 | 3.98 | 1.86E-03 |
| P01601 | 3.92 | 4.23E-03 |
| Q92542 | 3.90 | 8.08E-04 |
| P06865 | 3.89 | 3.35E-06 |
| P19320 | 3.88 | 3.06E-03 |

| | | |
|---|---|---|
| P04217 | 3.86 | 1.78E-03 |
| P01619 | 3.85 | 3.20E-03 |
| P08185 | 3.83 | 4.71E-06 |
| O43399 | 3.80 | 1.38E-03 |
| P00450 | 3.80 | 1.90E-04 |
| Q96FW1 | 3.72 | 5.15E-03 |
| P09923 | 3.72 | 2.01E-02 |
| P23083 | 3.71 | 2.61E-04 |
| P61916 | 3.68 | 4.65E-07 |
| Q6FHJ7 | 3.67 | 5.57E-03 |
| P01860 | 3.61 | 1.06E-02 |
| Q03591 | 3.55 | 1.01E-02 |
| O43570 | 3.55 | 4.13E-05 |
| P80723 | 3.53 | 3.43E-02 |
| Q8WU39 | 3.52 | 3.84E-04 |
| Q8IW52 | 3.47 | 2.32E-03 |
| P83110 | 3.46 | 6.52E-04 |
| P54289 | 3.40 | 1.10E-03 |
| Q9Y274 | 3.40 | 2.84E-03 |
| P01715 | 3.38 | 3.55E-05 |
| O75608 | 3.35 | 1.39E-05 |
| Q99972 | 3.33 | 7.39E-04 |
| P01344 | 3.28 | 1.08E-04 |
| P28838 | 3.28 | 3.87E-04 |
| Q15121 | 3.28 | 1.82E-03 |
| Q9Y5Z4 | 3.27 | 9.34E-04 |
| P35613 | 3.20 | 8.11E-04 |
| Q03167 | 3.17 | 8.55E-04 |
| Q5JTV8 | 3.17 | 2.57E-03 |
| P30048 | 3.16 | 2.08E-02 |
| P13667 | 3.16 | 9.11E-05 |
| O15204 | 3.15 | 5.45E-03 |
| Q9Y275 | 3.13 | 8.96E-03 |
| P60983 | 3.12 | 6.98E-03 |
| Q8NBS9 | 3.12 | 1.87E-04 |
| P22223 | 3.11 | 8.11E-04 |
| P67809 | 3.10 | 2.09E-03 |
| P37840 | 3.08 | 9.41E-03 |
| P62328 | 3.08 | 1.04E-02 |
| A0A075B6K4 | 3.06 | 7.83E-03 |
| P50440 | 3.05 | 3.32E-02 |
| O76076 | 3.05 | 1.54E-02 |
| Q9BYE9 | 3.04 | 3.50E-04 |
| Q86YD5 | 3.04 | 1.01E-02 |
| P50053 | 3.03 | 5.91E-04 |
| Q12765 | 3.03 | 4.88E-03 |
| P55103 | 3.03 | 4.88E-03 |
| ENSEMBL:ENSBTAP00000007350 | 3.00 | 3.09E-03 |

| | | |
|---|---|---|
| P20142 | 3.00 | 4.96E-02 |
| Q9NY33 | 3.00 | 2.19E-02 |
| O75531 | 2.98 | 1.98E-03 |
| Q8NFQ8 | 2.93 | 7.58E-03 |
| Q9H0U4 | 2.93 | 3.73E-02 |
| Q9H6X2 | 2.91 | 5.57E-03 |
| P02671 | 2.89 | 2.87E-03 |
| P0DOX7 | 2.89 | 2.32E-03 |
| P10619 | 2.89 | 2.17E-04 |
| Q13421 | 2.88 | 2.48E-02 |
| P18428 | 2.85 | 3.19E-04 |
| P02790 | 2.85 | 9.96E-05 |
| P30101 | 2.84 | 5.45E-03 |
| P08571 | 2.84 | 3.94E-03 |
| Q8N2U0 | 2.81 | 1.06E-02 |
| P04233 | 2.77 | 1.09E-02 |
| Q8N126 | 2.75 | 3.34E-03 |
| P15586 | 2.72 | 4.11E-05 |
| Q00839 | 2.72 | 3.70E-03 |
| Q9NVJ2 | 2.72 | 1.33E-02 |
| P07858 | 2.71 | 7.91E-04 |
| Q9UGM5 | 2.70 | 5.45E-03 |
| P11279 | 2.69 | 2.40E-03 |
| Q92520 | 2.68 | 8.08E-05 |
| Q13510 | 2.66 | 1.33E-04 |
| A0A087WSX0 | 2.66 | 2.68E-02 |
| A0A0B4J1V6 | 2.65 | 1.91E-03 |
| P19438 | 2.65 | 3.81E-04 |
| P06312 | 2.63 | 8.47E-03 |
| A0A0A0MS15 | 2.63 | 3.20E-03 |
| P04216 | 2.62 | 1.87E-05 |
| Q24JP5 | 2.61 | 4.66E-02 |
| P12318 | 2.60 | 2.87E-02 |
| P06681 | 2.56 | 3.45E-02 |
| A0A075B6Q5 | 2.56 | 2.94E-02 |
| Q8NHJ6 | 2.54 | 3.78E-03 |
| Q9UHI7 | 2.53 | 9.42E-03 |
| P07225 | 2.53 | 1.20E-03 |
| Q03405 | 2.51 | 8.85E-03 |
| P16422 | 2.50 | 1.17E-02 |
| P01593 | 2.49 | 1.07E-02 |
| Q96FN5 | 2.49 | 3.82E-02 |
| Q7L5L3 | 2.49 | 1.37E-02 |
| P0CG47 | 2.48 | 5.54E-03 |
| Q9HD89 | 2.47 | 6.55E-03 |
| P39687 | 2.46 | 3.56E-02 |
| Q92820 | 2.46 | 7.39E-04 |
| P05362 | 2.46 | 2.90E-02 |

| | | |
|---|---|---|
| P07339 | 2.45 | 2.37E-04 |
| P00746 | 2.45 | 1.04E-02 |
| SWISS-PROT:P00978 | 2.44 | 1.43E-02 |
| Q02487 | 2.43 | 6.41E-03 |
| Q6UXB8 | 2.42 | 2.32E-02 |
| P07602 | 2.42 | 8.55E-04 |
| Q96GW7 | 2.40 | 2.98E-02 |
| Q16563 | 2.39 | 1.58E-02 |
| Q92626 | 2.39 | 1.88E-03 |
| P02452 | 2.39 | 3.88E-02 |
| Q8WWB7 | 2.39 | 1.09E-02 |
| Q92692 | 2.38 | 4.38E-03 |
| Q9BVM4 | 2.38 | 6.85E-03 |
| Q14112 | 2.38 | 6.99E-03 |
| P17858 | 2.37 | 1.94E-03 |
| O95336 | 2.35 | 3.69E-03 |
| Q14108 | 2.35 | 1.13E-02 |
| P48551 | 2.34 | 3.59E-02 |
| O75936 | 2.34 | 1.66E-03 |
| Q6ZQN7 | 2.33 | 4.29E-02 |
| P29279 | 2.31 | 4.74E-03 |
| P05091 | 2.30 | 2.19E-02 |
| P02760 | 2.30 | 1.72E-02 |
| A0A0J9YXX1 | 2.29 | 1.01E-02 |
| P0C0L5 | 2.29 | 4.89E-03 |
| O75503 | 2.26 | 6.22E-04 |
| P61026 | 2.25 | 3.52E-02 |
| P02787 | 2.24 | 2.83E-02 |
| Q9HAT2 | 2.24 | 2.24E-03 |
| Q9BPX5 | 2.24 | 1.90E-02 |
| Q9Y646 | 2.23 | 4.34E-03 |
| P55285 | 2.23 | 1.04E-02 |
| P36941 | 2.22 | 6.85E-03 |
| P00747 | 2.22 | 1.71E-02 |
| P23470 | 2.21 | 5.56E-04 |
| Q9BTM9 | 2.20 | 1.01E-02 |
| P07711 | 2.18 | 1.07E-03 |
| P17936 | 2.16 | 2.13E-02 |
| Q96HD9 | 2.15 | 3.24E-02 |
| Q92496 | 2.15 | 3.88E-02 |
| Q9H6B4 | 2.14 | 1.67E-02 |
| Q96CX2 | 2.13 | 1.33E-02 |
| P11597 | 2.13 | 1.06E-02 |
| P05981 | 2.13 | 3.78E-04 |
| P01008 | 2.12 | 2.40E-03 |
| Q99571 | 2.11 | 1.04E-02 |
| Q8TDQ1 | 2.10 | 1.01E-02 |
| Q14624 | 2.10 | 2.89E-02 |

| | | |
|---|---|---|
| P09382 | 2.10 | 2.78E-02 |
| P98172 | 2.10 | 8.64E-03 |
| Q92485 | 2.09 | 3.24E-02 |
| Q14165 | 2.07 | 1.15E-02 |
| P01859 | 2.07 | 3.62E-02 |
| Q15043 | 2.06 | 1.10E-02 |
| Q16851 | 2.06 | 4.38E-03 |
| P28300 | 2.04 | 3.29E-02 |
| Q9UM22 | 2.02 | 6.86E-03 |
| P01871 | 2.02 | 2.69E-02 |
| Q92734 | 2.01 | 3.62E-02 |
| P09327 | 2.00 | 3.24E-02 |
| Q8NC42 | 0.49 | 4.96E-02 |
| P55899 | 0.48 | 3.04E-02 |
| O60361 | 0.48 | 4.36E-02 |
| O43278 | 0.47 | 2.86E-02 |
| Q9HBR0 | 0.47 | 1.46E-02 |
| Q01973 | 0.46 | 3.81E-02 |
| O60888 | 0.46 | 8.47E-03 |
| Q86T13 | 0.45 | 1.15E-02 |
| P16234 | 0.45 | 4.65E-02 |
| Q9Y287 | 0.45 | 3.26E-02 |
| P10586 | 0.45 | 2.32E-03 |
| Q8N386 | 0.43 | 7.83E-03 |
| Q9UBG0 | 0.42 | 7.19E-03 |
| P43121 | 0.41 | 4.00E-03 |
| P05026 | 0.41 | 2.32E-02 |
| Q9UNN8 | 0.40 | 2.06E-03 |
| Q9NPF0 | 0.40 | 3.62E-02 |
| Q9Y4D7 | 0.40 | 2.32E-02 |
| P29323 | 0.40 | 2.09E-03 |
| P02774 | 0.40 | 3.64E-02 |
| Q9BQ51 | 0.40 | 1.23E-03 |
| O00391 | 0.39 | 1.10E-02 |
| P63241 | 0.39 | 4.69E-02 |
| P40189 | 0.39 | 2.26E-02 |
| Q9Y653 | 0.39 | 4.92E-02 |
| O60449 | 0.39 | 4.63E-02 |
| P30530 | 0.38 | 1.65E-03 |
| Q08722 | 0.38 | 1.04E-02 |
| P58499 | 0.38 | 3.50E-02 |
| P13598 | 0.38 | 1.37E-02 |
| P06280 | 0.38 | 3.26E-02 |
| P15151 | 0.38 | 2.32E-02 |
| Q6P9A2 | 0.37 | 3.82E-02 |
| Q9NQS3 | 0.37 | 4.61E-03 |
| Q9Y5K6 | 0.37 | 3.63E-02 |
| Q6P531 | 0.36 | 2.32E-02 |

| | | |
|---|---|---|
| Q9P0T7 | 0.36 | 4.98E-02 |
| P30086 | 0.36 | 1.37E-02 |
| Q04760 | 0.35 | 1.80E-02 |
| Q08334 | 0.34 | 2.65E-02 |
| P63092 | 0.33 | 2.70E-02 |
| P82980 | 0.33 | 3.36E-02 |
| Q9HBB8 | 0.33 | 4.62E-02 |
| Q06418 | 0.33 | 1.23E-02 |
| Q06481 | 0.33 | 4.59E-03 |
| P12259 | 0.33 | 1.75E-02 |
| P15291 | 0.32 | 1.33E-02 |
| Q6UX71 | 0.32 | 2.56E-03 |
| Q15223 | 0.32 | 3.70E-03 |
| P17931 | 0.32 | 4.25E-02 |
| Q6EMK4 | 0.31 | 3.20E-02 |
| Q9NY97 | 0.31 | 4.26E-02 |
| Q12860 | 0.31 | 4.61E-02 |
| P16070 | 0.31 | 1.34E-02 |
| Q7Z7M0 | 0.31 | 2.26E-02 |
| Q86YT9 | 0.30 | 1.71E-02 |
| Q8WUM4 | 0.30 | 1.06E-02 |
| Q9HCU0 | 0.30 | 1.03E-02 |
| P0DMQ5 | 0.30 | 1.71E-02 |
| Q02413 | 0.30 | 2.13E-02 |
| O43895 | 0.29 | 3.31E-02 |
| Q99816 | 0.29 | 2.63E-02 |
| P15313 | 0.29 | 4.55E-02 |
| Q8N5I2 | 0.29 | 2.32E-02 |
| Q8NBJ4 | 0.29 | 4.05E-02 |
| P14784 | 0.29 | 2.12E-02 |
| Q9Y639 | 0.29 | 3.07E-02 |
| Q9H9H4 | 0.28 | 3.43E-02 |
| Q6UXG3 | 0.28 | 1.34E-02 |
| Q8TB96 | 0.28 | 4.15E-03 |
| Q9H4M9 | 0.28 | 3.41E-03 |
| Q9UQB8 | 0.28 | 2.40E-04 |
| Q7KYR7 | 0.28 | 4.74E-03 |
| Q9HCM3 | 0.28 | 8.47E-03 |
| P32942 | 0.27 | 3.15E-02 |
| P09486 | 0.27 | 1.34E-02 |
| Q16651 | 0.27 | 3.24E-02 |
| P21266 | 0.27 | 3.88E-02 |
| P09619 | 0.27 | 2.55E-03 |
| Q9BY67 | 0.27 | 3.99E-02 |
| P61106 | 0.27 | 2.86E-02 |
| O75351 | 0.27 | 2.13E-02 |
| Q13443 | 0.26 | 1.44E-02 |
| P20827 | 0.26 | 5.91E-03 |

| | | |
|---|---|---|
| P05452 | 0.26 | 3.81E-02 |
| Q05707 | 0.26 | 1.04E-02 |
| Q99835 | 0.26 | 2.06E-03 |
| Q96PD5 | 0.25 | 3.15E-02 |
| Q9H741 | 0.25 | 1.24E-02 |
| Q8IWA5 | 0.25 | 1.86E-02 |
| Q92673 | 0.24 | 1.01E-02 |
| O75144 | 0.24 | 4.52E-03 |
| Q01151 | 0.24 | 1.62E-02 |
| Q96FE7 | 0.24 | 2.66E-02 |
| Q8TBP5 | 0.24 | 1.52E-02 |
| Q9UJJ9 | 0.24 | 9.42E-03 |
| Q9Y376 | 0.24 | 3.29E-02 |
| Q04721 | 0.23 | 2.11E-02 |
| Q9H9P2 | 0.23 | 4.69E-02 |
| P19256 | 0.23 | 5.71E-03 |
| Q12913 | 0.23 | 1.76E-02 |
| Q96CF2 | 0.23 | 3.82E-02 |
| O75882 | 0.22 | 4.15E-03 |
| Q9NP85 | 0.22 | 1.39E-02 |
| Q15116 | 0.22 | 7.52E-03 |
| P01042 | 0.22 | 3.45E-02 |
| P04180 | 0.22 | 3.76E-03 |
| Q5SZK8 | 0.21 | 4.59E-03 |
| Q96NY8 | 0.21 | 1.78E-02 |
| Q13591 | 0.20 | 2.91E-02 |
| P35858 | 0.20 | 4.26E-02 |
| P16035 | 0.20 | 1.16E-02 |
| P26992 | 0.19 | 1.30E-02 |
| Q6UY11 | 0.19 | 3.74E-02 |
| Q9UK41 | 0.19 | 8.00E-03 |
| Q92859 | 0.19 | 1.46E-03 |
| P15941 | 0.19 | 2.69E-02 |
| P24855 | 0.19 | 5.45E-03 |
| Q15262 | 0.18 | 5.15E-03 |
| P11047 | 0.18 | 2.09E-03 |
| P23284 | 0.18 | 9.29E-03 |
| Q9UGT4 | 0.18 | 2.51E-02 |
| Q96JQ0 | 0.18 | 3.09E-03 |
| P27930 | 0.17 | 3.31E-02 |
| P45877 | 0.17 | 8.21E-03 |
| Q9NZZ3 | 0.17 | 4.68E-02 |
| P50148 | 0.17 | 1.62E-02 |
| Q86XT2 | 0.16 | 3.50E-02 |
| Q9UKU6 | 0.16 | 5.16E-03 |
| Q9ULK6 | 0.16 | 1.11E-02 |
| Q8IUL8 | 0.16 | 8.65E-03 |
| Q9UN37 | 0.16 | 6.55E-03 |

| | | |
|---|---|---|
| Q9NU53 | 0.15 | 2.55E-03 |
| Q5IJ48 | 0.15 | 3.26E-02 |
| Q9H0X4 | 0.15 | 5.39E-03 |
| P08582 | 0.14 | 3.45E-02 |
| O75339 | 0.14 | 7.83E-03 |
| Q9UQN3 | 0.14 | 4.87E-02 |
| Q96EY5 | 0.14 | 5.60E-03 |
| P80370 | 0.13 | 2.26E-02 |
| P41217 | 0.13 | 6.41E-03 |
| Q9NP79 | 0.13 | 1.42E-02 |
| Q9H1U4 | 0.13 | 7.83E-03 |
| P41181 | 0.13 | 3.92E-02 |
| P32004 | 0.12 | 4.36E-02 |
| Q9UBQ6 | 0.11 | 1.21E-02 |
| Q7LBR1 | 0.11 | 8.26E-03 |
| Q5VW32 | 0.11 | 3.82E-02 |
| P21926 | 0.11 | 3.36E-02 |
| Q12864 | 0.10 | 4.59E-03 |
| P0C7U0 | 0.10 | 6.86E-03 |
| Q9P121 | 0.10 | 3.88E-02 |
| Q8NDA2 | 0.09 | 1.18E-02 |
| P55017 | 0.09 | 1.01E-02 |
| Q8IX04 | 0.08 | 4.65E-02 |
| Q9Y6W3 | 0.07 | 2.69E-02 |
| P12273 | 0.07 | 1.61E-02 |
| Q6V0I7 | 0.07 | 1.23E-02 |

| F7 | | |
|---|---|---|
| **Protein Accessions** | **Fold Change** | **P value** |
| Q8WWA0 | 17.17 | 4.18E-07 |
| Q07837 | 13.00 | 9.22E-08 |
| Q8N2U0 | 12.29 | 4.18E-07 |
| Q99102 | 11.38 | 1.41E-05 |
| Q00325 | 10.97 | 2.44E-06 |
| P21796 | 10.77 | 4.17E-07 |
| P45880 | 10.20 | 4.60E-06 |
| Q9UGM3 | 9.47 | 5.11E-07 |
| Q7L5L3 | 9.27 | 1.95E-06 |
| Q8N357 | 8.60 | 8.02E-07 |
| P02792 | 7.93 | 3.42E-05 |
| Q9H3K2 | 6.89 | 2.76E-05 |
| P49788 | 6.63 | 2.92E-04 |
| Q13113 | 6.62 | 4.60E-06 |
| P02741 | 6.31 | 4.17E-07 |
| P27105 | 5.90 | 1.60E-05 |
| P01903 | 5.87 | 1.41E-03 |
| Q92542 | 5.66 | 1.05E-04 |
| Q14116 | 5.52 | 4.29E-04 |
| Q6UX06 | 5.45 | 1.17E-03 |
| Q695T7 | 5.33 | 1.05E-04 |
| P11279 | 5.20 | 3.15E-05 |
| P09923 | 5.18 | 4.50E-03 |
| Q9HD89 | 5.04 | 2.13E-05 |
| O96009 | 4.95 | 4.38E-05 |
| P10109 | 4.85 | 1.72E-04 |
| Q8N307 | 4.76 | 1.05E-04 |
| Q1EHB4 | 4.74 | 1.05E-04 |
| Q9NVJ2 | 4.71 | 3.85E-04 |
| Q9UHG3 | 4.58 | 1.69E-04 |
| Q9Y6R7 | 4.52 | 2.89E-02 |
| O43866 | 4.49 | 1.41E-05 |
| P54793 | 4.40 | 3.02E-04 |
| Q16563 | 4.35 | 2.27E-04 |
| O00182 | 4.35 | 1.60E-05 |
| Q9UHI7 | 4.27 | 3.15E-05 |
| P02511 | 4.24 | 3.90E-04 |

| | | |
|---|---|---|
| Q06495 | 4.07 | 5.10E-03 |
| Q96A25 | 4.06 | 3.92E-04 |
| Q9NS93 | 4.03 | 1.88E-03 |
| A0A075B6K5 | 4.00 | 6.06E-03 |
| Q9NZT2 | 3.92 | 3.75E-03 |
| Q6UWR7 | 3.91 | 1.45E-03 |
| P36639 | 3.90 | 1.79E-04 |
| Q9NY25 | 3.89 | 3.42E-05 |
| Q9C0H2 | 3.88 | 9.09E-04 |
| Q93091 | 3.87 | 2.42E-03 |
| Q9NRX4 | 3.85 | 3.65E-02 |
| Q9BXN2 | 3.72 | 2.84E-05 |
| Q9Y696 | 3.72 | 2.63E-03 |
| P08473 | 3.68 | 1.47E-03 |
| Q9BRT3 | 3.64 | 1.47E-03 |
| A0A075B6K4 | 3.63 | 4.69E-03 |
| Q9Y512 | 3.59 | 5.09E-05 |
| Q07075 | 3.48 | 3.99E-02 |
| A0PJK1 | 3.46 | 5.87E-04 |
| Q9HB90 | 3.44 | 1.69E-02 |
| P02654 | 3.42 | 9.09E-04 |
| P61970 | 3.39 | 2.51E-04 |
| A0A0J9YXX1 | 3.34 | 5.46E-04 |
| Q14108 | 3.32 | 1.27E-03 |
| P23297 | 3.30 | 1.15E-02 |
| Q7Z3F1 | 3.28 | 5.45E-03 |
| P18428 | 3.28 | 1.88E-04 |
| Q13183 | 3.25 | 1.51E-04 |
| O43451 | 3.23 | 2.25E-03 |
| Q9H665 | 3.17 | 3.02E-04 |
| Q495M3 | 3.16 | 1.04E-03 |
| A6NI73 | 3.11 | 5.09E-04 |
| O95336 | 3.09 | 5.46E-04 |
| Q99571 | 3.00 | 8.96E-04 |
| P07320 | 2.95 | 7.62E-03 |
| Q8WWB7 | 2.93 | 3.26E-04 |
| Q9BVM4 | 2.90 | 2.00E-03 |
| P28908 | 2.89 | 6.96E-05 |
| Q9BUH6 | 2.89 | 3.23E-03 |

| | | |
|---|---|---|
| Q08380 | 2.85 | 2.91E-04 |
| P01877 | 2.84 | 1.88E-03 |
| P01876 | 2.83 | 9.18E-03 |
| Q5T2W1 | 2.82 | 4.17E-04 |
| P09668 | 2.79 | 7.31E-04 |
| P15144 | 2.75 | 4.67E-02 |
| P23083 | 2.71 | 3.53E-03 |
| P55899 | 2.70 | 1.15E-04 |
| P13688 | 2.69 | 3.82E-04 |
| P13611 | 2.69 | 4.82E-04 |
| O94832 | 2.69 | 1.86E-03 |
| P05981 | 2.65 | 9.62E-05 |
| Q14764 | 2.64 | 1.26E-03 |
| Q96LD4 | 2.62 | 4.38E-05 |
| P11117 | 2.62 | 4.02E-03 |
| P98164 | 2.59 | 7.70E-05 |
| O60271 | 2.57 | 1.09E-02 |
| Q9H756 | 2.56 | 1.05E-04 |
| P25774 | 2.54 | 3.58E-03 |
| P16278 | 2.54 | 2.90E-03 |
| P60953 | 2.53 | 8.49E-03 |
| Q6ZQN7 | 2.52 | 2.03E-03 |
| O14745 | 2.52 | 8.96E-04 |
| Q9Y2E5 | 2.49 | 4.17E-02 |
| P01589 | 2.48 | 6.05E-03 |
| Q8IV08 | 2.48 | 2.97E-03 |
| O00754 | 2.47 | 1.07E-03 |
| P01699 | 2.46 | 2.20E-03 |
| P01591 | 2.45 | 9.10E-03 |
| Q9BZZ2 | 2.44 | 1.41E-03 |
| Q496F6 | 2.44 | 4.74E-03 |
| P20062 | 2.43 | 2.34E-03 |
| P01717 | 2.42 | 1.51E-03 |
| P30508 | 2.42 | 3.16E-02 |
| Q96FL8 | 2.40 | 4.22E-02 |
| P61026 | 2.40 | 6.75E-05 |
| P10619 | 2.38 | 2.45E-03 |
| P56537 | 2.38 | 2.92E-04 |
| Q13510 | 2.38 | 8.44E-04 |

| | | |
|---|---|---|
| P01700 | 2.37 | 3.35E-04 |
| P29972 | 2.36 | 2.08E-02 |
| Q9Y6R1 | 2.35 | 4.73E-02 |
| Q9BXJ7 | 2.35 | 9.69E-03 |
| P05091 | 2.35 | 3.83E-02 |
| O15118 | 2.33 | 7.31E-04 |
| Q15116 | 2.33 | 8.44E-04 |
| Q8NHJ6 | 2.33 | 7.55E-03 |
| P16444 | 2.30 | 2.45E-02 |
| Q6UWV6 | 2.28 | 7.56E-03 |
| P01833 | 2.28 | 1.04E-03 |
| Q9NUM4 | 2.28 | 4.95E-02 |
| P02655 | 2.26 | 4.10E-02 |
| Q16581 | 2.25 | 8.17E-04 |
| Q93050 | 2.25 | 3.23E-03 |
| P51149 | 2.21 | 7.62E-03 |
| Q8NHP8 | 2.20 | 7.05E-04 |
| Q8WWV6 | 2.18 | 3.02E-02 |
| P34059 | 2.17 | 2.08E-02 |
| P01701 | 2.16 | 3.77E-02 |
| Q9Y6X5 | 2.16 | 2.17E-02 |
| Q92626 | 2.14 | 4.70E-03 |
| Q9H4M9 | 2.13 | 4.93E-04 |
| O75531 | 2.12 | 3.12E-02 |
| Q5JS37 | 2.12 | 2.03E-02 |
| Q9H223 | 2.09 | 3.43E-02 |
| O75955 | 2.09 | 9.66E-03 |
| SWISS-PROT:Q03247 | 2.08 | 1.31E-02 |
| O75936 | 2.08 | 8.81E-03 |
| P50570 | 2.08 | 7.62E-03 |
| Q8WWI5 | 2.05 | 3.23E-03 |
| Q9UQB8 | 2.05 | 9.62E-05 |
| P02748 | 2.04 | 5.53E-03 |
| Q8N386 | 2.04 | 8.58E-04 |
| P68371 | 2.02 | 3.65E-02 |
| P07602 | 2.01 | 9.80E-03 |
| Q9HBH0 | 2.01 | 1.56E-02 |
| P26038 | 2.00 | 1.73E-02 |
| Q92859 | 0.50 | 3.09E-02 |

| | | |
|---|---|---|
| P11047 | 0.50 | 4.10E-02 |
| Q07507 | 0.47 | 1.92E-02 |
| P07942 | 0.47 | 2.45E-02 |
| Q9UBG0 | 0.45 | 1.69E-02 |
| P13598 | 0.41 | 3.50E-02 |
| P04216 | 0.40 | 6.05E-03 |
| P09619 | 0.38 | 1.10E-02 |
| Q06418 | 0.38 | 3.50E-02 |
| P04180 | 0.32 | 1.39E-02 |
| O43556 | 0.29 | 2.27E-02 |
| P43652 | 0.28 | 4.17E-02 |
| P26992 | 0.26 | 3.76E-02 |
| P43121 | 0.22 | 1.47E-03 |
| O75339 | 0.20 | 2.06E-02 |
| Q96PD5 | 0.13 | 3.18E-02 |
| Q9Y6N8 | 0.12 | 1.73E-02 |

| F8 | | |
|---|---|---|
| **Protein Accessions** | **Fold Change** | **P value** |
| P06454 | 431.16 | 8.95E-13 |
| O75368 | 170.21 | 4.72E-11 |
| P62328 | 161.55 | 1.97E-06 |
| O60234 | 123.71 | 1.06E-14 |
| P07737 | 121.54 | 6.77E-07 |
| P00915 | 118.58 | 6.20E-10 |
| Q9P1F3 | 113.57 | 8.95E-13 |
| P52272 | 96.31 | 1.18E-06 |
| Q9Y547 | 81.74 | 2.83E-07 |
| P35754 | 58.11 | 8.60E-12 |
| O00244 | 57.79 | 1.80E-09 |
| Q96H15 | 50.15 | 1.48E-09 |
| O95861 | 49.71 | 1.01E-10 |
| Q14019 | 46.04 | 6.20E-10 |
| Q9UHY7 | 44.40 | 2.40E-08 |
| O75608 | 41.10 | 2.23E-08 |
| Q9NWV4 | 38.72 | 6.09E-11 |
| P39687 | 35.01 | 1.27E-09 |
| P07360 | 33.60 | 1.01E-10 |
| P84090 | 31.14 | 6.20E-10 |
| Q9H3K6 | 31.04 | 3.19E-08 |
| Q8IX05 | 27.35 | 4.06E-08 |
| P61769 | 25.81 | 3.22E-08 |
| P30050 | 24.65 | 1.22E-08 |
| Q9Y279 | 23.24 | 6.45E-08 |
| P26368 | 22.34 | 1.80E-09 |
| P36639 | 21.85 | 5.22E-09 |
| Q9NZT2 | 21.04 | 6.09E-09 |
| P08833 | 20.85 | 1.10E-06 |
| Q93091 | 19.91 | 9.56E-08 |
| P01589 | 19.60 | 1.55E-07 |
| P13640 | 18.89 | 5.14E-08 |
| P02763 | 18.63 | 2.59E-09 |
| Q9Y5Y7 | 18.62 | 4.69E-09 |
| Q9Y6X5 | 18.41 | 8.85E-08 |
| P62993 | 17.91 | 5.24E-10 |
| O43396 | 17.89 | 4.19E-09 |
| Q8IWT0 | 17.89 | 1.06E-06 |
| Q9ULZ3 | 17.74 | 1.16E-06 |
| P29966 | 17.62 | 8.72E-07 |
| P01717 | 17.18 | 4.39E-08 |
| P60983 | 16.81 | 4.17E-09 |
| O75531 | 16.78 | 1.06E-06 |
| Q9Y4L1 | 16.52 | 4.73E-08 |
| P05534 | 16.49 | 4.04E-09 |
| Q9Y490 | 16.43 | 1.87E-07 |

| | | |
|---|---|---|
| Q969H8 | 15.14 | 5.73E-07 |
| Q9H299 | 15.11 | 1.97E-06 |
| O95450 | 14.93 | 3.32E-07 |
| Q86WQ0 | 14.87 | 1.80E-08 |
| P12104 | 14.86 | 5.26E-08 |
| P0CG47 | 14.70 | 1.35E-08 |
| Q92530 | 14.69 | 4.69E-09 |
| Q9BTM9 | 14.47 | 1.38E-06 |
| Q13113 | 14.09 | 1.97E-09 |
| Q8WU39 | 13.93 | 1.13E-08 |
| Q02790 | 13.78 | 4.30E-09 |
| P28070 | 13.74 | 2.55E-09 |
| P30508 | 13.71 | 1.35E-08 |
| Q16661 | 13.69 | 2.17E-05 |
| Q15293 | 13.60 | 1.30E-07 |
| O43491 | 13.55 | 6.12E-07 |
| P67809 | 13.55 | 4.27E-07 |
| P00441 | 13.22 | 4.19E-09 |
| Q9NRX4 | 13.15 | 1.07E-07 |
| P00995 | 13.10 | 4.88E-07 |
| Q9BZM5 | 13.06 | 2.09E-06 |
| P18465 | 12.95 | 5.39E-08 |
| Q8TDQ1 | 12.66 | 5.37E-08 |
| Q6UWV6 | 12.20 | 2.88E-04 |
| Q16186 | 12.09 | 1.62E-07 |
| P02776 | 11.83 | 1.18E-06 |
| P26641 | 11.80 | 4.46E-07 |
| Q9H8J5 | 11.62 | 4.69E-09 |
| P02753 | 11.55 | 2.13E-07 |
| Q92954 | 11.55 | 1.48E-06 |
| Q9UBR2 | 11.37 | 4.23E-11 |
| Q15121 | 11.23 | 1.62E-07 |
| P56537 | 11.21 | 3.03E-11 |
| P80723 | 10.89 | 7.84E-06 |
| Q9BRT3 | 10.79 | 9.87E-08 |
| Q92743 | 10.52 | 9.01E-06 |
| P20333 | 10.50 | 6.39E-07 |
| Q496F6 | 10.47 | 4.38E-07 |
| Q8NHJ6 | 10.44 | 1.22E-08 |
| P07910 | 10.41 | 4.12E-07 |
| P55884 | 10.40 | 9.86E-04 |
| P01704 | 10.34 | 3.74E-04 |
| P02461 | 10.32 | 7.71E-06 |
| Q6UWR7 | 10.25 | 3.30E-07 |
| Q8TCD5 | 10.21 | 1.67E-08 |
| Q14515 | 10.03 | 1.37E-06 |
| P14207 | 10.01 | 8.85E-08 |
| P36269 | 9.96 | 1.87E-06 |

| | | |
|---|---|---|
| P83110 | 9.92 | 1.86E-07 |
| Q13421 | 9.91 | 1.48E-06 |
| P13796 | 9.77 | 4.46E-07 |
| P15814 | 9.75 | 3.58E-04 |
| P0DOX3 | 9.69 | 1.45E-05 |
| P0DOX8 | 9.69 | 4.06E-08 |
| O43556 | 9.66 | 1.48E-09 |
| P02792 | 9.61 | 7.86E-07 |
| Q8NCW5 | 9.52 | 1.08E-05 |
| Q9BVM4 | 9.44 | 4.01E-05 |
| Q92484 | 9.27 | 4.27E-06 |
| P63208 | 9.21 | 5.46E-03 |
| Q9NY33 | 9.19 | 3.06E-06 |
| P04118 | 9.10 | 3.74E-04 |
| P07108 | 8.96 | 1.34E-06 |
| Q1EHB4 | 8.83 | 1.45E-04 |
| O95336 | 8.67 | 1.59E-07 |
| Q96GD0 | 8.66 | 1.87E-07 |
| P02750 | 8.48 | 3.63E-07 |
| P0CF74 | 8.45 | 1.82E-05 |
| P36980 | 8.45 | 9.23E-07 |
| P01700 | 8.34 | 1.79E-04 |
| Q9H3Z4 | 8.31 | 2.67E-09 |
| P62253 | 8.10 | 3.94E-06 |
| A0A0C4DH72 | 8.04 | 2.62E-05 |
| P55058 | 7.99 | 1.78E-07 |
| Q00839 | 7.99 | 1.86E-07 |
| P80404 | 7.98 | 9.56E-08 |
| Q6UXN8 | 7.93 | 6.45E-08 |
| P50440 | 7.90 | 5.31E-07 |
| Q12765 | 7.83 | 1.57E-06 |
| A0A075B6K4 | 7.74 | 2.44E-04 |
| P17900 | 7.71 | 2.04E-05 |
| P05413 | 7.69 | 1.44E-06 |
| P25942 | 7.68 | 1.40E-06 |
| P00751 | 7.67 | 1.49E-03 |
| P78310 | 7.66 | 1.57E-06 |
| P0DOY3 | 7.55 | 1.06E-07 |
| P12111 | 7.44 | 4.34E-06 |
| Q96HD9 | 7.43 | 5.32E-06 |
| Q9Y5Z4 | 7.41 | 7.86E-07 |
| P07686 | 7.29 | 1.99E-07 |
| Q9GZP4 | 7.28 | 5.17E-05 |
| P28300 | 7.23 | 2.47E-05 |
| Q16822 | 7.19 | 1.06E-04 |
| O43399 | 7.16 | 3.06E-06 |
| Q14210 | 7.15 | 4.06E-03 |
| P01714 | 7.07 | 1.09E-06 |

| | | |
|---|---|---|
| Q9NUM4 | 7.02 | 2.84E-06 |
| Q07075 | 6.86 | 1.05E-04 |
| O14618 | 6.85 | 6.45E-08 |
| Q8N2U0 | 6.81 | 6.33E-05 |
| O96033 | 6.79 | 8.64E-06 |
| A0A075B6K5 | 6.79 | 3.56E-05 |
| P55957 | 6.73 | 1.87E-08 |
| Q13740 | 6.69 | 1.87E-07 |
| P12318 | 6.63 | 1.33E-05 |
| P00742 | 6.62 | 8.55E-07 |
| P19652 | 6.58 | 2.45E-06 |
| Q9Y696 | 6.53 | 2.71E-06 |
| Q16531 | 6.49 | 2.63E-04 |
| P04211 | 6.47 | 8.98E-07 |
| P09382 | 6.36 | 3.27E-06 |
| P11684 | 6.31 | 3.59E-03 |
| Q02487 | 6.31 | 1.03E-06 |
| P61970 | 6.29 | 1.68E-07 |
| P25311 | 6.28 | 7.89E-05 |
| Q9UHI7 | 6.27 | 4.57E-07 |
| Q8NFQ8 | 6.18 | 4.87E-06 |
| Q99988 | 6.17 | 1.34E-04 |
| A0A075B6S5 | 6.12 | 1.75E-04 |
| Q08431 | 6.10 | 1.87E-06 |
| O00754 | 6.01 | 8.98E-08 |
| P01599 | 5.96 | 7.02E-05 |
| P01624 | 5.91 | 2.80E-05 |
| P01911 | 5.90 | 3.73E-06 |
| A0PJK1 | 5.88 | 2.77E-06 |
| P09668 | 5.81 | 3.55E-06 |
| P30047 | 5.66 | 8.21E-06 |
| P19320 | 5.60 | 1.10E-04 |
| Q10567 | 5.59 | 8.47E-04 |
| P04066 | 5.56 | 2.62E-06 |
| P07306 | 5.55 | 5.52E-05 |
| O14745 | 5.54 | 2.97E-06 |
| Q7Z7D3 | 5.53 | 3.75E-05 |
| Q5T2W1 | 5.52 | 2.07E-07 |
| P02452 | 5.51 | 3.41E-05 |
| Q96JY6 | 5.41 | 1.19E-04 |
| P19438 | 5.27 | 4.46E-07 |
| P30101 | 5.25 | 1.79E-05 |
| Q9HD89 | 5.25 | 3.68E-04 |
| P25774 | 5.24 | 1.56E-06 |
| P02656 | 5.22 | 7.02E-05 |
| A0A075B6I0 | 5.19 | 3.41E-05 |
| P16422 | 5.16 | 2.54E-03 |
| P01699 | 5.10 | 1.67E-06 |

| | | |
|---|---|---|
| Q86VB7 | 5.06 | 2.22E-06 |
| Q96HE7 | 5.05 | 1.46E-02 |
| P01715 | 5.04 | 1.08E-06 |
| P50225 | 5.04 | 3.20E-03 |
| P46108 | 5.00 | 1.99E-04 |
| P07148 | 4.97 | 7.81E-06 |
| P35558 | 4.85 | 4.16E-06 |
| P01701 | 4.83 | 1.64E-05 |
| P48551 | 4.80 | 1.06E-04 |
| Q15517 | 4.80 | 1.97E-05 |
| P22897 | 4.80 | 1.20E-04 |
| Q8N357 | 4.78 | 8.22E-06 |
| P18428 | 4.77 | 2.74E-06 |
| P0DOX7 | 4.75 | 3.56E-05 |
| Q07812 | 4.67 | 4.65E-05 |
| Q16563 | 4.64 | 2.89E-05 |
| P09327 | 4.62 | 1.40E-05 |
| P35080 | 4.61 | 8.07E-06 |
| O96009 | 4.59 | 1.45E-05 |
| Q9BRA2 | 4.57 | 1.44E-06 |
| Q8NCC3 | 4.53 | 4.27E-06 |
| P07339 | 4.53 | 3.63E-07 |
| P01860 | 4.52 | 1.58E-03 |
| Q99653 | 4.49 | 6.69E-06 |
| Q6ZNA5 | 4.49 | 4.27E-06 |
| P01721 | 4.49 | 5.60E-05 |
| Q15185 | 4.48 | 1.08E-05 |
| P29279 | 4.47 | 5.69E-06 |
| Q9Y6H1 | 4.47 | 1.52E-04 |
| P02788 | 4.46 | 2.64E-02 |
| P49913 | 4.45 | 1.74E-04 |
| Q96IU4 | 4.45 | 8.42E-05 |
| Q9UM22 | 4.30 | 2.74E-06 |
| Q6ZVN8 | 4.29 | 1.58E-04 |
| P01619 | 4.27 | 9.44E-04 |
| Q9NS15 | 4.23 | 7.33E-04 |
| Q9UK55 | 4.23 | 7.78E-04 |
| P08123 | 4.23 | 3.06E-06 |
| P02741 | 4.20 | 2.52E-05 |
| P39060 | 4.18 | 3.93E-07 |
| P02794 | 4.18 | 5.14E-03 |
| P18084 | 4.17 | 1.06E-04 |
| P17405 | 4.17 | 7.86E-07 |
| P20774 | 4.16 | 3.96E-05 |
| A0A0A0MRZ8 | 4.14 | 7.55E-05 |
| O00499 | 4.13 | 2.92E-05 |
| A0A075B6J9 | 4.11 | 7.46E-03 |
| Q9BS26 | 4.11 | 1.88E-06 |

| | | |
|---|---|---|
| P08237 | 4.10 | 4.24E-05 |
| Q03405 | 4.10 | 5.71E-05 |
| P54727 | 4.10 | 1.37E-05 |
| P04626 | 4.09 | 6.78E-03 |
| Q9UJ14 | 4.08 | 7.13E-05 |
| P16401 | 4.06 | 2.08E-02 |
| A0A0B4J1Y8 | 4.06 | 1.63E-02 |
| Q15847 | 4.06 | 4.78E-04 |
| P00505 | 4.06 | 7.83E-04 |
| A0A0C4DH68 | 4.03 | 1.08E-03 |
| P08238 | 4.02 | 1.16E-06 |
| Q8IWY4 | 4.01 | 2.30E-04 |
| Q9BTY2 | 4.01 | 9.25E-06 |
| Q7L5L3 | 4.00 | 1.35E-03 |
| P23297 | 3.99 | 2.69E-03 |
| P04080 | 3.97 | 1.87E-02 |
| P07858 | 3.94 | 4.65E-05 |
| P15586 | 3.93 | 5.47E-07 |
| Q6GTX8 | 3.93 | 1.37E-06 |
| P28799 | 3.90 | 3.26E-04 |
| Q5KU26 | 3.88 | 1.44E-02 |
| O00584 | 3.88 | 2.10E-05 |
| P13693 | 3.88 | 1.22E-03 |
| Q9NP84 | 3.87 | 4.38E-02 |
| Q9Y274 | 3.85 | 1.00E-03 |
| Q96TA1 | 3.79 | 1.87E-02 |
| P13797 | 3.76 | 2.00E-03 |
| P06865 | 3.76 | 7.93E-06 |
| Q16581 | 3.74 | 3.27E-06 |
| P12259 | 3.73 | 2.53E-06 |
| Q9Y6R1 | 3.70 | 4.44E-04 |
| P01861 | 3.70 | 1.01E-02 |
| P26572 | 3.70 | 4.62E-03 |
| Q695T7 | 3.68 | 2.49E-04 |
| O75022 | 3.67 | 8.42E-05 |
| P08571 | 3.67 | 2.22E-04 |
| P15529 | 3.66 | 3.60E-06 |
| P81605 | 3.66 | 9.27E-04 |
| Q969Z4 | 3.65 | 1.64E-05 |
| P01703 | 3.59 | 2.01E-04 |
| Q02747 | 3.59 | 4.87E-03 |
| P14209 | 3.58 | 1.77E-02 |
| O43155 | 3.58 | 3.78E-03 |
| P27824 | 3.56 | 1.09E-03 |
| Q6E0U4 | 3.55 | 6.35E-03 |
| P27348 | 3.49 | 1.10E-04 |
| A6NDG6 | 3.49 | 2.10E-02 |
| P13667 | 3.48 | 2.00E-05 |

| | | |
|---|---|---|
| P23083 | 3.46 | 2.20E-02 |
| P27487 | 3.45 | 3.24E-05 |
| Q15084 | 3.44 | 6.53E-04 |
| P01593 | 3.44 | 5.29E-04 |
| ENSEMBL:ENSBTAP00000007350 | 3.42 | 9.43E-04 |
| P37840 | 3.39 | 4.25E-03 |
| P15374 | 3.39 | 6.89E-06 |
| Q86YD5 | 3.38 | 3.82E-03 |
| A0A0J9YXX1 | 3.38 | 8.45E-05 |
| P21980 | 3.38 | 4.33E-02 |
| Q96IQ7 | 3.38 | 2.23E-02 |
| P08779 | 3.37 | 2.54E-02 |
| P14780 | 3.35 | 3.00E-02 |
| P35606 | 3.35 | 6.53E-04 |
| O00299 | 3.34 | 1.71E-05 |
| Q9Y275 | 3.34 | 3.40E-04 |
| P11279 | 3.33 | 2.01E-04 |
| Q92542 | 3.32 | 1.64E-03 |
| P62195 | 3.32 | 1.09E-02 |
| P12814 | 3.32 | 2.03E-05 |
| P31947 | 3.31 | 3.69E-02 |
| Q15435 | 3.29 | 1.40E-03 |
| A0A087WSX0 | 3.29 | 4.27E-03 |
| Q14847 | 3.28 | 4.26E-05 |
| Q15907 | 3.26 | 3.76E-05 |
| Q15833 | 3.25 | 1.22E-03 |
| P22004 | 3.23 | 6.14E-04 |
| Q15828 | 3.21 | 8.83E-03 |
| Q9BXN2 | 3.19 | 1.45E-05 |
| Q13621 | 3.14 | 5.79E-05 |
| O00182 | 3.14 | 5.37E-05 |
| P02511 | 3.14 | 5.06E-04 |
| Q8WWV6 | 3.12 | 3.84E-04 |
| Q5JXA9 | 3.12 | 2.10E-05 |
| Q99650 | 3.10 | 6.88E-04 |
| P01344 | 3.08 | 1.05E-04 |
| P43652 | 3.08 | 6.82E-05 |
| O00264 | 3.08 | 4.58E-05 |
| P15144 | 3.07 | 8.36E-03 |
| A6NI73 | 3.04 | 1.45E-04 |
| P36941 | 3.03 | 6.69E-03 |
| P10619 | 3.03 | 6.82E-05 |
| Q01151 | 3.02 | 2.72E-02 |
| P35527 | 3.02 | 4.15E-02 |
| P06681 | 3.01 | 8.67E-03 |
| P00740 | 3.01 | 5.76E-03 |
| Q15223 | 3.00 | 4.07E-06 |
| P78325 | 2.96 | 1.79E-02 |

| | | |
|---|---|---|
| O43405 | 2.95 | 7.09E-03 |
| P35237 | 2.94 | 2.18E-03 |
| P36957 | 2.93 | 1.29E-03 |
| Q8N386 | 2.93 | 1.08E-04 |
| Q9Y3B3 | 2.93 | 3.51E-05 |
| O15540 | 2.92 | 3.28E-02 |
| P37802 | 2.92 | 8.52E-04 |
| Q9BUP0 | 2.90 | 3.03E-03 |
| Q9BUH6 | 2.89 | 9.85E-04 |
| Q9HAT2 | 2.86 | 1.09E-04 |
| A0A075B6Q5 | 2.83 | 1.31E-03 |
| P17050 | 2.82 | 1.72E-04 |
| Q9NPY3 | 2.82 | 6.68E-04 |
| Q9UNF0 | 2.81 | 2.16E-04 |
| P49006 | 2.81 | 1.06E-02 |
| P30044 | 2.79 | 1.55E-04 |
| Q9HB90 | 2.79 | 2.79E-02 |
| P07359 | 2.78 | 9.27E-04 |
| O15394 | 2.78 | 2.97E-02 |
| Q8WWB7 | 2.78 | 1.90E-04 |
| Q93050 | 2.78 | 1.11E-04 |
| Q96CX2 | 2.76 | 8.25E-04 |
| P30043 | 2.75 | 3.03E-03 |
| Q13291 | 2.75 | 5.23E-03 |
| P98172 | 2.75 | 2.61E-04 |
| Q13332 | 2.74 | 1.44E-02 |
| P24592 | 2.74 | 1.96E-02 |
| Q96C19 | 2.73 | 7.68E-03 |
| Q13361 | 2.73 | 2.19E-03 |
| Q02083 | 2.73 | 2.37E-02 |
| Q15904 | 2.71 | 2.80E-03 |
| A0A0B4J2D9 | 2.70 | 2.57E-02 |
| P04179 | 2.70 | 1.58E-02 |
| P02790 | 2.69 | 7.15E-05 |
| P00167 | 2.68 | 5.97E-04 |
| P00450 | 2.68 | 1.66E-03 |
| P26038 | 2.66 | 2.91E-04 |
| P07602 | 2.66 | 1.60E-04 |
| Q8NHP8 | 2.66 | 3.56E-05 |
| P17936 | 2.66 | 2.38E-03 |
| Q12794 | 2.65 | 6.00E-03 |
| P62942 | 2.65 | 1.67E-02 |
| P36897 | 2.64 | 2.96E-03 |
| Q13200 | 2.63 | 2.22E-02 |
| Q92485 | 2.63 | 3.65E-03 |
| O75936 | 2.62 | 3.98E-04 |
| P0DOX5 | 2.59 | 8.81E-05 |
| Q5JS37 | 2.57 | 9.56E-04 |

| | | |
|---|---|---|
| A2NJV5 | 2.55 | 4.25E-03 |
| Q96MU8 | 2.55 | 1.92E-04 |
| Q92734 | 2.54 | 4.29E-03 |
| Q8NC42 | 2.53 | 1.06E-04 |
| Q14315 | 2.52 | 1.07E-03 |
| P54803 | 2.51 | 6.53E-04 |
| A0A0J9YX35 | 2.51 | 3.59E-03 |
| Q9H665 | 2.48 | 5.36E-04 |
| P21583 | 2.47 | 6.79E-03 |
| A0A075B6H9 | 2.46 | 3.97E-03 |
| P16234 | 2.45 | 2.35E-04 |
| Q7Z6A9 | 2.45 | 4.40E-02 |
| P06312 | 2.45 | 1.14E-02 |
| O14786 | 2.44 | 1.98E-02 |
| P06331 | 2.44 | 3.28E-03 |
| P30046 | 2.44 | 6.78E-03 |
| P61916 | 2.44 | 8.22E-06 |
| Q07837 | 2.43 | 2.74E-03 |
| Q9Y646 | 2.42 | 1.28E-03 |
| O95084 | 2.42 | 3.61E-02 |
| P07237 | 2.41 | 4.37E-02 |
| Q99795 | 2.40 | 3.60E-04 |
| Q9BXS4 | 2.40 | 6.10E-03 |
| P48960 | 2.39 | 3.11E-02 |
| O75503 | 2.39 | 1.92E-04 |
| Q92520 | 2.38 | 6.33E-05 |
| P01229 | 2.37 | 5.01E-04 |
| Q9NQW7 | 2.36 | 5.46E-03 |
| P15311 | 2.36 | 1.84E-03 |
| Q13183 | 2.35 | 8.69E-04 |
| Q8NHL6 | 2.35 | 5.44E-03 |
| Q9BVK6 | 2.33 | 1.69E-02 |
| O15204 | 2.33 | 3.18E-02 |
| Q02985 | 2.32 | 3.83E-04 |
| P06858 | 2.32 | 2.76E-03 |
| Q9HBR0 | 2.31 | 1.23E-04 |
| P51688 | 2.30 | 1.55E-02 |
| Q9HC35 | 2.29 | 2.24E-03 |
| P28838 | 2.29 | 1.03E-02 |
| Q96AP7 | 2.28 | 1.12E-02 |
| Q9H0U4 | 2.28 | 5.45E-03 |
| P25325 | 2.27 | 2.44E-03 |
| P21333 | 2.27 | 2.47E-03 |
| P22223 | 2.27 | 7.91E-03 |
| P50570 | 2.26 | 6.91E-03 |
| P41222 | 2.26 | 7.99E-03 |
| P20062 | 2.26 | 3.49E-03 |
| P05023 | 2.25 | 2.69E-02 |

| | | |
|---|---|---|
| Q9UNZ2 | 2.23 | 4.92E-02 |
| P01011 | 2.23 | 5.93E-03 |
| Q9Y6E0 | 2.21 | 3.80E-03 |
| Q9BYE9 | 2.21 | 4.80E-03 |
| O60814 | 2.21 | 6.20E-03 |
| Q9BSG0 | 2.20 | 2.24E-03 |
| P20933 | 2.20 | 3.67E-05 |
| P51570 | 2.20 | 1.02E-04 |
| Q15043 | 2.18 | 4.30E-03 |
| O00338 | 2.18 | 1.28E-03 |
| Q96LD4 | 2.17 | 1.95E-04 |
| TREMBL:Q3ZBS7 | 2.16 | 8.70E-03 |
| P46940 | 2.16 | 2.86E-02 |
| P01127 | 2.16 | 2.75E-02 |
| SWISS-PROT:P19012 | 2.14 | 7.35E-03 |
| Q9H008 | 2.14 | 3.13E-02 |
| P35241 | 2.13 | 2.05E-04 |
| Q13228 | 2.12 | 2.78E-03 |
| O15118 | 2.11 | 5.34E-04 |
| P20827 | 2.11 | 1.09E-03 |
| P28332 | 2.11 | 4.98E-02 |
| Q96GP6 | 2.08 | 4.87E-03 |
| O00468 | 2.07 | 4.70E-02 |
| P17980 | 2.07 | 9.91E-03 |
| P20138 | 2.07 | 2.69E-03 |
| O43776 | 2.05 | 2.83E-02 |
| P61006 | 2.05 | 1.42E-02 |
| A0A0C4DH38 | 2.04 | 2.29E-02 |
| A0A0A0MS15 | 2.03 | 1.89E-02 |
| Q9UJ99 | 2.03 | 3.09E-02 |
| Q9UNN8 | 0.50 | 3.26E-03 |
| Q9BQ51 | 0.49 | 1.55E-02 |
| Q6UX71 | 0.49 | 7.09E-03 |
| Q08174 | 0.48 | 4.95E-02 |
| Q01973 | 0.48 | 3.47E-02 |
| O14498 | 0.46 | 2.89E-02 |
| Q02413 | 0.45 | 4.86E-02 |
| P30530 | 0.45 | 1.01E-03 |
| P19022 | 0.44 | 2.86E-02 |
| Q7KYR7 | 0.43 | 9.98E-03 |
| O43633 | 0.42 | 3.14E-02 |
| Q9UN37 | 0.41 | 2.51E-02 |
| Q8WUM4 | 0.40 | 1.71E-02 |
| Q9Y653 | 0.40 | 4.49E-02 |
| P05155 | 0.40 | 3.61E-02 |
| P05067 | 0.40 | 4.40E-02 |
| Q04721 | 0.40 | 4.29E-02 |
| Q12864 | 0.40 | 2.13E-02 |

| | | |
|---|---|---|
| P04180 | 0.39 | 9.00E-03 |
| Q9ULK6 | 0.38 | 3.31E-02 |
| Q9HC56 | 0.38 | 4.95E-02 |
| P14784 | 0.37 | 2.43E-02 |
| O75144 | 0.37 | 7.33E-03 |
| P41217 | 0.35 | 1.80E-02 |
| P02768 | 0.35 | 3.98E-02 |
| Q08334 | 0.35 | 2.25E-02 |
| P25940 | 0.35 | 1.09E-02 |
| Q9UBG0 | 0.35 | 4.75E-03 |
| Q86YT9 | 0.35 | 1.78E-02 |
| Q8NBJ4 | 0.34 | 4.58E-02 |
| P19256 | 0.33 | 7.95E-03 |
| P0C7U0 | 0.33 | 1.91E-02 |
| Q86UX2 | 0.33 | 4.49E-02 |
| P02462 | 0.32 | 3.14E-02 |
| Q8N2G4 | 0.32 | 4.33E-02 |
| Q9NPF0 | 0.32 | 1.63E-02 |
| P13598 | 0.32 | 6.22E-03 |
| P50148 | 0.31 | 2.84E-02 |
| Q8N3J6 | 0.31 | 4.97E-02 |
| Q7Z7G0 | 0.31 | 3.41E-02 |
| Q5SZK8 | 0.30 | 5.44E-03 |
| P11362 | 0.30 | 2.69E-02 |
| Q9NQS3 | 0.30 | 4.26E-03 |
| P04746 | 0.29 | 4.79E-02 |
| P13591 | 0.29 | 1.58E-02 |
| Q6UXD5 | 0.29 | 4.38E-02 |
| Q6UXG3 | 0.29 | 1.01E-02 |
| Q8TBP5 | 0.29 | 1.60E-02 |
| Q96PD5 | 0.29 | 3.07E-02 |
| P48745 | 0.29 | 3.14E-02 |
| P24855 | 0.28 | 6.86E-03 |
| Q9P2B2 | 0.28 | 1.68E-02 |
| O75351 | 0.27 | 1.63E-02 |
| P09619 | 0.26 | 1.40E-03 |
| Q92859 | 0.26 | 1.40E-03 |
| O75339 | 0.26 | 1.13E-02 |
| P01042 | 0.26 | 3.47E-02 |
| Q13443 | 0.26 | 1.04E-02 |
| Q96RW7 | 0.25 | 1.67E-02 |
| P15151 | 0.25 | 7.23E-03 |
| Q9Y2G1 | 0.25 | 9.12E-03 |
| Q7LBR1 | 0.25 | 1.34E-02 |
| P08697 | 0.25 | 4.37E-02 |
| Q86XT2 | 0.25 | 4.33E-02 |
| Q12860 | 0.25 | 2.52E-02 |
| Q15262 | 0.24 | 5.44E-03 |

| | | |
|---|---|---|
| Q6EMK4 | 0.23 | 1.58E-02 |
| Q9NP85 | 0.23 | 1.10E-02 |
| Q9UK41 | 0.23 | 7.08E-03 |
| Q9BY67 | 0.23 | 2.69E-02 |
| Q96NY8 | 0.23 | 1.55E-02 |
| P31997 | 0.22 | 4.51E-02 |
| Q9UGT4 | 0.22 | 2.52E-02 |
| P11047 | 0.22 | 1.56E-03 |
| P16035 | 0.22 | 9.57E-03 |
| Q9H741 | 0.22 | 7.20E-03 |
| Q9HD42 | 0.21 | 3.86E-02 |
| Q86TH1 | 0.21 | 2.51E-02 |
| P16112 | 0.21 | 1.40E-02 |
| P02774 | 0.20 | 7.95E-03 |
| P26992 | 0.20 | 9.81E-03 |
| Q99835 | 0.19 | 7.49E-04 |
| Q6V0I7 | 0.19 | 1.80E-02 |
| Q9HBB8 | 0.19 | 1.63E-02 |
| Q8TB96 | 0.18 | 1.27E-03 |
| P35858 | 0.18 | 3.14E-02 |
| Q96JQ0 | 0.18 | 1.84E-03 |
| P98095 | 0.18 | 3.09E-02 |
| P29323 | 0.16 | 1.49E-04 |
| P12273 | 0.14 | 1.73E-02 |
| P04745 | 0.14 | 2.92E-02 |
| Q9H444 | 0.14 | 3.62E-02 |
| Q6UVK1 | 0.13 | 4.63E-02 |
| P23327 | 0.13 | 3.76E-02 |
| Q14574 | 0.13 | 3.44E-02 |
| P39059 | 0.13 | 4.38E-02 |
| Q9P121 | 0.12 | 3.61E-02 |
| Q8NDA2 | 0.12 | 1.01E-02 |
| Q14982 | 0.11 | 4.86E-02 |
| P55017 | 0.10 | 7.20E-03 |
| Q9HCU0 | 0.09 | 1.74E-03 |
| P80370 | 0.09 | 1.36E-02 |
| Q5JRA6 | 0.07 | 1.78E-02 |

| F9 | | |
|---|---|---|
| **Protein Accessions** | **Fold Change** | **P value** |
| P01225 | 29.30 | 1.51E-07 |
| P69905 | 15.70 | 7.37E-04 |
| P14780 | 6.99 | 3.16E-03 |
| P01215 | 5.81 | 1.67E-04 |
| P00915 | 5.75 | 1.13E-02 |
| P01229 | 5.16 | 2.27E-05 |
| P54793 | 4.65 | 1.74E-03 |
| Q8N2U0 | 4.47 | 1.96E-03 |
| P02792 | 4.13 | 9.95E-03 |
| P01700 | 4.12 | 9.78E-06 |
| O75015 | 3.34 | 4.97E-02 |
| P0DOX6 | 3.30 | 1.77E-03 |
| A0A0J9YX35 | 3.16 | 7.47E-03 |
| Q96LD4 | 3.05 | 3.98E-05 |
| P01871 | 3.02 | 6.45E-03 |
| P01911 | 3.01 | 1.82E-02 |
| Q7L5L3 | 2.98 | 2.44E-02 |
| P32119 | 2.93 | 2.82E-02 |
| P01859 | 2.88 | 2.32E-02 |
| A0A0C4DH68 | 2.77 | 6.45E-03 |
| A2NJV5 | 2.55 | 4.29E-02 |
| O94832 | 2.48 | 9.95E-03 |
| P01780 | 2.47 | 6.45E-03 |
| Q9HAU8 | 2.32 | 5.86E-03 |
| P15814 | 2.30 | 4.97E-02 |
| P0DOX8 | 2.25 | 4.87E-02 |
| P01011 | 2.24 | 4.97E-02 |
| P22748 | 2.24 | 2.51E-02 |
| Q13621 | 2.23 | 2.41E-02 |
| P00441 | 2.04 | 4.97E-02 |
| P01715 | 2.02 | 3.33E-02 |
| A0A0B4J1U7 | 2.01 | 3.72E-02 |
| Q6UXB4 | 0.47 | 2.96E-02 |
| Q9UNN8 | 0.47 | 2.75E-02 |
| P30530 | 0.43 | 1.13E-02 |
| Q14194 | 0.14 | 3.48E-02 |

| F10 | | |
|---|---|---|
| **Protein Accessions** | **Fold Change** | **P value** |
| Q15293 | 68.15 | 1.57E-09 |
| P61769 | 59.84 | 1.57E-09 |
| A0A0B4J2D9 | 38.05 | 9.33E-06 |
| P02750 | 32.50 | 1.19E-08 |
| P02763 | 25.51 | 1.57E-09 |
| P00915 | 21.52 | 2.59E-07 |
| P05534 | 19.25 | 1.57E-09 |
| P12104 | 18.94 | 1.52E-07 |
| P11684 | 18.37 | 7.31E-06 |
| O75368 | 18.06 | 7.35E-09 |
| Q9NWV4 | 15.74 | 6.23E-07 |
| A0A087WSX0 | 13.97 | 2.59E-07 |
| P19652 | 13.50 | 9.88E-07 |
| P20774 | 13.41 | 1.91E-08 |
| P01717 | 12.90 | 1.11E-08 |
| Q9UBR2 | 12.62 | 5.28E-10 |
| Q9P1F3 | 10.90 | 3.08E-07 |
| P01019 | 10.76 | 1.19E-08 |
| P00738 | 10.47 | 3.72E-05 |
| P18465 | 10.14 | 4.00E-08 |
| P25311 | 10.09 | 5.80E-06 |
| P55103 | 9.95 | 8.36E-07 |
| P27824 | 9.87 | 1.38E-06 |
| P30508 | 9.69 | 2.41E-06 |
| P14207 | 8.91 | 5.46E-05 |
| Q9NR99 | 8.86 | 1.51E-02 |
| Q9UK55 | 8.36 | 1.16E-05 |
| P01703 | 8.33 | 7.98E-07 |
| P08833 | 8.32 | 2.04E-04 |
| Q969H8 | 8.26 | 1.16E-05 |
| Q99972 | 8.10 | 7.98E-07 |
| Q92530 | 8.09 | 5.99E-07 |
| P19320 | 7.92 | 1.87E-05 |
| P22692 | 7.64 | 2.09E-02 |
| O00244 | 7.61 | 2.01E-04 |
| A0A0C4DH72 | 7.40 | 1.38E-04 |
| Q9UHI8 | 7.40 | 2.55E-05 |

| | | |
|---|---|---|
| Q15435 | 7.34 | 2.62E-05 |
| Q5JTV8 | 7.33 | 2.06E-06 |
| P01011 | 7.27 | 9.84E-07 |
| P01714 | 7.19 | 7.23E-06 |
| P03951 | 7.07 | 4.00E-05 |
| A0A075B6K5 | 6.83 | 7.36E-05 |
| Q13421 | 6.60 | 7.41E-05 |
| P07359 | 6.57 | 1.44E-06 |
| P05413 | 6.54 | 1.56E-05 |
| Q9H8J5 | 6.47 | 7.23E-06 |
| Q16661 | 6.47 | 6.60E-03 |
| P33151 | 6.28 | 2.09E-04 |
| O00754 | 6.22 | 4.49E-07 |
| P01903 | 6.19 | 3.72E-02 |
| Q9UHY7 | 6.16 | 3.25E-06 |
| P01599 | 6.10 | 6.76E-05 |
| Q9UBP4 | 6.06 | 6.66E-05 |
| P0DOX8 | 5.87 | 4.48E-06 |
| Q9UHL4 | 5.86 | 6.97E-05 |
| Q9NUM4 | 5.80 | 1.38E-04 |
| P01008 | 5.77 | 4.37E-07 |
| P36980 | 5.57 | 2.91E-05 |
| O43556 | 5.55 | 2.59E-07 |
| P01721 | 5.49 | 8.31E-05 |
| P27797 | 5.43 | 2.73E-04 |
| Q93091 | 5.41 | 5.69E-05 |
| P53634 | 5.34 | 2.59E-07 |
| P07360 | 5.32 | 1.20E-05 |
| P21583 | 5.32 | 2.27E-05 |
| O95980 | 5.23 | 5.19E-03 |
| P15814 | 5.21 | 5.55E-06 |
| P02787 | 5.18 | 8.02E-05 |
| A0A075B6S5 | 5.14 | 5.72E-05 |
| P02741 | 5.13 | 1.38E-06 |
| Q8IX05 | 5.11 | 1.73E-05 |
| P02655 | 5.09 | 4.22E-05 |
| A0A075B6Q5 | 5.05 | 3.95E-03 |
| Q92954 | 5.04 | 1.71E-03 |
| P02790 | 5.03 | 1.71E-06 |

| | | |
|---|---|---|
| A0A075B6I0 | 5.01 | 2.67E-03 |
| P31151 | 4.99 | 9.88E-03 |
| O00468 | 4.98 | 1.53E-03 |
| A0A075B6K4 | 4.98 | 4.28E-04 |
| P01715 | 4.93 | 2.64E-06 |
| P17900 | 4.89 | 1.24E-03 |
| Q9Y6X5 | 4.86 | 2.62E-05 |
| P08185 | 4.79 | 9.84E-07 |
| P02656 | 4.77 | 3.00E-04 |
| P18428 | 4.68 | 8.20E-06 |
| P09382 | 4.67 | 3.12E-04 |
| P20142 | 4.67 | 4.00E-03 |
| P43652 | 4.67 | 3.25E-06 |
| P01911 | 4.63 | 5.90E-05 |
| A0A0C4DH68 | 4.60 | 9.05E-06 |
| Q86SG5 | 4.59 | 2.30E-02 |
| Q13740 | 4.44 | 2.64E-06 |
| Q9Y547 | 4.43 | 3.20E-03 |
| P0C0L4 | 4.29 | 1.93E-05 |
| P02748 | 4.29 | 3.70E-06 |
| P04217 | 4.28 | 8.34E-04 |
| O76076 | 4.26 | 1.95E-03 |
| P01860 | 4.24 | 4.70E-03 |
| P02792 | 4.20 | 1.10E-03 |
| P04211 | 4.17 | 2.52E-05 |
| P06312 | 4.12 | 2.74E-04 |
| P04216 | 4.06 | 2.59E-07 |
| Q8N2U0 | 4.04 | 4.47E-04 |
| Q6UXB8 | 3.95 | 7.94E-04 |
| Q9UDY2 | 3.92 | 1.17E-03 |
| Q14376 | 3.92 | 1.15E-03 |
| P36969 | 3.90 | 1.27E-03 |
| Q96FW1 | 3.87 | 4.64E-03 |
| Q9H299 | 3.81 | 1.94E-02 |
| Q9BTY2 | 3.80 | 3.78E-05 |
| Q99988 | 3.79 | 9.81E-03 |
| O00584 | 3.78 | 1.80E-04 |
| O00499 | 3.77 | 1.99E-04 |
| P17050 | 3.76 | 5.70E-05 |

| | | |
|---|---|---|
| P01619 | 3.74 | 4.64E-03 |
| P00441 | 3.74 | 3.13E-05 |
| P00747 | 3.73 | 2.82E-04 |
| P01701 | 3.73 | 2.34E-04 |
| Q8N126 | 3.70 | 5.42E-04 |
| P08571 | 3.69 | 5.98E-04 |
| P01699 | 3.66 | 3.86E-05 |
| P12318 | 3.66 | 5.35E-03 |
| Q14515 | 3.65 | 3.35E-02 |
| Q92743 | 3.65 | 2.11E-03 |
| P07306 | 3.64 | 3.95E-03 |
| P29966 | 3.58 | 3.97E-02 |
| P09228 | 3.58 | 3.21E-03 |
| A0A075B6J9 | 3.57 | 5.19E-03 |
| O00115 | 3.54 | 5.23E-05 |
| P25774 | 3.50 | 1.38E-04 |
| P01009 | 3.48 | 3.46E-05 |
| Q8NCW5 | 3.47 | 1.27E-03 |
| P02753 | 3.47 | 3.61E-04 |
| Q07075 | 3.47 | 2.38E-02 |
| SWISS-PROT:P02769 | 3.45 | 2.46E-02 |
| A0A0A0MRZ8 | 3.42 | 1.07E-02 |
| P01700 | 3.41 | 8.20E-06 |
| P01624 | 3.41 | 3.01E-03 |
| Q92765 | 3.40 | 1.43E-02 |
| Q9HAU8 | 3.40 | 1.29E-04 |
| Q9BZM5 | 3.38 | 2.59E-03 |
| Q92520 | 3.37 | 4.48E-06 |
| Q9UM22 | 3.37 | 9.01E-05 |
| O43491 | 3.36 | 3.02E-03 |
| Q9BW04 | 3.35 | 7.05E-03 |
| Q8NCC3 | 3.27 | 2.41E-04 |
| P37840 | 3.24 | 3.41E-03 |
| P00742 | 3.20 | 1.13E-03 |
| P35080 | 3.19 | 4.96E-03 |
| P21217 | 3.19 | 4.47E-04 |
| P28300 | 3.18 | 1.01E-03 |
| P23470 | 3.17 | 2.03E-05 |
| P14209 | 3.16 | 4.82E-02 |

| | | |
|---|---|---|
| Q02985 | 3.16 | 6.46E-05 |
| P00450 | 3.16 | 7.56E-04 |
| Q15113 | 3.10 | 1.10E-03 |
| Q14315 | 3.10 | 4.20E-04 |
| P02760 | 3.10 | 2.14E-03 |
| Q96IU4 | 3.08 | 3.20E-03 |
| Q9Y279 | 3.07 | 1.59E-02 |
| Q8IWY4 | 3.06 | 4.64E-03 |
| P06331 | 3.03 | 9.30E-04 |
| Q6UXN8 | 3.02 | 1.52E-02 |
| O15540 | 3.01 | 3.72E-02 |
| P78325 | 3.01 | 1.94E-02 |
| Q8NFQ8 | 3.00 | 1.10E-02 |
| P0DOX7 | 3.00 | 2.14E-03 |
| Q96FN5 | 3.00 | 2.61E-03 |
| O60234 | 2.98 | 3.87E-03 |
| Q15274 | 2.97 | 9.83E-03 |
| P00740 | 2.95 | 2.12E-03 |
| P09668 | 2.93 | 2.71E-04 |
| O96009 | 2.93 | 1.71E-03 |
| O15204 | 2.92 | 4.61E-02 |
| P12111 | 2.92 | 6.69E-03 |
| P01344 | 2.91 | 4.14E-04 |
| A0A0C4DH25 | 2.91 | 1.00E-02 |
| Q96C19 | 2.90 | 8.06E-03 |
| P0DOY3 | 2.89 | 9.27E-03 |
| A0A0B4J1U7 | 2.87 | 2.18E-04 |
| P07858 | 2.87 | 5.42E-04 |
| Q9NZ08 | 2.87 | 2.66E-03 |
| SWISS-PROT:P00978 | 2.84 | 5.19E-03 |
| Q9H0U4 | 2.83 | 1.35E-02 |
| Q9Y4L1 | 2.83 | 1.80E-02 |
| A2NJV5 | 2.82 | 3.40E-03 |
| P21796 | 2.81 | 2.46E-03 |
| Q5T0N5 | 2.79 | 1.88E-02 |
| P54793 | 2.79 | 7.05E-03 |
| Q9Y5Z4 | 2.79 | 2.83E-03 |
| P51688 | 2.79 | 5.72E-03 |
| P54803 | 2.75 | 2.74E-04 |

| | | |
|---|---|---|
| P48551 | 2.74 | 1.51E-02 |
| Q9H6B4 | 2.74 | 2.83E-03 |
| P31025 | 2.72 | 1.29E-04 |
| Q92484 | 2.72 | 2.29E-02 |
| SWISS-PROT:P07724 | 2.71 | 5.57E-03 |
| P04233 | 2.70 | 1.69E-02 |
| P10619 | 2.70 | 4.86E-04 |
| A0A0C4DH38 | 2.70 | 4.64E-03 |
| Q8N114 | 2.68 | 2.82E-02 |
| Q92496 | 2.65 | 1.76E-03 |
| P23083 | 2.64 | 4.64E-03 |
| Q6YHK3 | 2.63 | 9.30E-04 |
| P07148 | 2.62 | 3.99E-03 |
| P83110 | 2.62 | 6.49E-03 |
| P07737 | 2.61 | 3.61E-04 |
| P19438 | 2.61 | 5.42E-04 |
| Q15155 | 2.60 | 4.25E-03 |
| Q6UWV6 | 2.58 | 4.00E-02 |
| Q14165 | 2.54 | 2.31E-03 |
| P78310 | 2.54 | 1.44E-02 |
| P05091 | 2.51 | 1.18E-02 |
| Q13361 | 2.51 | 1.90E-03 |
| Q9Y646 | 2.51 | 1.74E-03 |
| Q7L5L3 | 2.50 | 1.71E-02 |
| O75629 | 2.49 | 1.08E-02 |
| Q14019 | 2.49 | 1.23E-02 |
| P24592 | 2.48 | 4.57E-02 |
| Q96CX2 | 2.48 | 6.03E-03 |
| Q15121 | 2.47 | 3.36E-02 |
| Q9H6X2 | 2.46 | 2.19E-02 |
| Q9BVK6 | 2.42 | 2.54E-02 |
| P15848 | 2.41 | 5.19E-03 |
| P02671 | 2.41 | 1.47E-02 |
| Q9UGM5 | 2.41 | 3.03E-02 |
| P26641 | 2.39 | 3.54E-02 |
| Q16563 | 2.38 | 1.53E-02 |
| P35754 | 2.37 | 5.00E-02 |
| P67809 | 2.35 | 2.45E-02 |
| Q00325 | 2.35 | 1.20E-02 |

| | | |
|---|---|---|
| Q9Y6R1 | 2.34 | 2.86E-02 |
| P15586 | 2.33 | 4.37E-04 |
| Q14126 | 2.32 | 8.49E-04 |
| P01229 | 2.32 | 1.35E-02 |
| Q9BTM9 | 2.32 | 2.35E-02 |
| O94919 | 2.31 | 2.85E-02 |
| Q14112 | 2.31 | 1.39E-02 |
| A0A0A0MS15 | 2.30 | 1.06E-02 |
| Q13057 | 2.29 | 4.62E-02 |
| P55058 | 2.28 | 5.69E-03 |
| Q8NHJ6 | 2.27 | 5.98E-03 |
| O00264 | 2.27 | 2.31E-03 |
| P01764 | 2.27 | 2.93E-02 |
| Q8NHP8 | 2.26 | 3.42E-04 |
| P04278 | 2.25 | 2.36E-02 |
| P27487 | 2.25 | 4.28E-03 |
| Q08431 | 2.25 | 1.41E-02 |
| P0C0L5 | 2.23 | 1.68E-02 |
| Q08830 | 2.23 | 4.29E-02 |
| Q13113 | 2.23 | 7.64E-03 |
| Q5JS37 | 2.22 | 7.75E-03 |
| P51884 | 2.21 | 7.15E-03 |
| P22223 | 2.21 | 1.68E-02 |
| O75531 | 2.21 | 1.80E-02 |
| P01859 | 2.20 | 2.65E-02 |
| Q9Y5Y7 | 2.19 | 3.03E-02 |
| P08236 | 2.18 | 7.25E-03 |
| P17405 | 2.17 | 7.53E-04 |
| P04066 | 2.16 | 1.62E-03 |
| Q14697 | 2.16 | 3.38E-02 |
| Q9HD89 | 2.14 | 5.19E-03 |
| P28070 | 2.13 | 1.35E-02 |
| P07585 | 2.11 | 3.40E-03 |
| P15374 | 2.09 | 2.66E-03 |
| Q6FHJ7 | 2.08 | 1.64E-02 |
| Q13510 | 2.08 | 2.14E-03 |
| Q08380 | 2.06 | 3.75E-03 |
| P02765 | 2.05 | 4.62E-02 |
| P00746 | 2.03 | 4.62E-02 |

| | | |
|---|---|---|
| P15289 | 2.01 | 3.05E-02 |
| Q6GTX8 | 0.50 | 4.26E-02 |
| Q9UBG0 | 0.49 | 1.68E-02 |
| P98082 | 0.48 | 4.95E-02 |
| P98164 | 0.48 | 1.71E-02 |
| P00558 | 0.48 | 2.18E-02 |
| O75084 | 0.48 | 3.01E-02 |
| Q01973 | 0.47 | 4.67E-02 |
| P20827 | 0.47 | 3.03E-02 |
| Q6P531 | 0.46 | 4.82E-02 |
| O75891 | 0.46 | 2.88E-02 |
| O00161 | 0.45 | 4.79E-02 |
| P12259 | 0.45 | 4.33E-02 |
| Q9Y4D7 | 0.45 | 3.38E-02 |
| P09619 | 0.45 | 1.19E-02 |
| Q8NC42 | 0.44 | 3.79E-02 |
| Q13445 | 0.44 | 2.75E-03 |
| P13598 | 0.44 | 2.48E-02 |
| P11586 | 0.43 | 3.46E-02 |
| Q15223 | 0.43 | 1.44E-02 |
| Q9UJJ9 | 0.43 | 4.04E-02 |
| Q9UKU6 | 0.42 | 3.20E-02 |
| P10586 | 0.42 | 3.95E-03 |
| P06744 | 0.42 | 4.89E-02 |
| P15531 | 0.42 | 2.21E-02 |
| P21709 | 0.42 | 2.75E-02 |
| Q7KYR7 | 0.42 | 1.60E-02 |
| P04180 | 0.41 | 1.69E-02 |
| P34896 | 0.41 | 4.61E-02 |
| P09543 | 0.41 | 4.49E-02 |
| P26992 | 0.41 | 4.92E-02 |
| P51148 | 0.41 | 4.75E-02 |
| Q06418 | 0.40 | 2.38E-02 |
| P34932 | 0.40 | 3.33E-02 |
| P30086 | 0.40 | 2.13E-02 |
| P21281 | 0.40 | 9.83E-03 |
| Q99835 | 0.39 | 7.02E-03 |
| O75144 | 0.39 | 1.66E-02 |
| P23526 | 0.39 | 4.37E-02 |

| | | |
|---|---|---|
| Q04760 | 0.39 | 2.70E-02 |
| P10768 | 0.38 | 4.61E-02 |
| Q8N386 | 0.38 | 1.62E-02 |
| Q9BXN2 | 0.38 | 3.36E-02 |
| Q9H4M9 | 0.38 | 8.15E-03 |
| O60494 | 0.37 | 7.75E-03 |
| O75339 | 0.37 | 3.36E-02 |
| Q6EMK4 | 0.37 | 4.76E-02 |
| Q8IWA5 | 0.37 | 4.06E-02 |
| O75351 | 0.37 | 4.59E-02 |
| P29323 | 0.37 | 1.86E-03 |
| Q9Y617 | 0.36 | 4.10E-02 |
| P0DMQ5 | 0.36 | 2.91E-02 |
| Q9P2T1 | 0.36 | 4.89E-02 |
| Q9Y6E0 | 0.36 | 3.08E-02 |
| Q9P0T7 | 0.36 | 4.92E-02 |
| P19022 | 0.36 | 2.33E-02 |
| Q86YT9 | 0.35 | 2.73E-02 |
| P78417 | 0.35 | 4.87E-02 |
| Q9UQB8 | 0.35 | 5.69E-04 |
| O75347 | 0.35 | 1.58E-02 |
| Q9NU53 | 0.34 | 1.19E-02 |
| P82980 | 0.34 | 3.98E-02 |
| Q15262 | 0.34 | 1.66E-02 |
| Q9ULK6 | 0.34 | 3.60E-02 |
| P11047 | 0.34 | 7.12E-03 |
| P19256 | 0.33 | 1.35E-02 |
| P02462 | 0.33 | 4.48E-02 |
| Q15833 | 0.33 | 3.78E-02 |
| O60361 | 0.33 | 2.42E-02 |
| P63096 | 0.33 | 3.52E-02 |
| P15151 | 0.33 | 1.91E-02 |
| Q6UXG3 | 0.33 | 2.01E-02 |
| Q6UX71 | 0.32 | 3.05E-03 |
| P16035 | 0.32 | 2.73E-02 |
| O75882 | 0.32 | 9.15E-03 |
| Q9HCU0 | 0.32 | 1.40E-02 |
| Q13443 | 0.31 | 2.27E-02 |
| Q9HC38 | 0.31 | 3.54E-02 |

| | | |
|---|---|---|
| Q8IUL8 | 0.31 | 2.38E-02 |
| P47755 | 0.31 | 3.98E-02 |
| Q9UNW1 | 0.30 | 4.62E-02 |
| Q12913 | 0.30 | 3.03E-02 |
| Q8TB96 | 0.30 | 5.40E-03 |
| Q92673 | 0.30 | 1.65E-02 |
| Q9NQS3 | 0.30 | 3.16E-03 |
| Q13591 | 0.30 | 4.82E-02 |
| O00560 | 0.29 | 4.53E-02 |
| Q92859 | 0.29 | 3.63E-03 |
| P16070 | 0.29 | 1.51E-02 |
| Q9BY67 | 0.29 | 4.81E-02 |
| P15291 | 0.29 | 1.18E-02 |
| Q9HBB8 | 0.28 | 3.81E-02 |
| Q8N5I2 | 0.28 | 2.48E-02 |
| P24855 | 0.28 | 1.19E-02 |
| Q96RW7 | 0.28 | 2.86E-02 |
| P01042 | 0.27 | 4.88E-02 |
| Q12860 | 0.26 | 3.79E-02 |
| O94856 | 0.26 | 4.82E-02 |
| Q9HCM3 | 0.26 | 1.04E-02 |
| Q96DG6 | 0.26 | 3.03E-02 |
| Q15116 | 0.26 | 1.29E-02 |
| Q7Z7G0 | 0.25 | 3.12E-02 |
| O43895 | 0.25 | 2.91E-02 |
| P13489 | 0.25 | 2.91E-02 |
| Q6V0I7 | 0.24 | 3.35E-02 |
| Q8NBJ4 | 0.24 | 3.51E-02 |
| P58499 | 0.24 | 1.70E-02 |
| Q96JQ0 | 0.24 | 5.35E-03 |
| Q5SZK8 | 0.24 | 6.15E-03 |
| Q9UN37 | 0.24 | 1.24E-02 |
| Q8WUM4 | 0.24 | 8.83E-03 |
| P23284 | 0.22 | 1.39E-02 |
| Q6P9A2 | 0.22 | 1.87E-02 |
| Q9UEF7 | 0.22 | 3.67E-02 |
| Q99816 | 0.22 | 1.94E-02 |
| Q8NDA2 | 0.21 | 2.60E-02 |
| P22607 | 0.21 | 4.26E-02 |

| | | |
|---|---|---|
| Q96MU8 | 0.21 | 1.58E-02 |
| Q9Y2G1 | 0.20 | 1.13E-02 |
| Q9Y5K6 | 0.20 | 1.66E-02 |
| P14784 | 0.20 | 1.35E-02 |
| P41217 | 0.20 | 1.13E-02 |
| Q9NR34 | 0.19 | 3.03E-02 |
| Q9UK41 | 0.19 | 9.40E-03 |
| Q9Y6W3 | 0.18 | 4.69E-02 |
| Q7LBR1 | 0.18 | 1.42E-02 |
| P55017 | 0.18 | 1.71E-02 |
| P50148 | 0.17 | 1.91E-02 |
| Q96EY5 | 0.17 | 7.75E-03 |
| Q9NP79 | 0.16 | 1.97E-02 |
| Q9H008 | 0.15 | 4.02E-03 |
| P0C7U0 | 0.14 | 1.03E-02 |
| Q9H1U4 | 0.13 | 9.26E-03 |
| P45877 | 0.11 | 6.09E-03 |
| Q9H9H4 | 0.10 | 1.51E-02 |
| Q9H444 | 0.10 | 4.14E-02 |
| P13688 | 0.07 | 4.88E-04 |
| P31997 | 0.06 | 3.02E-02 |

| F11 | | |
|---|---|---|
| **Protein Accessions** | **Fold Change** | **P value** |
| P05534 | 20.44 | 2.03E-07 |
| P01911 | 19.95 | 3.80E-07 |
| Q86SG5 | 17.98 | 1.37E-05 |
| Q15293 | 16.09 | 9.38E-04 |
| P18465 | 14.92 | 2.17E-09 |
| P30508 | 13.84 | 2.02E-06 |
| P31151 | 13.04 | 2.92E-05 |
| P04179 | 12.61 | 3.80E-07 |
| P01721 | 12.59 | 1.46E-07 |
| P02750 | 12.02 | 7.60E-07 |
| P01699 | 11.98 | 1.13E-08 |
| A0A0C4DH72 | 11.95 | 7.89E-06 |
| P61769 | 10.46 | 1.31E-06 |
| P01717 | 10.32 | 1.36E-07 |
| P01701 | 9.76 | 3.80E-07 |
| P28065 | 9.54 | 1.13E-08 |
| Q92876 | 9.47 | 4.22E-04 |
| P01714 | 9.03 | 1.33E-06 |
| P02763 | 8.52 | 3.01E-06 |
| Q9Y547 | 8.47 | 2.32E-06 |
| P01703 | 8.32 | 1.42E-06 |
| P01903 | 8.18 | 1.64E-04 |
| Q13291 | 7.98 | 3.54E-06 |
| P13796 | 7.80 | 9.00E-06 |
| A0A075B6S5 | 7.37 | 1.17E-05 |
| O00244 | 7.27 | 1.92E-04 |
| P01700 | 7.11 | 3.74E-02 |
| P24158 | 7.08 | 2.70E-02 |
| Q9NWV4 | 6.93 | 5.22E-05 |
| P0DOX8 | 6.90 | 2.01E-06 |
| P01715 | 6.89 | 7.60E-07 |
| P06312 | 6.55 | 1.37E-05 |
| P05109 | 6.48 | 1.80E-02 |
| P01619 | 6.46 | 1.56E-04 |
| A0A075B6K5 | 6.40 | 2.01E-04 |
| Q07075 | 6.27 | 6.83E-04 |
| P01599 | 6.20 | 2.80E-04 |
| P15814 | 6.16 | 2.92E-05 |
| Q71DI3 | 6.11 | 3.87E-03 |
| Q9H299 | 6.06 | 1.86E-03 |
| P0DOX3 | 5.82 | 1.79E-03 |
| A0A0J9YXX1 | 5.80 | 1.23E-05 |
| Q9UBG3 | 5.75 | 1.80E-02 |

| | | |
|---|---|---|
| P59666 | 5.62 | 1.61E-02 |
| Q15274 | 5.48 | 1.56E-04 |
| P01624 | 5.45 | 1.49E-04 |
| P06702 | 5.44 | 2.99E-02 |
| P23381 | 5.43 | 9.00E-06 |
| P02654 | 5.30 | 6.25E-05 |
| Q8N357 | 5.24 | 1.91E-05 |
| Q969H8 | 5.23 | 4.26E-04 |
| P02792 | 5.13 | 3.76E-04 |
| Q06495 | 5.10 | 1.09E-03 |
| Q92542 | 5.10 | 1.91E-04 |
| Q9NZT1 | 5.08 | 7.10E-03 |
| P16401 | 5.00 | 1.02E-02 |
| Q15084 | 4.91 | 1.49E-04 |
| P27824 | 4.76 | 3.76E-04 |
| A0A075B6J9 | 4.76 | 7.37E-04 |
| Q9P1F3 | 4.72 | 7.76E-05 |
| A0A075B6K4 | 4.70 | 1.82E-02 |
| P29966 | 4.61 | 4.28E-04 |
| P0C0S8 | 4.61 | 4.29E-02 |
| P16189 | 4.52 | 5.87E-04 |
| O43707 | 4.46 | 1.44E-04 |
| P37837 | 4.46 | 1.12E-02 |
| Q9H3K6 | 4.45 | 1.23E-02 |
| Q92887 | 4.41 | 3.03E-04 |
| Q8WU39 | 4.38 | 1.37E-04 |
| P28908 | 4.36 | 3.74E-07 |
| P62805 | 4.35 | 2.03E-02 |
| P19652 | 4.32 | 2.83E-04 |
| Q9NUM4 | 4.30 | 3.03E-04 |
| P09923 | 4.25 | 1.35E-02 |
| Q9NRX4 | 4.22 | 2.14E-03 |
| O95980 | 4.19 | 2.49E-03 |
| P52272 | 4.17 | 2.43E-02 |
| P06681 | 4.08 | 3.53E-03 |
| A0A0C4DH24 | 4.06 | 1.67E-03 |
| A0A0B4J1V2 | 4.06 | 2.83E-04 |
| P01011 | 4.02 | 1.49E-04 |
| P36980 | 4.00 | 5.58E-04 |
| Q9Y274 | 3.93 | 1.37E-03 |
| P04211 | 3.89 | 8.42E-05 |
| P06899 | 3.89 | 4.18E-02 |
| P07384 | 3.85 | 8.02E-03 |
| P25311 | 3.78 | 7.08E-03 |
| O60234 | 3.77 | 7.52E-04 |

| | | |
|---|---|---|
| P02545 | 3.74 | 3.42E-02 |
| P02741 | 3.71 | 2.26E-04 |
| P07451 | 3.66 | 1.90E-04 |
| P25815 | 3.64 | 2.08E-02 |
| A0A075B6P5 | 3.63 | 4.25E-02 |
| P54793 | 3.62 | 1.59E-03 |
| P26842 | 3.61 | 4.23E-04 |
| P35606 | 3.55 | 5.22E-05 |
| P0DOX6 | 3.54 | 1.81E-04 |
| P20333 | 3.53 | 9.04E-03 |
| P09972 | 3.53 | 2.28E-02 |
| P78423 | 3.52 | 2.03E-03 |
| P12104 | 3.49 | 1.74E-02 |
| Q15116 | 3.48 | 1.91E-05 |
| Q8N2U0 | 3.45 | 2.59E-03 |
| Q8WW52 | 3.45 | 3.86E-02 |
| Q14764 | 3.44 | 2.08E-04 |
| A0A075B6I0 | 3.44 | 2.65E-03 |
| A0A0C4DH68 | 3.43 | 1.69E-04 |
| P13797 | 3.43 | 8.85E-03 |
| A0A0C4DH25 | 3.42 | 4.12E-03 |
| O00754 | 3.37 | 5.82E-05 |
| P00747 | 3.37 | 1.06E-03 |
| Q8NHJ6 | 3.35 | 2.79E-04 |
| Q13421 | 3.35 | 1.41E-02 |
| P0DOX7 | 3.34 | 9.16E-04 |
| P00736 | 3.34 | 6.17E-04 |
| O75368 | 3.29 | 1.12E-03 |
| Q03591 | 3.29 | 2.41E-02 |
| Q9Y6X5 | 3.27 | 5.58E-04 |
| Q9NZT2 | 3.27 | 1.52E-02 |
| P04632 | 3.25 | 1.02E-02 |
| Q9BUH6 | 3.25 | 1.04E-03 |
| O75531 | 3.24 | 8.01E-04 |
| P25774 | 3.24 | 4.06E-04 |
| P01593 | 3.23 | 2.38E-03 |
| P01871 | 3.17 | 1.28E-03 |
| A2NJV5 | 3.16 | 1.67E-03 |
| O96009 | 3.14 | 1.04E-03 |
| P01877 | 3.13 | 7.22E-04 |
| Q96FW1 | 3.11 | 2.37E-02 |
| P26447 | 3.11 | 4.15E-02 |
| A0A0C4DH38 | 3.09 | 2.63E-03 |
| P20774 | 3.02 | 2.03E-03 |
| Q13113 | 3.01 | 8.01E-04 |

| | | |
|---|---|---|
| Q6UWV6 | 2.99 | 5.58E-04 |
| P01601 | 2.95 | 3.72E-02 |
| P21796 | 2.94 | 2.14E-03 |
| Q03167 | 2.92 | 2.60E-03 |
| A0A0B4J1Y9 | 2.91 | 1.80E-02 |
| O00584 | 2.88 | 1.05E-03 |
| P14324 | 2.83 | 4.73E-02 |
| Q9BTY2 | 2.83 | 7.25E-04 |
| A0A087WW87 | 2.81 | 2.74E-02 |
| P06331 | 2.80 | 2.49E-03 |
| P04217 | 2.78 | 2.38E-02 |
| P54803 | 2.74 | 3.67E-04 |
| ENSEMBL:ENSBTAP00000007350 | 2.73 | 9.08E-03 |
| P11279 | 2.73 | 3.29E-03 |
| Q6UWR7 | 2.72 | 2.30E-02 |
| P28070 | 2.72 | 3.29E-03 |
| A0A0A0MRZ8 | 2.72 | 8.45E-03 |
| P60900 | 2.70 | 5.56E-03 |
| Q96C19 | 2.68 | 1.79E-02 |
| P08670 | 2.68 | 3.78E-02 |
| P15144 | 2.67 | 3.92E-02 |
| P00450 | 2.66 | 4.07E-03 |
| P01019 | 2.65 | 8.01E-04 |
| P14151 | 2.62 | 2.23E-03 |
| Q86VB7 | 2.59 | 1.49E-04 |
| P36639 | 2.58 | 3.90E-03 |
| P25789 | 2.56 | 3.01E-03 |
| Q8TCD5 | 2.56 | 3.53E-03 |
| P01876 | 2.56 | 1.61E-02 |
| O60361 | 2.55 | 3.70E-04 |
| P04075 | 2.55 | 2.30E-02 |
| P29401 | 2.55 | 9.04E-03 |
| P27105 | 2.54 | 7.34E-03 |
| Q9UHY7 | 2.53 | 3.91E-03 |
| P0DOX5 | 2.52 | 2.29E-04 |
| P08195 | 2.51 | 7.56E-03 |
| P26368 | 2.50 | 9.81E-03 |
| Q8NFQ8 | 2.47 | 2.30E-02 |
| P15531 | 2.46 | 2.85E-04 |
| P36969 | 2.44 | 1.31E-04 |
| P20618 | 2.44 | 3.41E-02 |
| Q8NCC3 | 2.43 | 4.14E-03 |
| P36222 | 2.40 | 8.85E-03 |
| P01780 | 2.40 | 1.46E-03 |
| O60814 | 2.38 | 8.35E-03 |

| | | |
|---|---|---|
| P30101 | 2.37 | 3.40E-02 |
| P01591 | 2.37 | 1.02E-02 |
| Q8NBS9 | 2.34 | 1.07E-03 |
| Q9BXS4 | 2.34 | 1.89E-02 |
| Q8WWB7 | 2.34 | 2.86E-03 |
| P25786 | 2.32 | 1.59E-03 |
| Q92626 | 2.32 | 1.63E-03 |
| P04066 | 2.31 | 1.13E-03 |
| O95998 | 2.31 | 3.97E-03 |
| A0A0C4DH29 | 2.30 | 2.64E-02 |
| Q8TDQ1 | 2.28 | 7.51E-03 |
| Q99571 | 2.28 | 2.32E-02 |
| A0A0A0MS15 | 2.28 | 1.47E-02 |
| O00499 | 2.27 | 1.56E-02 |
| P01344 | 2.26 | 1.83E-02 |
| P02749 | 2.25 | 4.73E-02 |
| P05091 | 2.24 | 3.47E-02 |
| P15586 | 2.23 | 4.94E-04 |
| P09871 | 2.23 | 7.23E-04 |
| P81605 | 2.21 | 3.55E-02 |
| P28066 | 2.20 | 4.12E-03 |
| Q9BTM9 | 2.20 | 1.77E-02 |
| P27487 | 2.19 | 7.37E-03 |
| Q9UBR2 | 2.17 | 2.92E-05 |
| A0A075B6Q5 | 2.17 | 3.11E-02 |
| P21217 | 2.16 | 2.60E-02 |
| P08571 | 2.16 | 4.16E-02 |
| P17050 | 2.13 | 3.97E-03 |
| O14818 | 2.12 | 6.98E-03 |
| Q13867 | 2.11 | 2.73E-02 |
| P13798 | 2.11 | 1.27E-02 |
| P05981 | 2.11 | 7.23E-04 |
| Q03405 | 2.10 | 3.62E-02 |
| P08236 | 2.07 | 1.49E-02 |
| P05155 | 2.07 | 4.96E-03 |
| Q02985 | 2.06 | 3.29E-03 |
| P02511 | 2.03 | 4.16E-02 |
| Q9BXJ7 | 2.01 | 3.11E-02 |
| Q96CX2 | 2.00 | 3.52E-02 |
| P09619 | 0.50 | 2.30E-02 |
| P43121 | 0.50 | 1.27E-02 |
| Q9BQ51 | 0.49 | 4.63E-03 |
| Q6UX71 | 0.49 | 1.56E-02 |
| Q9HBR0 | 0.48 | 2.31E-02 |
| Q92859 | 0.47 | 1.83E-02 |

| | | |
|---|---|---|
| O75144 | 0.47 | 3.54E-02 |
| P55899 | 0.46 | 3.56E-02 |
| Q86VN1 | 0.45 | 3.61E-02 |
| Q9Y4D7 | 0.44 | 4.09E-02 |
| P29323 | 0.44 | 4.50E-03 |
| Q15223 | 0.44 | 1.45E-02 |
| P30086 | 0.44 | 3.56E-02 |
| Q8WUM4 | 0.43 | 3.78E-02 |
| P21709 | 0.43 | 3.67E-02 |
| Q8N386 | 0.42 | 1.24E-02 |
| P07195 | 0.41 | 4.83E-02 |
| P00568 | 0.41 | 2.50E-02 |
| P41217 | 0.41 | 4.65E-02 |
| P40189 | 0.41 | 3.78E-02 |
| Q6UXG3 | 0.41 | 4.08E-02 |
| Q9NQS3 | 0.41 | 9.29E-03 |
| P10586 | 0.40 | 2.52E-03 |
| P24855 | 0.40 | 3.11E-02 |
| Q9BWV1 | 0.40 | 4.73E-02 |
| P13598 | 0.39 | 2.30E-02 |
| P22304 | 0.39 | 2.78E-02 |
| Q07507 | 0.39 | 7.72E-03 |
| P30530 | 0.38 | 1.30E-03 |
| P0DMQ5 | 0.38 | 3.93E-02 |
| Q04760 | 0.37 | 3.10E-02 |
| Q05707 | 0.37 | 3.10E-02 |
| Q9H0X4 | 0.37 | 3.05E-02 |
| Q9H4M9 | 0.37 | 9.37E-03 |
| P0C7U0 | 0.36 | 4.16E-02 |
| O94760 | 0.36 | 2.10E-02 |
| Q8N5I2 | 0.36 | 4.96E-02 |
| P10768 | 0.36 | 4.89E-02 |
| Q9ULK6 | 0.36 | 4.71E-02 |
| Q8TBP5 | 0.35 | 4.15E-02 |
| Q9Y5E6 | 0.35 | 2.05E-02 |
| P14784 | 0.35 | 3.78E-02 |
| Q9UBG0 | 0.34 | 5.14E-03 |
| P07741 | 0.34 | 4.12E-02 |
| P25940 | 0.34 | 2.15E-02 |
| Q96MU8 | 0.33 | 3.79E-02 |
| P19256 | 0.33 | 1.62E-02 |
| Q9NP85 | 0.33 | 3.67E-02 |
| Q9UNN8 | 0.32 | 1.51E-03 |
| Q8IUL8 | 0.32 | 3.11E-02 |
| P50148 | 0.31 | 4.80E-02 |

| | | |
|---|---|---|
| Q6YHK3 | 0.30 | 4.83E-02 |
| Q9Y639 | 0.30 | 4.32E-02 |
| Q96RW7 | 0.30 | 3.79E-02 |
| Q13443 | 0.29 | 2.60E-02 |
| P35555 | 0.29 | 3.78E-02 |
| P26992 | 0.29 | 3.25E-02 |
| P11362 | 0.29 | 4.25E-02 |
| P05556 | 0.27 | 3.78E-02 |
| P11047 | 0.26 | 5.14E-03 |
| P22105 | 0.25 | 2.56E-02 |
| P82980 | 0.25 | 3.06E-02 |
| Q9HCU0 | 0.25 | 1.14E-02 |
| P09486 | 0.25 | 1.73E-02 |
| Q9Y6N8 | 0.24 | 2.85E-02 |
| O43921 | 0.23 | 1.67E-02 |
| P48745 | 0.22 | 3.87E-02 |
| A1L4H1 | 0.22 | 4.16E-02 |
| Q96JQ0 | 0.20 | 5.14E-03 |
| P98095 | 0.17 | 4.83E-02 |
| P80370 | 0.15 | 3.47E-02 |
| Q9HCM3 | 0.13 | 3.68E-03 |
| Q9H1U4 | 0.11 | 1.04E-02 |
| O75339 | 0.11 | 9.81E-03 |
| Q8NDA2 | 0.10 | 1.84E-02 |
| P55017 | 0.10 | 1.49E-02 |
| P10721 | 0.09 | 3.67E-02 |

| group analysis ( FUO & healthy people) | | |
|---|---|---|
| **Protein Accessions** | **Fold Change** | **P value** |
| P02792 | 6.18 | 3.08E-02 |
| Q07075 | 4.47 | 2.63E-02 |
| O00754 | 3.97 | 3.07E-02 |
| Q9NUM4 | 3.66 | 9.87E-03 |
| P02656 | 3.62 | 1.25E-02 |
| P02741 | 3.51 | 3.21E-02 |
| Q7L5L3 | 3.46 | 4.39E-02 |
| P25774 | 3.33 | 4.04E-03 |
| Q9UHI7 | 2.84 | 4.22E-02 |
| Q9BTY2 | 2.81 | 1.92E-02 |
| Q8NCC3 | 2.68 | 9.95E-03 |
| P53634 | 2.60 | 4.56E-02 |
| O96009 | 2.55 | 4.22E-02 |
| P04066 | 2.53 | 4.89E-02 |
| P07858 | 2.35 | 4.46E-02 |
| P17050 | 2.30 | 1.01E-02 |
| P00450 | 2.16 | 2.95E-02 |
| P06865 | 2.16 | 3.41E-02 |
| Q13510 | 2.06 | 3.44E-02 |
| P10619 | 2.03 | 3.39E-02 |
| O15197 | 0.50 | 1.47E-02 |
| Q86UD1 | 0.50 | 2.10E-02 |
| Q7KYR7 | 0.50 | 2.62E-03 |
| Q9H8L6 | 0.50 | 4.52E-02 |
| Q8TBP5 | 0.50 | 4.63E-03 |
| P14784 | 0.50 | 5.47E-03 |
| P29622 | 0.50 | 4.38E-02 |
| Q9UBQ6 | 0.49 | 1.19E-02 |
| Q5VW32 | 0.49 | 2.85E-02 |
| Q8NFZ8 | 0.49 | 1.65E-02 |
| P54753 | 0.49 | 1.09E-02 |
| Q9NP85 | 0.49 | 3.53E-03 |
| Q9Y2G1 | 0.49 | 7.29E-03 |
| P04745 | 0.49 | 4.53E-02 |
| Q9Y639 | 0.49 | 6.68E-03 |
| Q7Z7G0 | 0.49 | 1.21E-02 |
| Q99574 | 0.49 | 3.70E-02 |

| | | |
|---|---|---|
| Q7LBR1 | 0.48 | 1.18E-02 |
| Q9H444 | 0.48 | 3.69E-02 |
| Q6UXI9 | 0.48 | 2.64E-02 |
| P35858 | 0.48 | 1.88E-02 |
| Q6UXD5 | 0.48 | 8.35E-03 |
| Q13443 | 0.48 | 5.01E-03 |
| P16035 | 0.48 | 6.02E-03 |
| O75131 | 0.48 | 3.87E-02 |
| Q13591 | 0.48 | 5.30E-03 |
| P09619 | 0.47 | 4.00E-04 |
| Q86YT9 | 0.47 | 2.99E-03 |
| Q9UQV4 | 0.47 | 4.16E-02 |
| Q13477 | 0.47 | 2.14E-02 |
| Q09328 | 0.47 | 1.30E-02 |
| Q03403 | 0.47 | 4.57E-02 |
| Q9UPZ6 | 0.47 | 2.37E-02 |
| Q8WVQ1 | 0.47 | 2.90E-02 |
| P17342 | 0.47 | 1.88E-02 |
| P08582 | 0.46 | 1.18E-02 |
| P22607 | 0.46 | 9.87E-03 |
| Q6V0I7 | 0.46 | 1.01E-02 |
| Q53RD9 | 0.46 | 2.43E-02 |
| Q15303 | 0.46 | 1.63E-02 |
| P11362 | 0.45 | 2.74E-03 |
| O14594 | 0.45 | 1.92E-02 |
| Q5IJ48 | 0.45 | 3.95E-03 |
| P41217 | 0.45 | 9.61E-03 |
| Q9ULK6 | 0.45 | 2.62E-03 |
| Q9UKY0 | 0.45 | 1.10E-02 |
| P21926 | 0.44 | 9.99E-03 |
| P15151 | 0.44 | 5.67E-04 |
| Q96RW7 | 0.44 | 1.32E-03 |
| Q14574 | 0.44 | 1.47E-02 |
| Q9UBG0 | 0.44 | 8.43E-05 |
| P12273 | 0.44 | 3.91E-02 |
| P13598 | 0.44 | 7.25E-05 |
| P42702 | 0.43 | 3.08E-02 |
| P24855 | 0.43 | 7.97E-04 |
| Q8TB96 | 0.43 | 7.89E-04 |

| | | |
|---|---|---|
| P19961 | 0.43 | 1.90E-02 |
| O75144 | 0.42 | 1.29E-04 |
| Q9UNN8 | 0.42 | 1.48E-05 |
| P98095 | 0.42 | 1.64E-02 |
| Q6UXG3 | 0.42 | 5.81E-04 |
| Q8WZ75 | 0.41 | 1.47E-02 |
| Q9H9P2 | 0.41 | 2.99E-03 |
| P19256 | 0.41 | 1.26E-04 |
| Q14982 | 0.41 | 1.88E-02 |
| Q9NQS3 | 0.41 | 1.77E-05 |
| Q6UVK1 | 0.41 | 6.85E-03 |
| P01042 | 0.41 | 3.53E-03 |
| O94856 | 0.41 | 3.88E-03 |
| Q9P121 | 0.41 | 1.79E-02 |
| Q96DR8 | 0.41 | 3.61E-02 |
| Q8IUL8 | 0.40 | 1.92E-03 |
| Q8TER0 | 0.39 | 4.69E-03 |
| Q9BUN1 | 0.39 | 1.92E-02 |
| Q9UNE0 | 0.39 | 6.95E-03 |
| Q96NY8 | 0.39 | 2.54E-04 |
| Q8IUK5 | 0.39 | 2.92E-02 |
| P14384 | 0.39 | 4.59E-03 |
| Q16849 | 0.38 | 4.22E-02 |
| Q9H159 | 0.38 | 1.47E-02 |
| Q9NPG4 | 0.38 | 4.52E-02 |
| P06870 | 0.38 | 2.19E-02 |
| P0C7U0 | 0.38 | 7.93E-04 |
| P05154 | 0.38 | 6.07E-03 |
| Q92859 | 0.38 | 7.25E-05 |
| Q04756 | 0.38 | 8.60E-03 |
| P10912 | 0.38 | 9.61E-03 |
| Q9BY67 | 0.38 | 7.93E-04 |
| P26992 | 0.37 | 2.78E-04 |
| P11047 | 0.37 | 2.78E-04 |
| Q68CJ9 | 0.36 | 4.79E-03 |
| Q8NFT8 | 0.36 | 8.58E-03 |
| P50995 | 0.35 | 4.06E-02 |
| Q96JQ0 | 0.35 | 7.25E-05 |
| Q9HCU0 | 0.35 | 2.21E-04 |

| | | |
|---|---|---|
| Q6UY11 | 0.35 | 7.97E-04 |
| P23327 | 0.34 | 2.62E-03 |
| P01133 | 0.34 | 1.83E-02 |
| P22891 | 0.34 | 1.88E-02 |
| Q9H1U4 | 0.34 | 4.58E-04 |
| Q9UN70 | 0.34 | 9.95E-03 |
| P55083 | 0.33 | 8.79E-03 |
| P80370 | 0.32 | 7.89E-04 |
| P31997 | 0.31 | 2.62E-03 |
| Q5VY43 | 0.31 | 6.35E-03 |
| P39059 | 0.30 | 3.53E-03 |
| O75339 | 0.29 | 7.25E-05 |
| Q8NDA2 | 0.23 | 7.25E-05 |
| O43852 | 0.07 | 2.74E-03 |
